# Genome-wide Association Study of Traumatic Brain Injury in U.S. Military Veterans Enrolled in the VA Million Veteran Program

**DOI:** 10.1101/2023.02.16.23286045

**Authors:** Victoria C. Merritt, Adam X. Maihofer, Marianna Gasperi, Elizabeth Ketema, Catherine Chanfreau-Coffinier, Murray B. Stein, Matthew S. Panizzon, Richard L. Hauger, Mark W. Logue, Lisa Delano-Wood, Caroline M. Nievergelt, VA Million Veteran Program

## Abstract

Large-scale genetic studies of traumatic brain injury (TBI) are lacking; thus, our understanding of the influence of genetic factors on TBI risk and recovery is incomplete. This study aimed to conduct a genome-wide association study (GWAS) of TBI in VA Million Veteran Program enrollees. Participants included a multi-ancestry cohort (European, African, and Hispanic ancestries; N=304,485; 111,494 TBI cases, 192,991 controls). TBI was assessed using MVP survey data and ICD codes from the Veterans Health Administration’s electronic health record. GWAS was performed using logistic regression in PLINK, and meta-analyzed in METAL. FUMA was used for post-GWAS analysis. Genomic structural equation modeling (gSEM) was conducted to investigate underlying genetic associations with TBI, and bivariate MiXeR was used to estimate phenotype specific and shared polygenicity. SNP-based heritability was 0.060 (SE=0.004, *p*=7.83×10^−66^). GWAS analysis identified 15 genome-wide significant (GWS) loci at *p*<5×10^−8^. Gene-based analyses revealed 14 gene-wide significant genes, including *NCAM1, APOE, FTO*, and *FOXP2*. Gene tissue expression analysis identified the brain as significantly enriched, particularly in the frontal cortex, anterior cingulate cortex, and nucleus accumbens. Genetic correlations with TBI were significant for risk-taking behaviors and psychiatric disorders, but generally not significant for the neurocognitive variables investigated. gSEM analysis revealed stronger associations with risk-taking traits than with psychiatric traits. Finally, the genetic architecture of TBI was similar to polygenic psychiatric disorders. Neurodegenerative disorders including Alzheimer’s and Parkinson’s disease showed much less polygenicity, however, the proportion of shared variance with TBI was high. This first well-powered GWAS of TBI identified 15 loci including genes relevant to TBI biology, and showed that TBI is a heritable trait with comparable genetic architecture and high genetic correlation with psychiatric traits. Our findings set the stage for future TBI GWASs that focus on injury severity and diversity and chronicity of symptom sequelae.

## Introduction

Military personnel serving in Iraq and Afghanistan have sustained traumatic brain injuries (TBIs) at a higher frequency than prior conflicts^1^, prompting widespread interest in the assessment and management of TBI and its associated sequelae. Within military populations, the overwhelming majority of TBIs are considered “closed” head injuries; among these, approximately 80% are classified as mild in severity^2^. Although designated as “mild,” it has become well established that a host of clinically meaningful sequelae (i.e., neurobehavioral symptoms, psychiatric distress, and problems with thinking and cognition) develop acutely post-injury^3-5^. Despite the expectation of full recovery within days to weeks following mild TBI (mTBI)^6^, these difficulties can persist for many years for some individuals^7-10^. Moreover, long-term outcome studies, even in mTBI, have demonstrated an association between TBI and increased risk of dementia, including Alzheimer’s disease (AD), as well as chronic symptoms of posttraumatic stress disorder (PTSD) and depression^8,11^.

Accumulating evidence also suggests that Veterans with a history of TBI— especially when combined with psychiatric comorbidities—experience poor health-related quality of life^12-14^, increased difficulties with psychosocial functioning^15^, and greater global disability^16^. The costs associated with TBI assessment and management are also substantial, with one study estimating that the median annual healthcare cost per patient was approximately four times higher for Veterans with a history of TBI compared to Veterans without a history of TBI^17^. It is thus crucial to deepen our understanding of the risk factors associated with *sustaining* a TBI as well as the underlying factors that contribute to poor outcome and recovery *following* TBI.

It has been hypothesized that there are several mechanisms by which genetics may influence risk for and response to neurotrauma, including pre-injury risk factors for sustaining a TBI (e.g., risk-taking behaviors, impulsivity, substance use); the acute response to neurotrauma (i.e., extent of injury); the repair or recovery processes following injury (i.e., capacity for recovery); and pre- and post-injury cognitive capacity and cognitive reserve^18^. However, large-scale genetic studies of TBI are lacking and thus our understanding of the influence of genetic factors on TBI risk and recovery remains incomplete. Examining the role of genetics in TBI risk and recovery may offer novel mechanistic insights and lead to important discoveries that can inform preventive efforts and innovative treatments for affected individuals.

Among the published studies that have examined genetic associations with TBI risk and recovery, findings are considerably disparate, likely due to varied methodologies as well as inadequate sample sizes and therefore low power to detect meaningful differences in TBI samples^18-23^. Furthermore, existing studies have largely adopted a “candidate gene” approach, focusing on a specific gene of interest—most commonly, the apolipoprotein E *(APOE)* gene^24-28^, though it is widely believed that genetic predisposition to complex traits or conditions is highly polygenic. Thus, not only are adequately powered studies needed to better understand the influence of genetic markers on TBI risk and recovery but, given the likelihood that multiple genes may be involved, critical next steps are to apply the concept of polygenic risk to TBI and capitalize on recent advances in genetics and genomics research—namely, the implementation of genome-wide association studies.

Until recently, it had been very challenging to amass a sufficient number of cases to adequately carry out a genome-wide association study (GWAS) in TBI. However, the VA Million Veteran Program, a nationwide research initiative that examines how genes influence health and behaviors^29^, affords a unique opportunity to explore a large cohort of U.S. military Veterans with and without a history of TBI. In this GWAS of TBI using Veterans Health Administration’s (VHA) electronic health record data, we assessed TBI history among military Veterans. We examined genome-wide significant (GWS) variants associated with TBI in a multi-ancestry sample, and separately examined Veterans of European ancestry (EA), African ancestry (AA), and Hispanic ancestry (HA). We examined replication of results with data from another biobank, FinnGen (which included individuals of EA only), and assessed SNP-based heritability. We then examined genetic correlations with various health domains, including risk-taking behaviors, psychiatric disorders, neurocognition, and brain morphometrics, and evaluated phenotype-specific and shared polygenicity among these traits.

## Methods

### Procedures

This research was conducted as part of project “MVP026” within the Million Veteran Program (MVP). The MVP, sponsored by the Department of Veterans Affairs Office of Research and Development, is a nationwide initiative offering military Veterans the opportunity to participate in research that seeks to better understand how genetic factors influence health. Upon consenting to participate in MVP, participants are asked to complete two comprehensive surveys—the “Baseline Survey” and “Lifestyle Survey”—and provide a blood sample for genetic analysis. Participants also consent to provide MVP investigators access to their VHA electronic health record (EHR). MVP study procedures have been previously described by Gaziano and colleagues^29^. The overarching MVP project was approved by the VA Central Institutional Review Board (IRB) in 2010 and this specific project (“MVP026”) received VA Central IRB approval in 2019.

### Participants

Veterans (N=304,485; 111,494 TBI cases; 192,991 controls) were included in this study if they had both phenotype and genotyping data available (described below in more detail). The EA cohort included 249,785 Veterans (85,613 cases; 164,172 controls); the AA cohort included 35,470 Veterans (15,714 cases; 19,756 controls); and the HA cohort included 19,230 Veterans (10,167 cases; 9,063 controls). See Table 1 for overall (multi-ancestry) sample characteristics and Supplemental Table 1 for EA, AA, and HA sample characteristics.

**Table 1.** Multi-ancestry participant characteristics.

|  | **Overall**<br>**N=304,485** | **TBI Case**<br>**n=111,494** | **Control**<br>**n=192,991** | ***p*** |
| --- | --- | --- | --- | --- |
| Age, Mean (SD) | 64.36 (13.16) | 59.92 (14.55) | 66.92 (11.54) | <.001 |
| Sex, N (%) |  |  |  |  |
| Male | 281,108 (92.3) | 103,378 (92.7) | 177,730 (92.1) | <.001 |
| Female | 23,370 (7.7) | 8,109 (7.3) | 15,261 (7.9) |  |
| Missing | 7 (0.0) | 7 (0.0) | 0 (0.0) |  |
| Ancestry, N (%) |  |  |  |  |
| European | 249,785 (82.0) | 85,613 (76.8) | 164,172 (85.1) | <.001 |
| African | 35,470 (11.6) | 15,714 (14.1) | 19,756 (10.2) |  |
| Hispanic | 19,230 (6.3) | 10,167 (9.1) | 9,063 (4.7) |  |
| Data Source, N (%) |  |  |  |  |
| BL | 9,039 (3.0) | 9,039 (8.1) | -- | -- |
| LS | 4,101 (1.3) | 4,101 (3.7) |  |  |
| ICD | 23,776 (7.8) | 23,776 (21.3) |  |  |
| BL + LS | 45,270 (14.8) | 44,544 (40.0) |  |  |
| BL + ICD | 11,557 (3.8) | 11,557 (10.4) |  |  |
| LS + ICD | 2,558 (0.8) | 2,558 (2.3) |  |  |
| BL + LS + ICD | 208,184 (68.4) | 15,919 (14.3) |  |  |
| TBI Source, N (%) |  |  |  |  |
| BL-CC | 6,623 (2.2) | 6,623 (5.9) | -- | -- |
| BL-TBI | 1,251 (0.4) | 1,251 (1.1) |  |  |
| LS | 4,101 (1.3) | 4,101 (3.7) |  |  |
| ICD | 58,606 (19.2) | 57,880 (51.9) |  |  |
| BL-CC + BL-TBI | 1,165 (0.4) | 1,165 (1.0) |  |  |
| BL-CC + LS | 13,982 (4.6) | 13,982 (12.5) |  |  |
| BL-CC + ICD | 1,399 (0.5) | 1,399 (1.3) |  |  |
| BL-TBI + ICD | 3,138 (1.0) | 3,138 (2.8) |  |  |
| LS + ICD | 205,154 (67.4) | 12,889 (11.6) |  |  |
| BL-CC + BL-TBI + ICD | 3,478 (1.1) | 3,478 (3.1) |  |  |
| BL-CC + LS + ICD | 2,106 (0.7) | 2,106 (1.9) |  |  |
| BL-TBI + LS + ICD | 1,419 (0.5) | 1,419 (1.3) |  |  |
| All 4 Sources | 2,063 (0.7) | 2,063 (1.9) |  |  |

Abbreviations: BL = MVP Baseline Survey; LS = MVP Lifestyle Survey; ICD = International Statistical Classification of Diseases; BL-CC = MVP Baseline Survey, Concussion or Loss of Consciousness; BL-TBI = MVP Baseline Survey, Traumatic Brain Injury

### MVP Data Sources

The following data sources (from MVP v20_1 data release) were used to gather information pertaining to TBI history: MVP Baseline Survey, MVP Lifestyle Survey, and *International Classification of Diseases, 9*^*th*^ *Revision Clinical Modification* (ICD-9) and *International Classification of Diseases, 10*^*th*^ *Revision Clinical Modification* (ICD-10) codes gleaned from Veterans’ EHRs.

#### MVP Baseline Survey

The MVP Baseline Survey gathers information pertaining to sociodemographics, military history, health status and habits, and medical history and health care usage. Within the “Medical History and Health Care Usage” section of the survey, Veterans are provided a list of health conditions and are asked to self-report whether they have been diagnosed with any of the health conditions. Two questions pertaining to TBI were extracted from the MVP Baseline Survey and utilized in our TBI phenotype: “Concussion or loss of consciousness” and “Traumatic brain injury” (hereafter labeled “BL-CC” and “BL-TBI,” respectively).

#### MVP Lifestyle Survey

The MVP Lifestyle Survey gathers information pertaining to psychosocial health and activities, men’s and women’s health, exercise and nutrition, well-being, and military and environmental experiences. Within the “Military and Environmental Experiences” section of the survey, Veterans are asked specifically about deployment-related experiences. If a Veteran indicates that they have been deployed and served in a combat or war zone, they are asked additional questions to assess for history of deployment-related TBI. These questions, based on the Brief Traumatic Brain Injury Screen^30^, were designed to assess mechanism of injury, signs or symptoms associated with TBI, and current symptoms. Specific questions were as follows: (1) “Did you have any injury(ies) during your deployment from any of the following?”, with response options including “fragment,” “bullet,” “vehicular,” “fall,” “blast,” “other,” and “none”; (2) “Did any injury received while you were deployed result in any of the following?”, with response options including “being dazed, confused or seeing stars,” “not remembering the injury,” “losing consciousness [knocked out] for less than a minute,” “losing consciousness for 1-20 minutes,” “losing consciousness for longer than 20 minutes,” “having any symptoms of concussion afterward (such as headache, dizziness, irritability, etc.),” “head injury,” and “none of the above”; and (3) “Are you currently experiencing any of the following problems that you think might be related to a possible head injury or concussion?”, with response options including “headache,” “dizziness,” “memory problems,” “balance problems,” “ringing in the ears,” “irritability,” “sleep problems,” and “other”. Responses to the second question pertaining to signs and symptoms associated with TBI (hereafter labeled “LS-Q39”) were extracted from the MVP Lifestyle Survey and utilized in our TBI phenotype.

#### ICD-9/ICD-10 Codes

ICD-9/10 codes pertaining to TBI were extracted from the EHR. TBI-specific ICD-9/10 codes were established using surveillance case definitions for TBI set forth by the Armed Forces Health Surveillance Branch of the Military Health System.^31^ The list of ICD-9 and ICD-10 codes utilized in this study is provided in Supplemental Tables 2 and 3, respectively.

### TBI Phenotyping

#### TBI Case Definition

To be considered a TBI case, there must have been evidence of TBI as determined by: (1) endorsement of BL-CC *or* (2) endorsement of BL-TBI *or* (3) endorsement of at least one sign/symptom associated with TBI on LS-Q39 *or*

(4) at least one inpatient or outpatient ICD-9/10 code for TBI. In other words, Veterans with evidence of *any one or more* of these criteria were classified as a TBI case.

#### TBI Control Definition

To be considered a TBI control, there must have been no evidence of TBI as determined by: (1) no endorsement of BL-CC *and* (2) no endorsement of BL-TBI *and* (3) no endorsement of any signs/symptoms associated with TBI on LS-Q39 *and* (4) at least one inpatient or outpatient ICD-9/10 diagnosis in the EHR but no ICD-9/10 codes for TBI at any point in the EHR. In other words, Veterans must have completed the MVP Baseline Survey and MVP Lifestyle Survey and not reported any event consistent with TBI on either survey. Additionally, there must have been evidence of VHA usage as determined by the presence of an inpatient or outpatient ICD-9/10 diagnosis, but no TBI ICD codes in the EHR.

### MVP Genotyping Procedures

The MVP Bioinformatics Core completed the data cleaning and processing for all genetics data. All genetic analyses for this study utilized MVP Release 4 data. MVP genotyping procedures have been described in detail elsewhere^29^; briefly, the “Affymetrix Axiom Biobank Array” was used for genotyping and standard quality control procedures were applied. Phasing occurred using Eagle v2.4 and imputation occurred through Minimac v4 using the Haplotype Reference Consortium panel^32,33^.

### Ancestry Assignment & Principal Components

For this study, the harmonized ancestry and race/ethnicity (HARE) method^34^ was used to inform genetic ancestry for GWAS; ancestry groups included EA, AA, and HA. KING^35^ was used to estimate relatedness (defined as kinship coefficient > 0.0884); related individuals were removed from analysis by prioritizing cases and removing controls. When related pairs were both cases (or both controls), one individual was removed at random. FlashPCA2^36^ was used to compute principal components (PCs) within each ancestry group.

### GWAS and Meta-Analysis

Separate GWASs were conducted for each ancestry group (EA, AA, and HA). Logistic regression analyses were performed to evaluate associations between genotype dosage data and TBI case-control status using PLINK^37^, including the first 10 PCs as covariates. The standard genome-wide significance threshold was used (*p* < 5 × 10^−8^). SNPs with MAF < 1% or an imputation information score < 0.6 were excluded from the results. Multi-ancestry meta-analysis was conducted in METAL^38^ using SNPs present in all datasets. To produce regional visualizations of GWS loci, LocusZoom 1.4^39^ was used. The 1000 Genomes phase 3 reference data (1KGPp3)^32^ was utilized to calculate LD, relying on ancestry-specific reference genotypes.

### Replication

GWS SNPs in the EA MVP cohort were compared to the largest available external TBI data repository—FinnGen (https://www.finngen.fi/en). Within FinnGen (Release 7), two TBI phenotypes were examined, one for “concussion” (15,787 cases, 184,565 controls) and one for “severe TBI” (4,927 cases, 304,227 controls).

### Functional Mapping and Annotation

Functional Mapping and Annotation (FUMA) software^40^ was used for functional mapping and annotation of GWAS results (default settings were used unless otherwise specified), with annotations derived from human genome assembly GRCh37 (hg19). To delineate independent genomic risk loci (defined by r^2^ > 0.6) and variants in LD with lead SNPs, the SNP2Gene module was applied using ancestry-appropriate reference genotypes. To map risk loci to protein-coding genes, a 10kb window size was used. eQTL mapping was completed for significant SNP-gene pairs (FDR < 0.05) using GTEx v8 brain tissue expression data.

### Gene-Based Results and Gene-Pathway and Tissue-Enrichment Analyses with MAGMA

Multi-Marker Analysis of GenoMic Annotation (MAGMA^41^) was used to conduct analyses pertaining to genes and tissue enrichment (i.e., gene-based, gene-pathway, and tissue-enrichment analyses). Regarding gene-based analyses, SNPs were mapped to 18,873 protein coding genes, and Bonferroni correction was used to establish the GWS threshold for these analyses (*p*=0.05/18,873=2.65×10^−6^). To determine whether specific biological pathways were associated with TBI, 15,485 curated gene sets and GO terms (from MsigDB) were used in a pathway analysis; Bonferroni correction was again applied (*p*=0.05/15,485=3.23×10^−6^). Finally, tissue-enrichment analyses were carried out using GTEx v8 RNA-seq and BrainSpan RNA-seq expression data.

### SNP-Based Heritability Estimation and Genetic Correlations

SNP-based heritability (SNP-*h*^*2*^) was computed using linkage disequilibrium score regression (LDSC)^42^ in the EA cohort. The 1KGPp3 EA reference sample was used to calculate input LD scores. Both observed scale and liability scale SNP-*h*^*2*^ are reported; liability scale SNP-*h*^*2*^ was computed using a population prevalence of 2% and 20%. Genetic correlations (r_g_) were also computed using LDSC.

### Univariate and Bivariate Gaussian Mixer Model (MiXeR) Analysis

MiXeR v1.3^43^ was used to estimate polygenicity and discoverability of causal variants for TBI. Polygenicity estimates reflect the number of loci needed to explain 90% of SNP-*h*^*2*^. Bivariate MiXeR^44^ was utilized to estimate phenotype specific and shared polygenicity. Akaike information criteria (AIC) values were used to evaluate model fit.

### Genomic Structural Equation Modeling (gSEM)

Genomic structural equation modeling (gSEM) was conducted using GWAS summary statistics in R^45^ (using the genomicSEM package^46^). LDSC^42^ was carried out in the EA cohort to estimate the genetic covariance matrix and the sampling covariance matrix using the 1KGPp3 EA reference for the following variables: TBI, general risk tolerance (GRT), major depressive disorder (MDD), attention-deficit/hyperactivity disorder (ADHD), alcohol dependence (ALCH), posttraumatic stress disorder (PTSD), reaction time (RT), verbal numeric reasoning (VNR), neuroticism (NEUR), Parkinson’s disease (PD), and Alzheimer’s disease (AD). Additionally, we evaluated the following brain morphometric variables: nucleus accumbens (NCA), amygdala (AM), brainstem (BS), caudate nucleus (CN), globus pallidus (GP), putamen (PU), thalamus (TH), hippocampal volume (HV), intracranial volume (ICV), full surface area (FSA), and full average thickness (FT). SNPs within the major histocompatibility region were removed in the calculation of genetic covariance.

Exploratory factor analysis (EFA) was used to assess the number of latent factors among variables pertaining to TBI, risk-taking behaviors, and psychiatric disorders. Using the R *factanal* function, EFA was carried out on the odd chromosomes and then confirmatory factor analyses (CFAs) were carried out on the covariance matrix from the even chromosomes. A factor loading threshold of 0.35 or greater was required from the EFA for variables to be assigned to factors in the CFA. The weighted least squares estimator was used to fit the data. Model fit was evaluated using the chi-square test *p*-value, comparative fit index (CFI), AIC, and standardized root mean square residual (SRMR).

### Phenome-wide Association Study (PheWAS)

A PheWAS for GWS SNPs from the TBI GWAS was conducted using GWAS Atlas (https://atlas.ctglab.nl/PheWAS)^47^. SNP rs-ID’s were entered and traits were sorted by “Domain and P-value” and the max P-value was set to 0.05. Data were downloaded from the GWAS Atlas website and plotted in R. Bonferroni correction was applied (*p*=0.05/3,302=1.51×10^−5^).

## Results

### SNP-Based Results

The multi-ancestry meta-analysis GWAS of TBI (case-control) was conducted using a sample of 304,485 Veterans (111,494 cases; 192,991 controls). A Manhattan plot of the complete results is shown in Figure 1. We identified 15 genome-wide significant (GWS) independent loci (Table 2). Regional annotation plots of GWS loci are presented in Supplemental Figure 1. The leading SNP of the most significant locus (rs1940701, locus 13) was located in *NCAM1* on chromosome 11. The leading SNP of the second most significant locus (rs729053, locus 17) was in *TCF4* on chromosome 18. The leading SNP of the third most significant locus (rs429358, locus 19) was in *APOE* on chromosome 19. Notably, rs429358 is one of the two polymorphisms (alongside rs7412) that determines the *APOE* haplotype^48^.

**Table 2.**
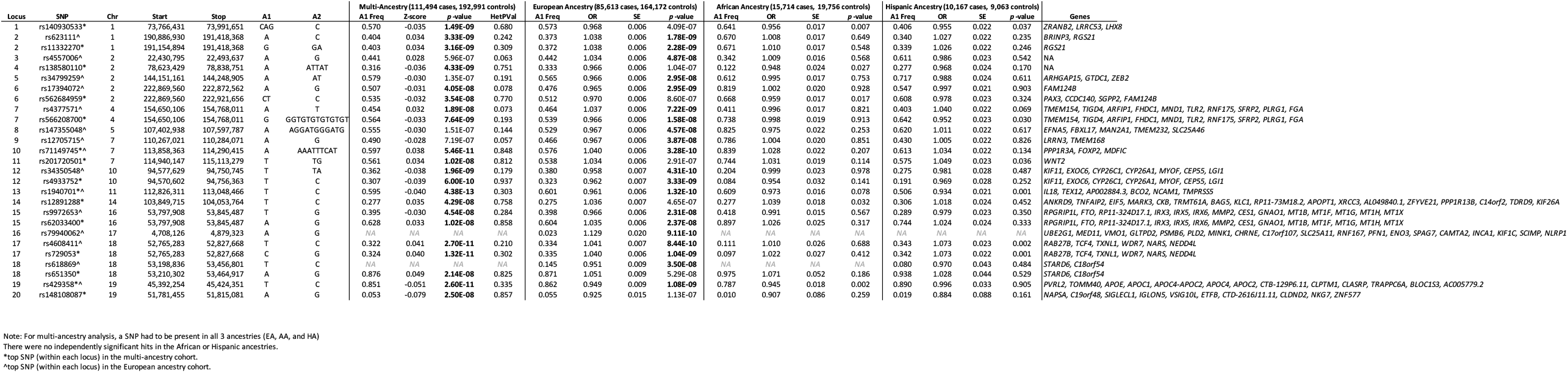
Genome-wide significant loci associated with TBI (case-control) for multi-ancestry, EA, AA, and HA cohorts.

**Figure 1.**
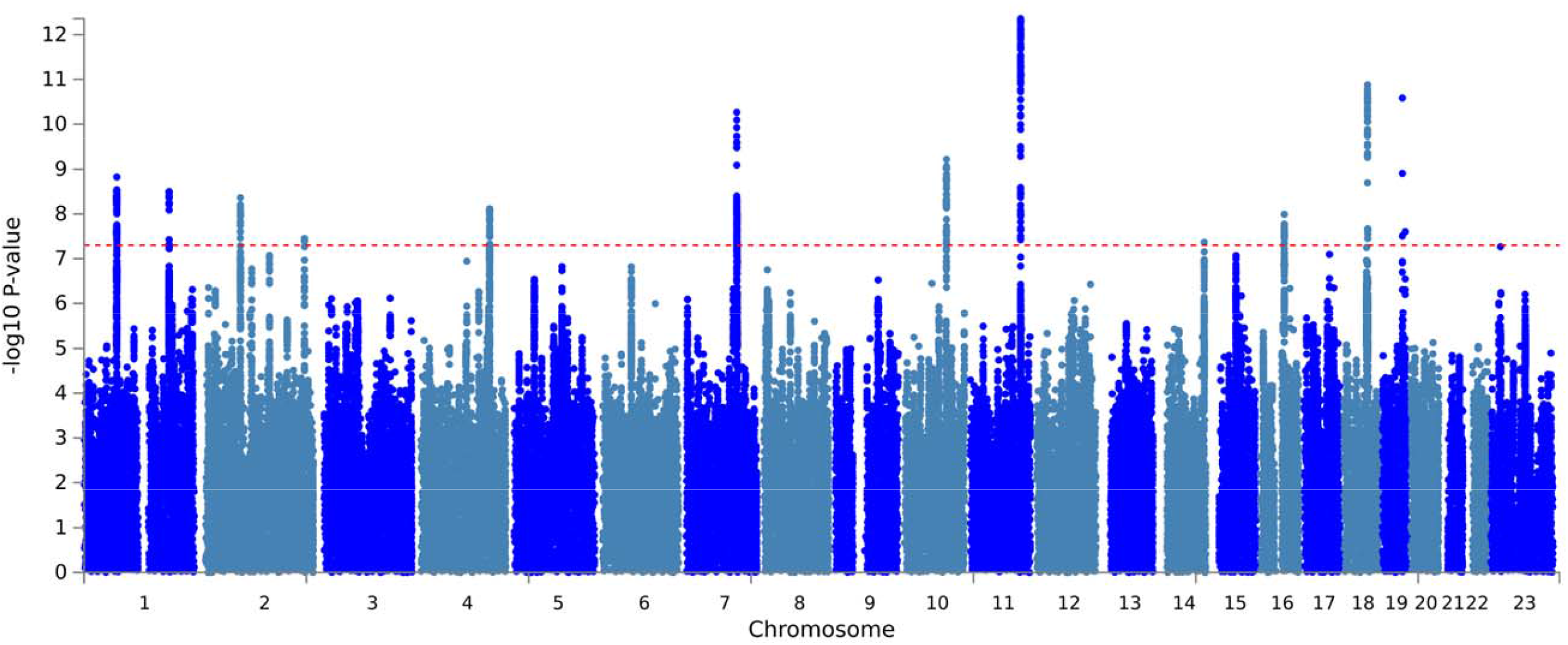
Manhattan plot of TBI risk for the **multi-ancestry cohort**. The GWAS was conducted using logistic regression, including first 10 principal components of ancestry as covariates. The x-axis displays the chromosome position, and the y-axis shows the GWAS *p* value on a -log_10_ scale. The red dashed line reflects genome-wide significance at *p* < 5 × 10^−8^.

When examining Veterans of EA only (N=249,785; 85,613 cases, 164,172 controls), we similarly identified 15 GWS independent loci. As in the multi-ancestry analysis, the most significant locus was in *NCAM1*, with the same leading variant (rs1940701) identified. Of the 15 loci, 4 were distinct from the multi-ancestry analysis (Table 2, Supplemental Figure 2), but all had been suggestive of significance (*p*<1×10^−6^) in the multi-ancestry meta-analysis. The leading SNP of the most significant of these 4 distinct loci was in *ARHGAP15* on chromosome 2 (rs34799259, locus 5). Supplemental Table 4 shows functional mapping and annotation of the TBI GWAS results in the EA cohort. No GWS loci were identified in Veterans of AA (N=35,470; 15,714 cases, 19,756 controls) or HA (N=19,230; 10,167 cases, 9,063 controls; Table 2, Supplemental Figures 3-4).

### Replication

We performed replication analysis of the 15 EA GWS loci in the FinnGen concussion phenotype GWAS (15,787 cases, 184,565 controls; Supplemental Table 5). rs71149745 (locus 10, *FOXP2*) remained significant after adjustment for multiple comparisons. Three additional loci were at least nominally significant (at *p*<.05): (1) rs1940701, nearest to *NCAM1* (locus 13); (2) rs12705715 (locus 9); and (3) rs1320139 (a proxy variant for rs34799259, locus 5), nearest to *ARHGAP15*. A sign test was performed to determine whether the direction of effects corresponded across studies at a rate beyond chance (50%). The direction of effects matched for 13 of the 15 variants, a significantly greater rate of correspondence than chance (*p*=.001). When comparing the MVP EA case-control TBI phenotype to the FinnGen severe TBI phenotype (4,927 cases, 304,227 controls; Supplemental Table 4), two of the 15 possible SNPs nominally replicated at *p*<.05: (1) rs71149745, nearest to *FOXP2* (locus 10) and (2) rs4933752 (a proxy variant for rs34350548, locus 12), nearest to *EXOC6*.

### Gene-Based Analyses

In the genome-wide gene-based association study in the multi-ancestry cohort, 14 genes were determined to be gene-wide significant following Bonferroni correction for multiple comparisons (Table 3, Supplemental Table 6, Supplemental Figure 5a). The top gene identified was *NCAM1* (*p*=4.10×10^−12^), which was already named in positional mapping of the SNP-based analysis results, followed by *APOE* (*p*=1.30×10^−9^), *FTO* (*p*=6.94×10^−9^), and *FOXP2* (*p*=2.68×10^−8^), also all positionally mapped to significant loci from the SNP-based analysis. In contrast, five of the significant genes from this analysis (*MSRA, FAM120A, NAV3, RBFOX1*, and *DCC*) were not identified based on positional mapping of loci from the SNP-based analysis. *MSRA* is located on the chromosome 8 inversion region^49^ where LD patterns are such that the association may be tagging the inversion itself rather than *MSRA* specifically.

**Table 3.** Gene-based loci analysis (Bonferroni significant) in the multi-ancestry and European ancestry cohorts.

| SYMBOL | CHR | START | STOP | Multi-Ancestry |  |  |  | European Ancestry |  |  |  |
| --- | --- | --- | --- | --- | --- | --- | --- | --- | --- | --- | --- |
|  |  |  |  | NSNPS | NPARAM | ZSTAT | P | NSNPS | NPARAM | ZSTAT | P |
| *SFRP2* *^ | 4 | 154701744 | 154710272 | 16 | 4 | 4.761 | 9.65E-07 | 18 | 5 | 4.579 | 2.34E-06 |
| *RNF175* *^ | 4 | 154631277 | 154681387 | 208 | 12 | 4.951 | 3.70E-07 | 216 | 13 | 5.040 | 2.33E-07 |
| *TSSK1B*^ | 5 | 112768251 | 112770728 | - | - | - | - | 15 | 4 | 4.605 | 2.06E-06 |
| *FBXL17*^ | 5 | 107194736 | 107717799 | - | - | - | - | 1390 | 38 | 4.703 | 1.28E-06 |
| *FOXP2* *^ | 7 | 113726382 | 114333827 | 889 | 34 | 5.439 | 2.68E-08 | 1122 | 49 | 5.794 | 3.43E-09 |
| *MSRA* * | 8 | 9911778 | 10286401 | 1471 | 59 | 4.707 | 1.25E-06 | - | - | - | - |
| *FAM120A* *^ | 9 | 96214004 | 96328397 | 300 | 15 | 4.704 | 1.28E-06 | 354 | 19 | 5.054 | 2.17E-07 |
| *PHF2*^ | 9 | 96338689 | 96441869 | - | - | - | - | 491 | 26 | 5.270 | 6.84E-08 |
| *EXOC6* *^ | 10 | 94590935 | 94819250 | 525 | 19 | 4.775 | 8.97E-07 | 652 | 26 | 4.663 | 1.56E-06 |
| *NCAM1* *^ | 11 | 112831997 | 113149158 | 848 | 38 | 6.835 | 4.10E-12 | 999 | 57 | 6.195 | 2.92E-10 |
| *NAV3* *^ | 12 | 78224685 | 78606790 | 1251 | 63 | 5.152 | 1.29E-07 | 1463 | 78 | 5.097 | 1.73E-07 |
| *TRMT61A* * | 14 | 103995521 | 104003410 | 18 | 3 | 4.488 | 3.59E-06 | - | - | - | - |
| *RCN2*^ | 15 | 77223960 | 77242601 | - | - | - | - | 34 | 12 | 4.499 | 3.42E-06 |
| *TCF12*^ | 15 | 57210821 | 57591479 | - | - | - | - | 1009 | 18 | 4.515 | 3.16E-06 |
| *FTO* *^ | 16 | 53737875 | 54155853 | 1303 | 93 | 5.675 | 6.94E-09 | 1503 | 116 | 5.617 | 9.72E-09 |
| *RBFOX1* * | 16 | 6069095 | 7763340 | 9543 | 385 | 4.937 | 3.98E-07 | - | - | - | - |
| *AKAP10*^ | 17 | 19807615 | 19881656 | - | - | - | - | 123 | 8 | 4.674 | 1.48E-06 |
| *DCC* *^ | 18 | 49866542 | 51057784 | 4415 | 85 | 4.993 | 2.97E-07 | 5169 | 101 | 4.926 | 4.19E-07 |
| *APOE* *^ | 19 | 45409011 | 45412650 | 5 | 2 | 5.955 | 1.30E-09 | 6 | 3 | 5.682 | 6.66E-09 |
| *APOC1* * | 19 | 45417504 | 45422606 | 11 | 3 | 4.621 | 1.91E-06 | - | - | - | - |
| *FAM46D*^ | X | 79591003 | 79700810 | - | - | - | - | 58 | 3 | 5.055 | 2.15E-07 |
| *BRWD3*^ | X | 79926353 | 80065187 | - | - | - | - | 180 | 10 | 4.540 | 2.81E-06 |

Notes: N=304,485 for the multi-ancestry cohort; N=249,785 for the European ancestry cohort.
\*top gene in the multi-ancestry cohort.
^top gene in the European ancestry cohort.

We also examined GWS genes in Veterans of EA, AA, and HA separately. In the EA cohort, 18 genes were determined to be GWS following Bonferroni correction (Table 3, Supplemental Table 7, Supplemental Figure 5b). Eight of these genes (*AKAP10, BRWD3, FAM46D, FBXL17, PHF2, RCN2, TCF12*, and *TSSK1B*) were distinct from the multi-ancestry gene-based analysis. Among these, the leading association was with *PHF2*. In the AA and HA cohorts, there were no gene-wide significant genes identified (data not shown).

### Gene Tissue Expression

We used FUMA software^40^ to evaluate gene-tissue expression in the multi-ancestry cohort. Results showed that among 30 general tissues, the brain was the only significantly enriched tissue, particularly in the frontal cortex, anterior cingulate cortex, and nucleus accumbens (tissues aggregated: Figure 2a, Supplemental Table 8; stratified by tissue subtype: Figure 2b, Supplemental Table 9). Additionally, gene-set analyses were conducted to understand potential gene pathways of importance; however, results did not identify any significant gene sets in the multi-ancestry cohort (Supplemental Table 10). When evaluating Veterans of EA separately, there were no significant gene-tissue expression results and no significant gene sets (Supplemental Tables 11-12; Supplemental Figures 6a-6b).

**Figure 2a.**
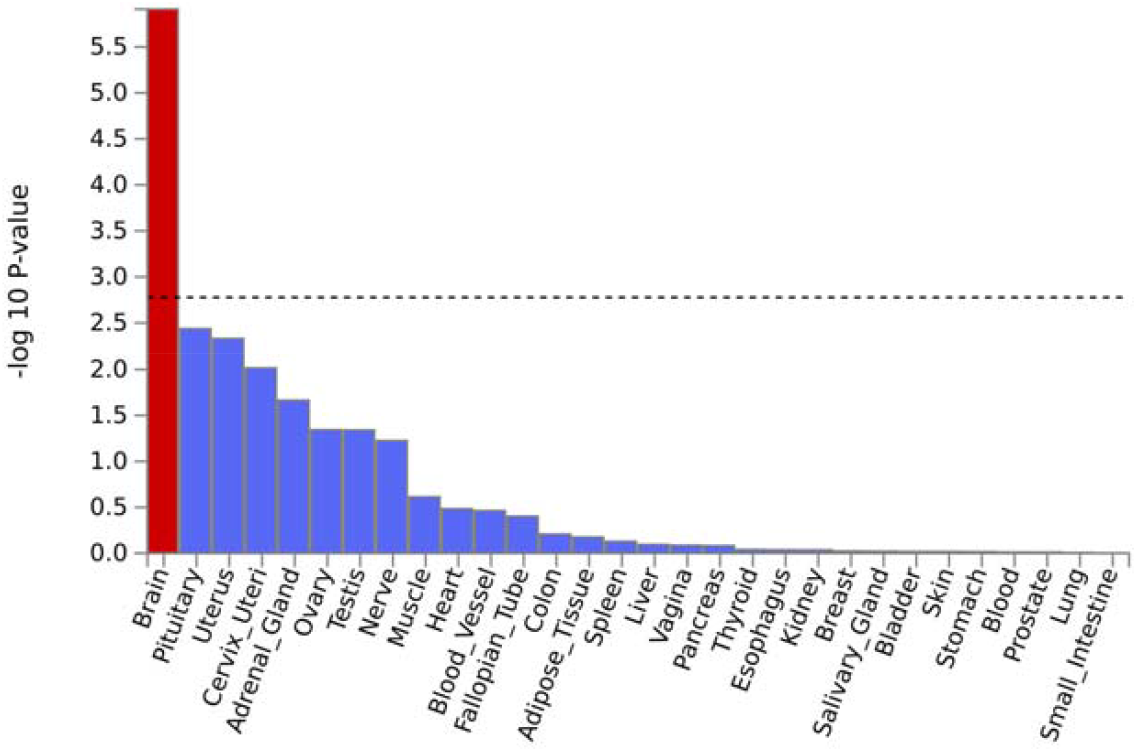
Gene tissue expression (tissues aggregated) in the **multi-ancestry** cohort.

**Figure 2b.**
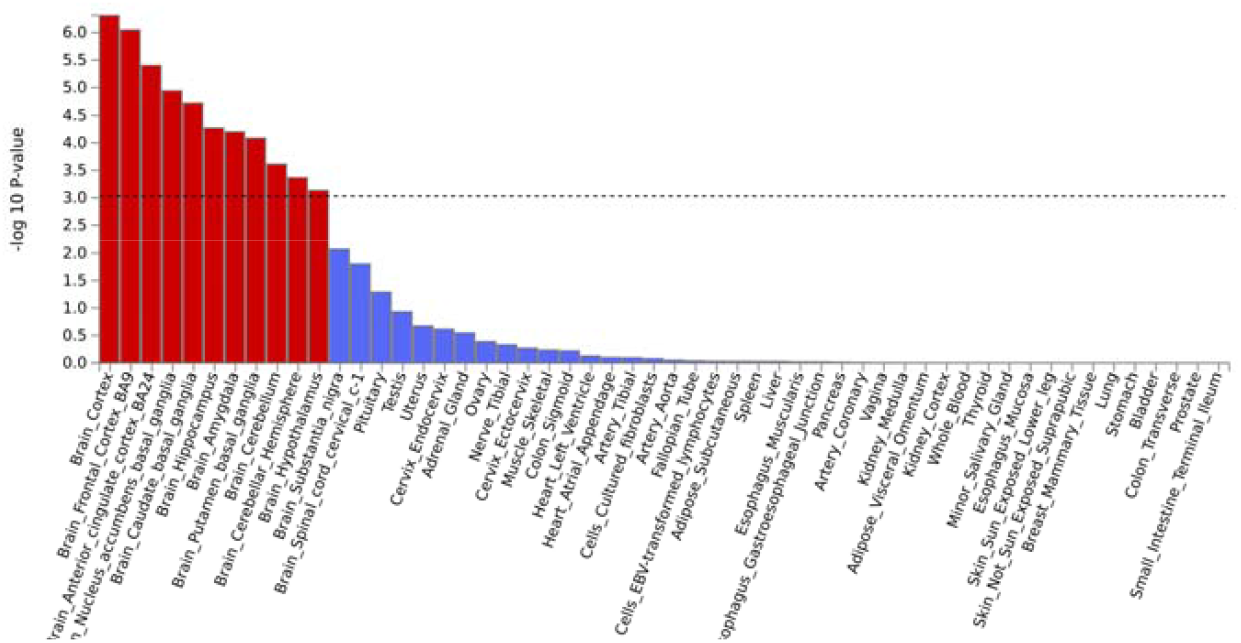
Gene tissue expression (stratified by tissue subtype) in the **multi-ancestry** cohort.

### SNP-Based Heritability

Linkage disequilibrium score regression (LDSC)^42^ was used to estimate SNP-based heritability (SNP-*h*^*2*^) of the case-control TBI phenotype in the EA cohort. Observed SNP-*h*^*2*^ was estimated to be 0.060 (SE=0.004, *p*=7.83×10^−66^). On the liability scale, at 2% population prevalence, SNP-*h*^*2*^ was 0.044 (SE=0.003); at 20% population prevalence, SNP-*h*^*2*^ was 0.087 (SE=0.005). To obtain further insights into the genetic architecture of TBI, we used univariate MiXeR analysis to decompose heritability into discoverability and polygenicity (number of influential variants) subcomponents. An estimated 10,470 (SE=848) influential variants explain 90% of the SNP-*h*^*2*^ and the discoverability was 9.2×10^−6^ (SE=6.7×10^−7^). The 15 variants identified in the EA GWAS explained an estimated 0.2% of the heritability of TBI.

### Genetic Overlap

We examined genetic correlations (*r*_*g*_) between the MVP TBI variable and a variety of *a priori* selected health domains including risk-taking behaviors, psychiatric disorders, neurocognitive variables, and brain morphometrics in the EA cohort (Figure 3, Supplemental Tables 13-16, Supplemental Figures 7-8). The strongest genetic correlation was with PTSD (*r*_*g*_=0.69, SE=0.04, *p*=2.5×10^−75^). Broadly, genetic correlations with TBI were moderate-to-large for risk-taking behaviors and psychiatric disorders. TBI was also negatively correlated with verbal numeric reasoning (*r*_*g*_=-0.35, SE=0.03, *p*=4.2×10^−42^). Genetic correlations with reaction time and neurogenerative disorders (i.e., AD and PD) were not significantly different from 0 after adjustment for multiple comparisons. With regard to brain morphometrics, TBI was weakly genetically correlated to ICV (*r*_*g*_=-0.12, SE=0.04, *p*=0.002), but all other brain morphometric correlations were not significantly different from 0.

**Figure 3.**
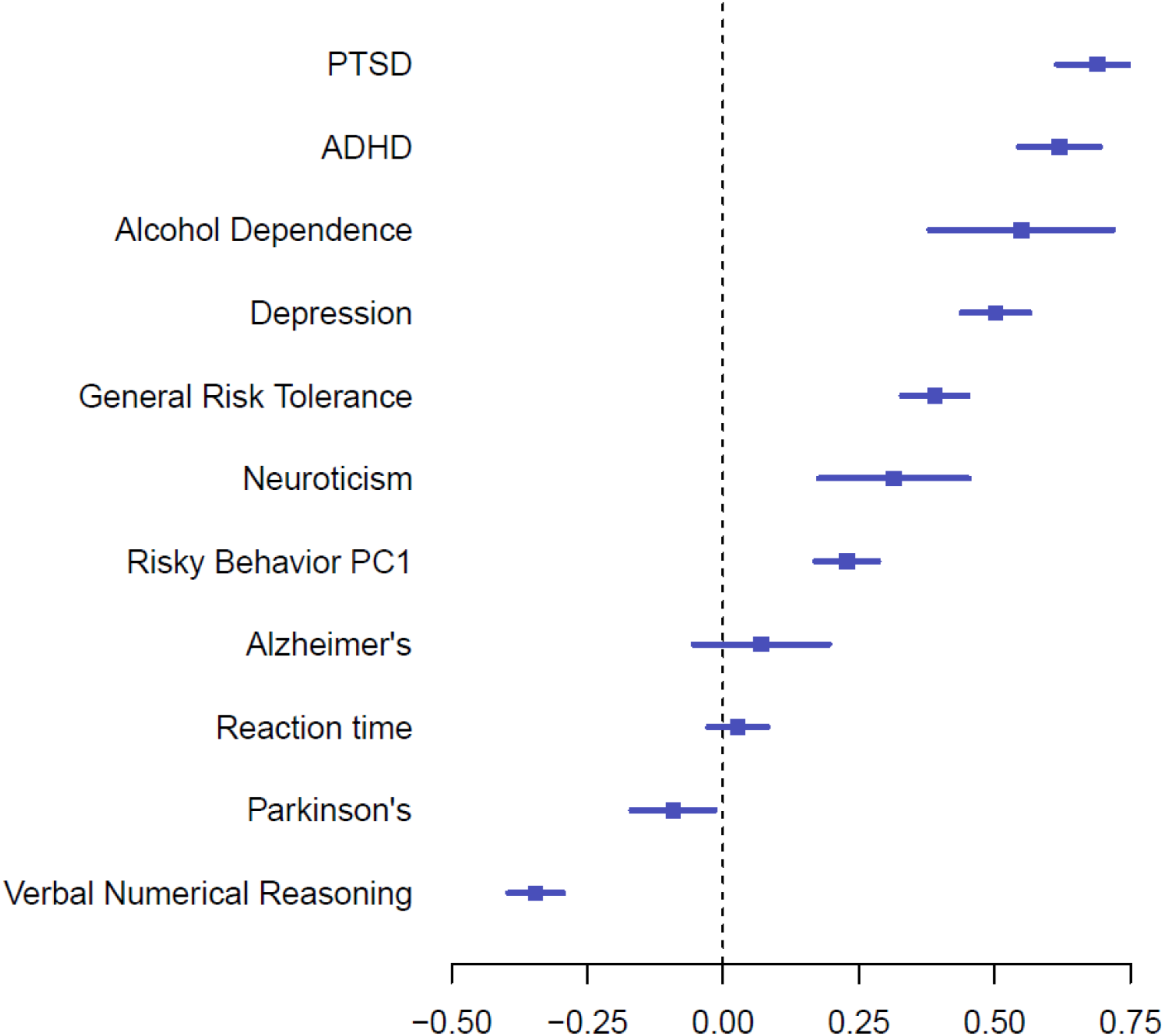
Genetic correlation between TBI and other phenotypes. The x-axis denotes the magnitude of genetic correlation. The blue squares represent the genetic correlation between TBI and each given phenotype, with lines representing 95% confidence intervals.

We used univariate and bivariate MiXeR to perform a more detailed evaluation of the genetic overlap between TBI and these phenotypes. Univariate analyses suggest that the genetic architecture of the TBI phenotype (i.e., the polygenicity and discoverability of risk variants) was similar to polygenic psychiatric disorders (Supplemental Tables 17-18). In contrast, neurodegenerative disorders such as AD were characterized by relatively low polygenicity but high discoverability. In bivariate analyses, for five phenotypes investigated (AD, neuroticism, risky behaviors, reaction time (RT), and verbal numeric reasoning), there was evidence of polygenic overlap with TBI beyond what was explained by a genetic correlation model (best vs min AIC > 0). Notably, while the genetic correlations of TBI with AD and RT were not significantly different from 0, MiXeR reported that 60% (number of shared influential variants=57, SE=23) of the influential variation in AD and 94% (number of shared influential variants=8,902, SE=502) of the influential variation in RT were shared with TBI (Supplemental Figures 9-10). MiXeR also reported the existence of genetic variation specific to PD (best vs max AIC = 0.57), where the *r*_*g*_ estimated among only shared influential variants was -0.83 (SE=0.14).

### Genomic Structural Equation Modeling (gSEM)

Given the strong genetic correlations between TBI and risk-taking behaviors and psychiatric disorders, genomic structural equation models^46^ were conducted within the EA cohort to better understand whether TBI is more genetically associated with a latent factor representing risk-taking behaviors versus a latent factor representing psychiatric disorders. Pairwise genetic correlations were calculated between all phenotypes to produce a genetic correlation matrix (Supplemental Figure 7-8) for use in factor analysis. In genomic confirmatory factor analysis, the TBI variable loaded onto its own factor while the psychiatric disorders loaded onto one factor and the risk-taking behaviors loaded onto another factor (CFI=0.908) (Figure 4). The correlation between the TBI and psychiatric disorders factors was 0.55 (SE=0.04), and the correlation between the TBI and risk-taking behaviors factors was 0.73 (SE=0.06), indicating that TBI risk has a stronger genetic correlation with risk-taking behaviors than with the psychiatric disorders. When examining genetic associations between TBI and individual psychiatric disorders, PTSD, alcohol dependence, and ADHD showed the strongest associations with TBI (Supplemental Figure 7). Given this observation, a final set of analyses was run to examine the genetic correlation between these individual disorders and TBI, controlling for the genetic association with risk-taking. These multiple regression models are presented in Figure 5; the model with PTSD continued to showed the strongest relationship with TBI even after accounting for risk-taking.

**Figure 4.**
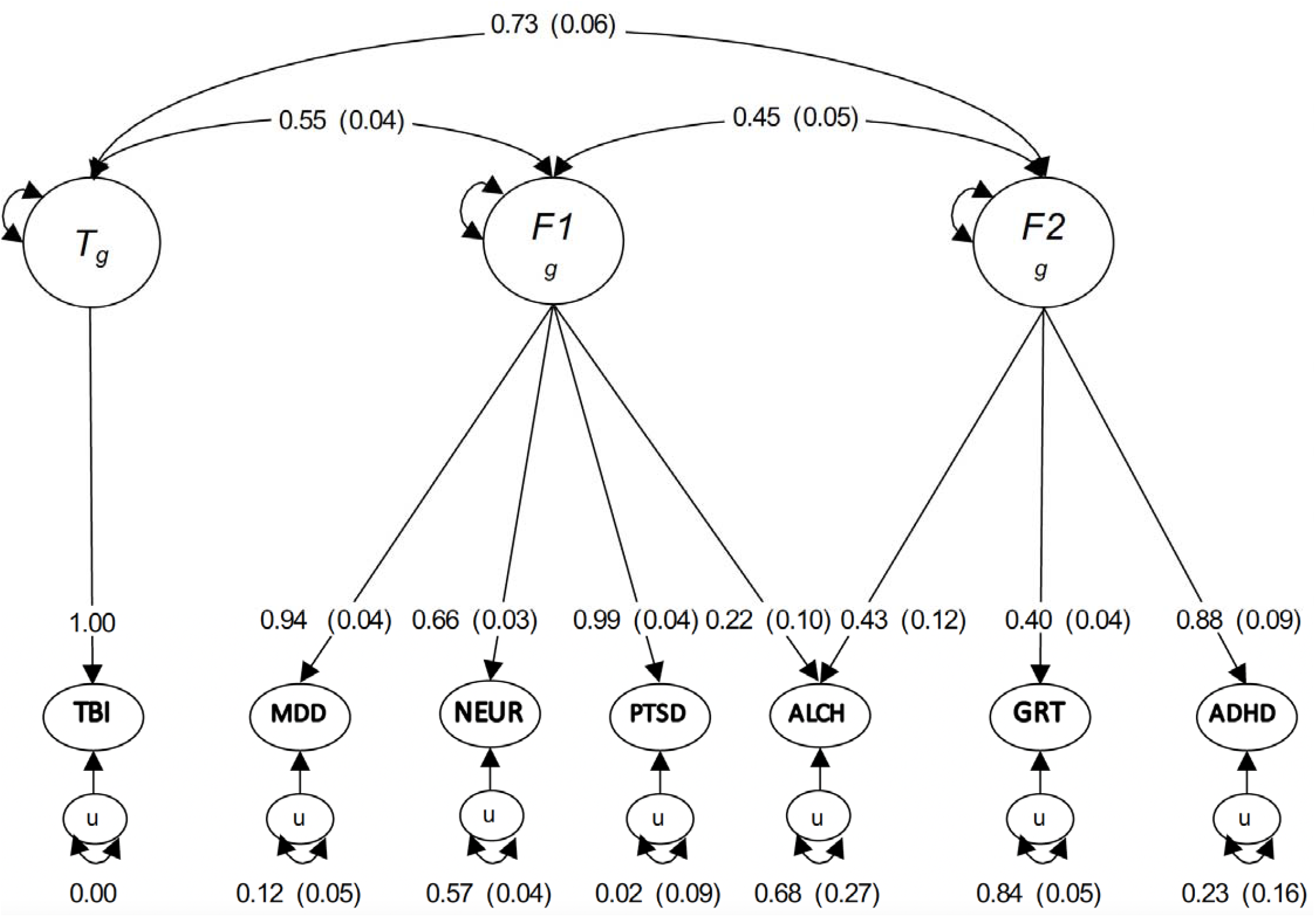
Genomic structural equation modeling (gSEM) results. Correlated factors model with standardized estimates and standard error estimates in parentheses. Genetic components of disorders are in circles and latent factors are represented by ovals. Single-headed arrows represent regression relationships and parameter estimates that can be squared to estimate the proportion of variance accounted for by components. Double-headed arrows represent correlations. Residual variance represented by double-headed arrow connecting the variable to itself. Each phenotype loads onto at least one factor. The TBI risk variable is loaded onto its own factor (*T*_*g*_) while the psychiatric disorders loaded onto one factor (*F1*_*g*_) and the risk-taking behaviors loaded onto another factor (*F2*_*g*_); CFI=0.908. *<u>Abbreviations</u>*: TBI = traumatic brain injury; MDD = major depressive disorder; NEUR = neuroticism; PTSD = posttraumatic stress disorder; ALCH = alcohol dependence; GRT = general risk tolerance; ADHD = attention-deficit/hyperactivity disorder.

**Figure 5.**
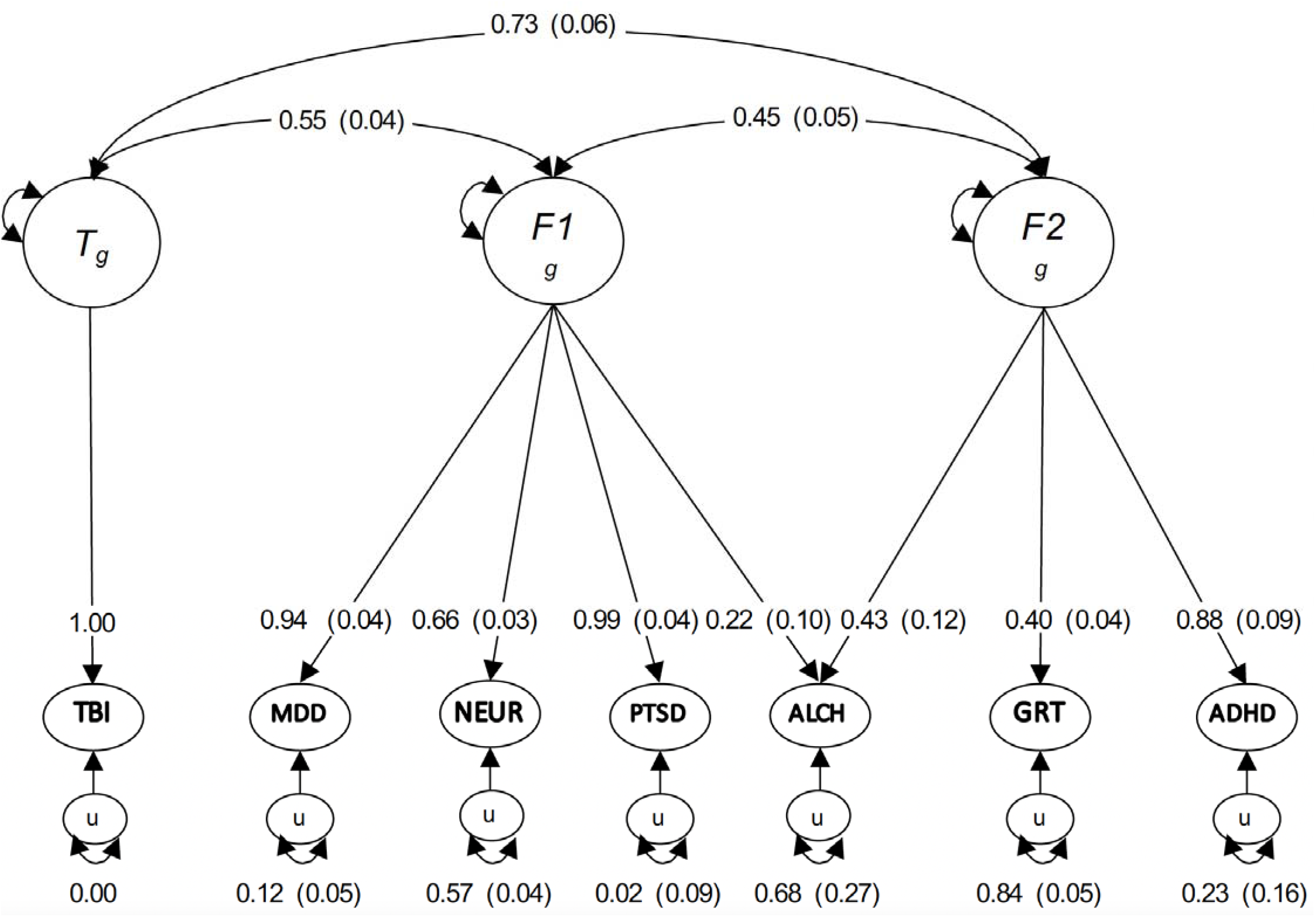
Multiple regression results. Genetic components of disorders are in circles. Single-headed arrows represent regression relationships and parameter estimates that can be squared to estimate the proportion of variance accounted for. Double-headed arrows represent correlations. Residual variance represented by double-headed arrow connecting the variable to itself. The models reflect the genetic association between TBI and (A) PTSD, (B) alcohol dependence, and (C) ADHD while controlling for general risk taking. *<u>Abbreviations</u>*: PTSD = posttraumatic stress disorder; GRT = general risk tolerance; TBI = traumatic brain injury; ALCH = alcohol dependence; ADHD = attention-deficit/hyperactivity disorder.

### PheWAS

Our PheWAS examined associations of leading variants in TBI risk loci with phenotypes across 28 domains (Supplemental Figure 11, Supplemental Table 19). The most common associations were with phenotypes in psychiatric (11 loci), activities (9 loci), metabolic (8 loci), and environment (7 loci) domains (Supplemental Figure 12). Considering the degree of pleiotropy of the loci, the (median) average locus was associated with phenotypes across three different domains. Loci 15 (*FTO*) and 19 (*APOE*) had significant associations with the largest number of domains (16 and 15 domains, respectively). In contrast, six loci were only associated with phenotypes within a single domain (e.g., locus 7, which was only associated with a phenotype in the skeletal domain [osteoarthritis]).

## Discussion

Examining the role of genetics in TBI risk and recovery may offer novel mechanistic insights and lead to important discoveries that can inform preventive efforts and innovative treatments for patients who have sustained TBIs. In this study, we present the largest GWAS to date for TBI using data from the VA Million Veteran Program. Our results showed GWS variants associated with a TBI case-control phenotype in a multi-ancestry cohort of Veterans. These associations were primarily driven by the EA cohort, which comprised the majority of the overall sample (82%), and findings replicated in the FinnGen cohort (notably, the direction of effects matched for 13 of the 15 variants). We also independently evaluated Veterans of African and Hispanic ancestry, but found no independent GWS loci, likely owing to reduced sample size and therefore lower power in those groups.

In the multi-ancestry cohort, our SNP-based results revealed 15 GWS loci, and our gene-based results revealed 14 gene-wide significant genes, many of which have presumed roles in psychiatric and neurologic functioning. The top GWS SNPs within the multi-ancestry sample were in and around *NCAM1*, followed by *APOE, FTO*, and *FOXP2*. Notably, many of these genes have emerged as significant genes in prior studies of neuropsychiatric and neurodegenerative conditions. For example, NCAM1 (neural cell adhesion molecule 1) is a glycoprotein expressed on the surface of neurons, glia, and skeletal muscle, and has been shown to have a role in several brain-related biological processes such as neuronal development and maintenance as well as synaptic plasticity^50-52^. The *NCAM1* gene has been associated with cognitive functioning, particularly learning and memory, and has been implicated in a range of neuropsychiatric and neurodegenerative disorders including schizophrenia, mood and anxiety disorders, and AD^51^.

Another notable gene to emerge as significant was *APOE*. APOE is a protein expressed in peripheral tissues and the central nervous system and its primary function is facilitating the transportation and distribution of lipids throughout the body. APOE plays a role in neuronal maintenance, neural transmission, neuroinflammation, and synaptic plasticity within the brain and has been associated with response and repair processes following neurotrauma^53,54^. The *APOE* ε4 allele is a major risk factor for AD^55-57^, and other research has shown associations between *APOE* and medical conditions such as vascular dementia, diabetes mellitus, and cardiovascular disease^58,59^, as well as with cognitive functioning^60^. Furthermore, our findings of an association between *APOE* and TBI align with previous candidate gene studies in TBI^18,19,61-64^ and may help to explain the heterogeneous neurocognitive outcomes observed chronically following TBI. However, it is important to note that our association was observed with a single *APOE* SNP (and not the *APOE* haplotype which defines the *APOE* ε4 allele, which was not examined), and the effect of the haplotype may be different. Follow-up studies by our group are planned to more comprehensively address the role of the *APOE* haplotype in the context of TBI.

As for *FTO* (Fat and Obesity-Associated), less is known about the physiological function of this gene in the brain; however, *FTO* has been found to have important actions in metabolic and cardiovascular systems and may be associated with body mass index, obesity risk, and type 2 diabetes^65,66^. *FTO* gene variants have also been causally associated with risk of AD^67^ and have been reported to promote phosphorylation of tau by activating the mTor pathway^68^, which has been implicated in AD pathophysiology. Finally, FOXP2 (forkhead box P2) is a protein that is active in several tissues including the brain, and was initially implicated in speech and language development^69,70^. *FOXP2* has also been shown to play a role in other biological processes including brain development, cell differentiation and proliferation, and neurodegeneration^69,70^, and has previously been implicated as a genetic variant associated with ADHD, risk-taking behaviors, and psychiatric disorders such as PTSD^70,71^. Taken together, these observations offer critical insights into the genes that are associated with TBI. Although the functional significance of these genes within the context of TBI will still need to be determined, these initial findings are promising and offer targets for future research.

To better understand the role of TBI-associated genes in higher-order systems, we examined gene tissue expression. As expected, our results showed that the brain was the only significantly enriched region, with TBI-associated genes specifically implicated in the cortex, frontal cortex, anterior cingulate cortex (ACC), nucleus accumbens, caudate, hippocampus, amygdala, putamen, cerebellum, cerebellar hemisphere, and hypothalamus. These findings converge with a recent GWAS examining risk tolerance that similarly showed significant gene expression in brain regions such as the frontal cortex, ACC, and cortex^72^. Of note, the ACC is well-known to have connections with both the limbic system and the prefrontal cortex and has been implicated in decision-making and impulse control^73,74^, underscoring the biological link between TBI and risk-taking.

With regard to SNP-based heritability of the TBI phenotype, we found an observed heritability of 6%, which is generally consistent with other heritability statistics reported for risk-taking behaviors (e.g., general risk tolerance: SNP-*h*^*2*^=0.05^72^), psychopathology (e.g., PTSD [MVP]: SNP-*h*^*2*^=0.05-0.07^71^), and neurocognition (e.g., AD, SNP-*h*^*2*^=0.07^75^). Notably, there are no adequately powered studies reporting SNP-*h*^*2*^ for TBI for comparison. Future studies are thus needed to further explore the heritability of TBI and to specifically tease apart heritability associated with TBI acquisition versus TBI sequelae.

Finally, we examined genetic correlations and our LDSC results suggest that there is significant genetic overlap between the TBI case-control phenotype and risk-taking behaviors and psychopathology. In contrast, there was weak genetic correlation between the TBI case-control phenotype and cognition and brain morphometrics, with the exception of verbal numeric reasoning. gSEM analyses further confirmed underlying genetic associations between TBI and both risk-taking behaviors and psychiatric disorders, though the association was stronger between TBI and risk-taking. Still, when evaluating individual traits/conditions, PTSD had the strongest genetic overlap with TBI, followed by ADHD and alcohol use when controlling for general risk tolerance, providing evidence of shared genetic etiology. Finally, MiXeR results showed that the genetic architecture of TBI was similar to polygenic psychiatric disorders and risk-taking traits. In contrast, neurodegenerative disorders such as AD showed much less polygenicity but had a high degree of shared variance with TBI. The discrepancies between the architectures of TBI and neurodegenerative disorders suggest that results need to be interpreted with caution; however, they are at least indicative of shared genetics.

This is the first large-scale GWAS of TBI in the VHA and findings significantly advance understanding of the genetic basis of TBI. Nevertheless, there are caveats to consider when interpreting our findings. To maximize sample size and ensure sufficient power, we defined a TBI case as any positive indication of TBI on either the MVP Baseline Survey or MVP Lifestyle Survey, or at least one inpatient or outpatient ICD code for TBI. In theory, this approach allowed us to capture lifetime history of TBI. However, the diverse methods of data collection, as well as the lack of precise information on the timing between injury event and reporting or documentation of the event, may have impacted results. Of note, the MVP Baseline Survey specifically assessed for lifetime history of concussion/TBI whereas the MVP Lifestyle Survey evaluated deployment-related TBI. Furthermore, the ICD codes were limited to the data available within the Veterans’ VA EHRs, which may omit historical TBIs (i.e., TBI sustained prior to military service/enrollment in VA). Given the data, our TBI phenotype specifically focused on whether or not Veterans had a history of TBI, though it is possible that our phenotype also captured aspects of TBI recovery. Future MVP studies are planned to further understand the genetics associated with TBI-associated *sequelae* (e.g., “post-concussive syndrome” or “persistent post-concussive symptoms”) and other aspects of TBI *recovery*, as it is likely that different genes may be associated with TBI acquisition/susceptibility vs. TBI outcome. Moreover, given that we did not evaluate or consider TBI severity in our analyses, future research may also consider evaluating TBI or TBI-associated variables on a continuum—for example, examining injury severity or injury characteristics such as LOC or PTA.

Final study caveats relate to sample characteristics. Notably, the MVP cohort is predominately male; given our findings and the strong associations observed with risk-taking behaviors and psychiatric traits, it will be important for future research to determine whether genetic associations differ as a function of biological sex. The MVP cohort may be unique in other ways (e.g., high frequency of medical comorbidities in this population) and possibly distinct from other major biobanks; thus, it will be necessary for other biobanks to also examine the genetics associated with TBI.

## Conclusions

Learning more about the genetics associated with TBI risk and recovery is necessary for developing a better understanding of the pathophysiology of this injury and could aid in developing novel therapeutics. The results of this first large-scale GWAS examining TBI in military Veterans identified 15 loci in the multi-ancestry cohort, including genes previously known to be relevant to TBI biology that will be important to investigate in future studies. Findings also showed that TBI is a heritable trait with comparable genetic architecture and high genetic correlation with psychiatric and risk-taking traits. Results set the stage for future TBI GWAS studies within MVP that focus on diversity and chronicity of symptom sequelae as well as severity of injury.

## Supporting information

STROBE

## Data Availability

Final data sets underlying this study cannot be shared outside the VA, except as required under FOIA, per VA policy, due to the sensitive nature of medical health records of Veterans. However, upon request through the formal mechanisms in place and pending approval from the VHA Office of Research Oversight (ORO), a de-identified, anonymized dataset underlying this study can be created and shared once the study investigators have had sufficient time to analyze and publish research findings.

## Acknowledgements

The authors sincerely thank the Veterans who volunteered to participate in the Million Veteran Program. This research is based on data from the Million Veteran Program (Project MVP026), Office of Research and Development, Veterans Health Administration. This publication does not represent the views of the Department of Veteran Affairs or the United States Government.

## Author Contributions

Overall study coordination: VCM, CMN. Study concept and design: VCM, CMN, AXM, LDW. Data curation: CCC. Phenotype analysis: CCC (lead), VCM, LDW. Statistical analysis: AXM (lead), MG, EK. Data interpretation: AXM, MG, CMN, MBS, MSP, RLH, MWL, VCM. Writing: VCM (lead), AXM, CMN. All authors provided edits, feedback, and approved the final version of the paper for submission.

## References

1. Helmick KM, Spells CA, Malik SZ, Davies CA, Marion DW, Hinds SR. Traumatic brain injury in the US military: Epidemiology and key clinical and research programs. Brain Imaging and Behavior. 2015;9(3):358–366.

2. DoD Numbers for Traumatic Brain Injury Worldwide - Totals (Defense Health Agency) (2021).

3. Karr JE, Areshenkoff CN, Garcia-Barrera MA. The neuropsychological outcomes of concussion: a systematic review of meta-analyses on the cognitive sequelae of mild traumatic brain injury. Neuropsychology. 2014;28(3):321–336.

4. McCrea M, Guskiewicz K, Doncevic S, et al. Day of injury cognitive performance on the Military Acute Concussion Evaluation (MACE) by US military service members in OEF/OIF. Military Medicine. 2014;179(9):990–997.

5. Nelson LD, Temkin NR, Dikmen S, et al. Recovery after mild traumatic brain injury in patients presenting to US level I trauma centers: A transforming research and clinical knowledge in traumatic brain injury (TRACK-TBI) study. JAMA Neurology. 2019;76(9):1049–1059.

6. Chapman JC, Diaz-Arrastia R. Military traumatic brain injury: A review. Alzheimer’s & Dementia. 2014;10:S97–S104.

7. Schwab K, Terrio HP, Brenner LA, et al. Epidemiology and prognosis of mild traumatic brain injury in returning soldiers: A cohort study. Neurology. 2017;88(16):1571–1579.

8. Perry DC, Sturm VE, Peterson MJ, et al. Traumatic brain injury is associated with subsequent neurologic and psychiatric disease: A meta-analysis. Journal of Neurosurgery 2016;124(2):511–526.

9. Stein MB, Ursano RJ, Campbell-Sills L, et al. Prognostic indicators of persistent post-concussive symptoms after deployment-related mild traumatic brain injury: A prospective longitudinal study in US Army soldiers. Journal of Neurotrauma. 2016;33(23):2125–2132.

10. Stein MB, Jain S, Giacino JT, et al. Risk of posttraumatic stress disorder and major depression in civilian patients after mild traumatic brain injury: A TRACK-TBI study. JAMA Psychiatry. 2019;76(3):249–258.

11. Barnes DE, Byers AL, Gardner RC, Seal KH, Boscardin WJ, Yaffe K. Association of mild traumatic brain injury with and without loss of consciousness with dementia in US military veterans. JAMA Neurology. 2018;75(9):1055–1061.

12. Brickell TA, Lange RT, French LM. Health-related quality of life within the first 5 years following military-related concurrent mild traumatic brain injury and polytrauma. Military Medicine. 2014;179(8):827–838.

13. Sakamoto MS, Delano-Wood L, Schiehser DM, Merritt VC. Predicting veteran health-related quality of life following mild traumatic brain injury. Rehabilitation Psychology. 2021;doi:10.1037/rep0000392

14. Agtarap SD, Campbell-Sills L, Jain S, et al. Satisfaction with life after mild traumatic brain injury: A TRACK-TBI study. Journal of Neurotrauma. 2021;38(5):546–554.

15. Pietrzak RH, Johnson DC, Goldstein MB, Malley JC, Southwick SM. Posttraumatic stress disorder mediates the relationship between mild traumatic brain injury and health and psychosocial functioning in veterans of Operations Enduring 1. Freedom and Iraqi Freedom. The Journal of nervous and mental disease. 2009;197(10):748–753.

16. MacDonald CL, Barber J, Jordan M, et al. Early clinical predictors of 5-year outcome after concussive blast traumatic brain injury. JAMA Neurology. 2017;74(7):821–829.

17. Taylor BC, Hagel EM, Carlson KF, et al. Prevalence and costs of co-occurring traumatic brain injury with and without psychiatric disturbance and pain among Afghanistan and Iraq War Veteran VA users. Medical Care. 2012:342–346.

18. McAllister TW. Genetic factors modulating outcome after neurotrauma. PM&R. 2010;2(12):S241–S252.

19. Jordan BD. Genetic influences on outcome following traumatic brain injury. Neurochemical Research. 2007;32(4):905–915.

20. Weaver SM, Chau A, Portelli JN, Grafman J. Genetic polymorphisms influence recovery from traumatic brain injury. The Neuroscientist. 2012;18(6):631–644.

21. Dardiotis E, Fountas KN, Dardioti M, et al. Genetic association studies in patients with traumatic brain injury. Neurosurgical Focus. 2010;28(1):E9.

22. Zeiler FA, McFadyen C, Newcombe VF, et al. Genetic influences on patient-oriented outcomes in traumatic brain injury: A living systematic review of non-apolipoprotein E single-nucleotide polymorphisms. Journal of Neurotrauma. 2021;38(8):1107–1123.

23. Panenka WJ, Gardner AJ, Dretsch MN, Crynen GC, Crawford FC, Iverson GL. Systematic review of genetic risk factors for sustaining a mild traumatic brain injury. Journal of Neurotrauma. 2017;34(13):2093–2099.

24. Lawrence DW, Comper P, Hutchison MG, Sharma B. The role of apolipoprotein E episilon (L)-4 allele on outcome following traumatic brain injury: A systematic review. Brain Injury. 2015;29(9):1018–1031.

25. Sun X-C, Jiang Y. Genetic susceptibility to traumatic brain injury and apolipoprotein E gene. Chinese Journal of Traumatology. 2008;11(4):247–252.

26. Zhou W, Xu D, Peng X, Zhang Q, Jia J, Crutcher KA. Meta-analysis of APOE 4 allele and outcome after traumatic brain injury. Journal of Neurotrauma. 2008;25(4):279–290.

27. Zeng S, Jiang J-X, Xu M-H, et al. Prognostic value of apolipoprotein E epsilon4 allele in patients with traumatic brain injury: A meta-analysis and meta-regression. Genetic Testing and Molecular Biomarkers. 2014;18(3):202–210.

28. McFadyen CA, Zeiler FA, Newcombe V, et al. Apolipoprotein E4 polymorphism and outcomes from traumatic brain injury: A living systematic review and meta-analysis. Journal of Neurotrauma. 2021;38(8):1124–1136.

29. Gaziano JM, Concato J, Brophy M, et al. Million Veteran Program: A mega-biobank to study genetic influences on health and disease. Journal of Clinical Epidemiology. 2016;70:214–223.

30. Schwab K, Ivins B, Cramer G, et al. Screening for traumatic brain injury in troops returning from deployment in Afghanistan and Iraq: initial investigation of the usefulness of a short screening tool for traumatic brain injury. The Journal of Head Trauma Rehabilitation. 2007;22(6):377–389.

31. U.S. Department of Defense. Traumatic Brain Injury (TBI): DoD Standard Surveillance Case Definition for TBI Adapted for AFHSD Use. U.S. Department of Defense.

32. The 1000 Genomes Project Consortium. A global reference for human genetic variation. Nature. 2015;526(7571):68–74.

33. Loh P-R, Danecek P, Palamara PF, et al. Reference-based phasing using the Haplotype Reference Consortium panel. Nature Genetics. 2016;48(11):1443–1448.

34. Fang H, Hui Q, Lynch J, et al. Harmonizing genetic ancestry and self-identified race/ethnicity in genome-wide association studies. The American Journal of Human Genetics. 2019;105(4):763–772.

35. Manichaikul A, Mychaleckyj JC, Rich SS, Daly K, Sale M, Chen W-M. Robust relationship inference in genome-wide association studies. Bioinformatics. 2010;26(22):2867–2873.

36. Abraham G, Qiu Y, Inouye M. FlashPCA2: Principal component analysis of Biobank-scale genotype datasets. Bioinformatics. 2017;33(17):2776–2778.

37. Chang CC, Chow CC, Tellier LC, Vattikuti S, Purcell SM, Lee JJ. Second-generation PLINK: rising to the challenge of larger and richer datasets. Gigascience. 2015;4(7)

38. Willer CJ, Li Y, Abecasis GR. METAL: Fast and efficient meta-analysis of genomewide association scans. Bioinformatics. 2010;26(17):2190–2191.

39. Pruim RJ, Welch RP, Sanna S, et al. LocusZoom: Regional visualization of genome-wide association scan results. Bioinformatics. 2010;26(18):2336–2337.

40. Watanabe K, Taskesen E, Van Bochoven A, Posthuma D. Functional mapping and annotation of genetic associations with FUMA. Nature Communications. 2017;8(1):1826.

41. Lee SH, Wray NR, Goddard ME, Visscher PM. Estimating missing heritability for disease from genome-wide association studies. The American Journal of Human Genetics. 2011;88(3):294–305.

42. Bulik-Sullivan BK, Loh P-R, Finucane HK, et al. LD Score regression distinguishes confounding from polygenicity in genome-wide association studies. Nature Genetics. 2015;47(3):291–295.

43. Holland D, Frei O, Desikan R, et al. Beyond SNP heritability: Polygenicity and discoverability of phenotypes estimated with a univariate Gaussian mixture model. PLoS Genetics. 2020;16(5):e1008612.

44. Frei O, Holland D, Smeland OB, et al. Bivariate causal mixture model quantifies polygenic overlap between complex traits beyond genetic correlation. Nature Communications. 2019;10:2417.

45. R Core Team. R: A language and environment for statistical computing. https://www.R-project.org/

46. Grotzinger AD, Rhemtulla M, de Vlaming R, et al. Genomic structural equation modelling provides insights into the multivariate genetic architecture of complex traits. Nature Human Behaviour. 2019;3(5):513–525.

47. Watanabe Kea. A global view of genetic architecture in human complex traits. [under preparation].

48. Seripa D, D’Onofrio G, Panza F, Cascavilla L, Masullo C, Pilotto A. The genetics of the human APOE polymorphism. Rejuvenation Research. 2011;14(5):491-500.1.

49. Broman KW, Matsumoto N, Giglio S, et al. Common long human inversion polymorphism on chromosome 8p. Lecture Notes-Monograph Series. 2003:237–245.

50. Schmid RS, Maness PF. Cell recognition molecules and disorders of neurodevelopment. International Handbook on Brain and Behaviour in Human Development. Kluwer Academic Publishers; 2001:199–218.

51. Brennaman LH, Maness PF. NCAM in neuropsychiatric and neurodegenerative disorders. vol 663. Structure and Function of the Neural Cell Adhesion Molecule NCAM: Advances in Experimental Medicine and Biology Springer; 2010:299–317.

52. Vukojevic V, Mastrandreas P, Arnold A, et al. Evolutionary conserved role of neural cell adhesion molecule-1 in memory. Translational Psychiatry. 2020;10:217.

53. Mahley RW, Huang Y. Apolipoprotein e sets the stage: Response to injury triggers neuropathology. Neuron. 2012;76(5):871–885.

54. Mahley RW, Weisgraber KH, Huang Y. Apolipoprotein E4: A causative factor and therapeutic target in neuropathology, including Alzheimer’s disease. Proceedings of the National Academy of Sciences. 2006;103(15):5644–5651.

55. Farrer LA, Cupples LA, Haines JL, et al. Effects of age, sex, and ethnicity on the association between apolipoprotein E genotype and Alzheimer disease: A meta-analysis. JAMA. 1997;278(16):1349–1356.

56. Horsburgh K, McCarron MO, White F, Nicoll JA. The role of apolipoprotein E in Alzheimer’s disease, acute brain injury and cerebrovascular disease: Evidence of common mechanisms and utility of animal models. Neurobiology of Aging. 2000;21(2):245–255.

57. Verghese PB, Castellano JM, Holtzman DM. Apolipoprotein E in Alzheimer’s disease and other neurological disorders. The Lancet Neurology. 2011;10(3):241–252.

58. Reinvang I, Espeseth T, Westlye LT. APOE-related biomarker profiles in non-pathological aging and early phases of Alzheimer’s disease. Neuroscience & Biobehavioral Reviews. 2013;37(8):1322–1335.

59. Van Giau V, Bagyinszky E, An SSA, Kim SY. Role of apolipoprotein E in neurodegenerative diseases. Neuropsychiatric Disease and Treatment. 2015;11:1723–1737.

60. Wisdom NM, Callahan JL, Hawkins KA. The effects of apolipoprotein E on non-impaired cognitive functioning: A meta-analysis. Neurobiology of Aging. 2011;32(1):63–74.

61. McAllister TW, Flashman LA, Harker Rhodes C, et al. Single nucleotide polymorphisms in ANKK1 and the dopamine D2 receptor gene affect cognitive outcome shortly after traumatic brain injury: A replication and extension study. Brain Injury. 2008;22(9):705–714.

62. Merritt VC, Clark AL, Sorg SF, et al. Apolipoprotein E ε4 genotype is associated with elevated psychiatric distress in veterans with a history of mild to moderate traumatic brain injury. Journal of Neurotrauma. 2018;35(19):2272–2282.

63. Merritt VC, Clark AL, Sorg SF, et al. Apolipoprotein E (APOE) ε4 genotype is associated with reduced neuropsychological performance in military veterans with a history of mild traumatic brain injury. Journal of Clinical and Experimental Neuropsychology. 2018;40(10):1050-1061.1.

64. Merritt VC, Lange RT, Lippa SM, et al. Apolipoprotein e (APOE) ε4 genotype influences memory performance following remote traumatic brain injury in US military service members and veterans. Brain and cognition. 2021;154:105790.

65. Zhao X, Yang Y, Sun B-F, Zhao Y-L, Yang Y-G. FTO and obesity: Mechanisms of association. Current Diabetes Reports. 2014;14(5):1–9.

66. McCarthy MI. Genomics, type 2 diabetes, and obesity. New England Journal of Medicine. 2010;363(24):2339–2350.

67. Reitz C, Tosto G, Mayeux R, Luchsinger JA, Group N-LNFS, Initiative tAsDN. Genetic variants in the Fat and Obesity Associated (FTO) gene and risk of Alzheimer’s disease. PloS One. 2012;7(12):e50354.

68. Li H, Ren Y, Mao K, et al. FTO is involved in Alzheimer’s disease by targeting TSC1-mTOR-Tau signaling. Biochemical and Biophysical Research Communications. 2018;498(1):234–239.

69. Co M, Anderson AG, Konopka G. FOXP transcription factors in vertebrate brain development, function, and disorders. Wiley Interdisciplinary Reviews: Developmental Biology. 2020;9(5):e375.

70. Den Hoed J, Devaraju K, Fisher SE. Molecular networks of the FOXP2 transcription factor in the brain. EMBO Reports. 2021;22(8):e52803.

71. Stein MB, Levey DF, Cheng Z, et al. Genome-wide association analyses of post-traumatic stress disorder and its symptom subdomains in the Million Veteran Program. Nature Genetics. 2021;53(2):174–184.

72. Linnér RK, Biroli P, Kong E, et al. Genome-wide association analyses of risk tolerance and risky behaviors in over 1 million individuals identify hundreds of loci and shared genetic influences. Nature Genetics. 2019;51(2):245–257.

73. Bush G, Luu P, Posner MI. Cognitive and emotional influences in anterior cingulate cortex. Trends in Cognitive Sciences. 2000;4(6):215–222.

74. Stevens FL, Hurley RA, Taber KH. Anterior cingulate cortex: unique role in cognition and emotion. The Journal of Neuropsychiatry and Clinical Neurosciences. 2011;23(2):121–125.

75. Kunkle BW, Grenier-Boley B, Sims R, et al. Genetic meta-analysis of diagnosed Alzheimer’s disease identifies new risk loci and implicates Aβ, tau, immunity and lipid processing. Nature Genetics. 2019;51(3):414–430.

