## Supplemental Figures for "Genome-wide Association Study of Traumatic Brain Injury in U.S. Military Veterans Enrolled in the VA Million Veteran Program"

Supplemental Figure 1 (1a-1aa). LocusZoom plots.

Supplemental Figure 2. Manhattan plot of TBI risk for the European ancestry cohort.

Supplemental Figure 3. Manhattan plot of TBI risk for the African ancestry cohort.

Supplemental Figure 4. Manhattan plot of TBI risk for the Hispanic ancestry cohort.

Supplemental Figure 5a. Gene-based results for multi-ancestry cohort.

Supplemental Figure 5b. Gene-based results for European ancestry cohort.

Supplemental Figure 6a. Gene tissue expression (tissues aggregated) in the European ancestry cohort.

Supplemental Figure 6b. Gene tissue expression (stratified by tissue subtype) in the European ancestry cohort.

Supplemental Figure 7. LDSC correlations between TBI and risk-taking behaviors, psychiatric disorders, and neurocognition.

Supplemental Figure 8. LDSC correlations between TBI and ENIGMA variables.

Supplemental Figure 9. Bivariate MiXeR analysis of TBI and Alzheimer's disease.

Supplemental Figure 10. Bivariate MiXeR analysis of TBI and reaction time.

Supplemental Figure 11. PheWAS plots for European ancestry cohort.

Supplemental Figure 12. PheWAS results for TBI risk loci.

**Supplemental Figure 1 (1a-1aa).** LocusZoom plots.

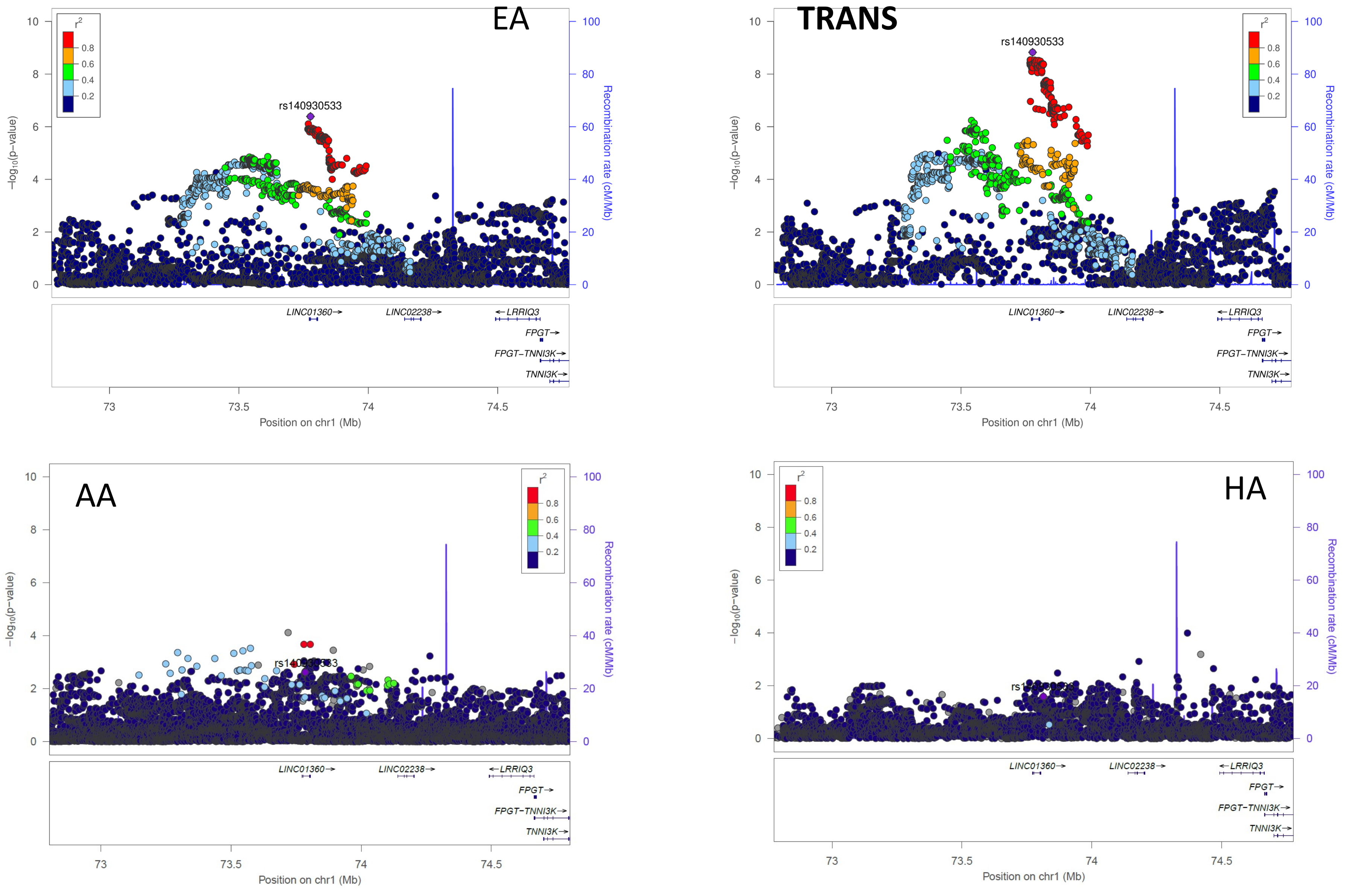


**Supplemental Figure 1a: Comparison of the European ancestry hit rs140930533 across European ancestry (EA), Transethnic ancestry (TRANS), African ancestry (AA), and Hispanic ancestry (HA) TBI studies.** Chromosomal position of the regional association plots is indicated on the x-axis, -log10 *p* values for each SNP (filled circles) is indicated on the y-axis, with the lead SNP shown in purple. Annotated genes in the region are drawn in the lower panel. Recombination rate is indicated by a blue line. Additional SNPs in the locus are colored according to linkage disequilibrium (*r*2) with the lead SNP.

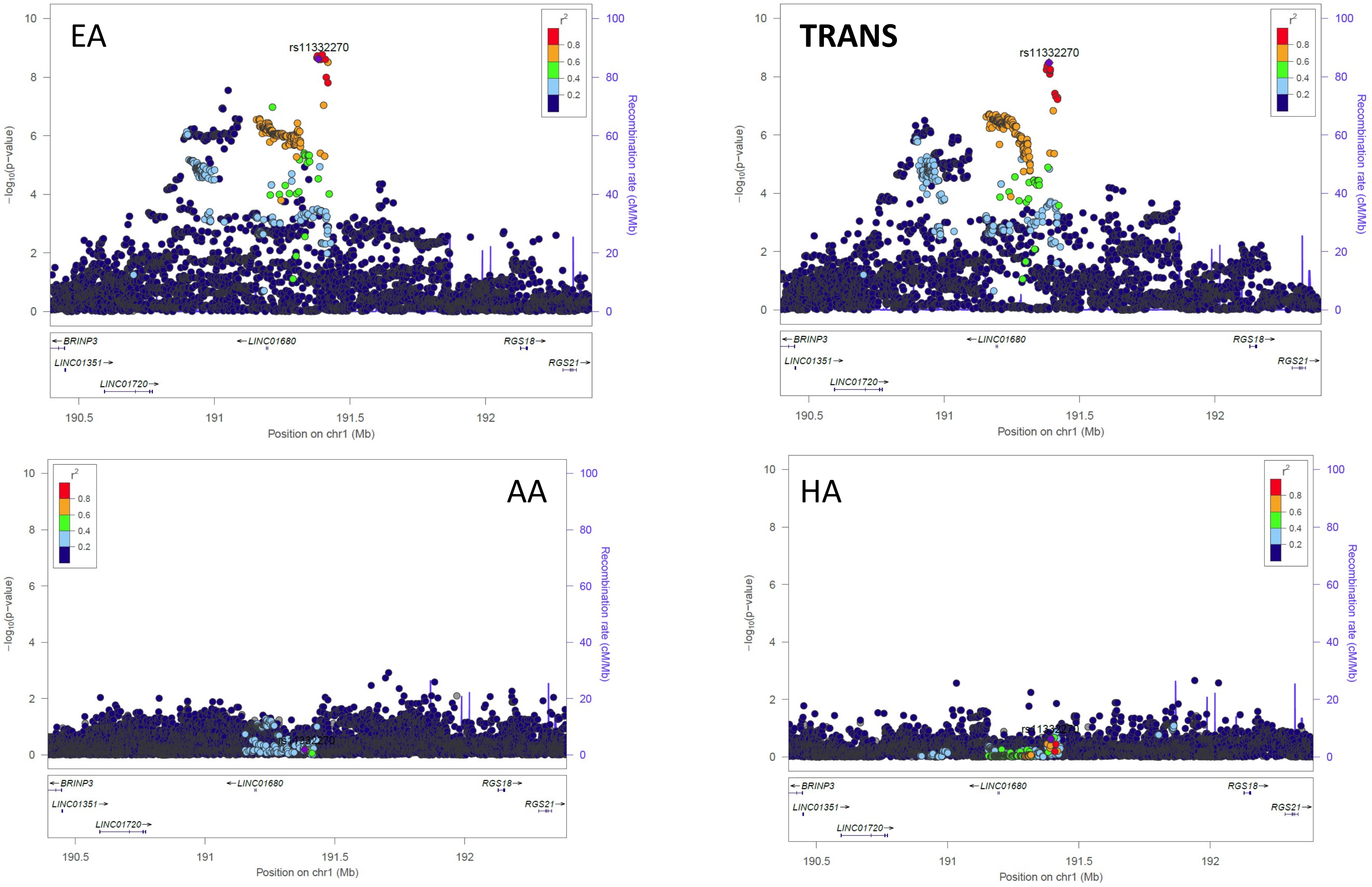


**Supplemental Figure 1b: Comparison of the European ancestry hit rs11332270 across European ancestry (EA), Transethnic ancestry (TRANS), African ancestry (AA), and Hispanic ancestry (HA) TBI studies.** Chromosomal position of the regional association plots is indicated on the x-axis, -log10 *p* values for each SNP (filled circles) is indicated on the y-axis, with the lead SNP shown in purple. Annotated genes in the region are drawn in the lower panel. Recombination rate is indicated by a blue line. Additional SNPs in the locus are colored according to linkage disequilibrium (*r*2) with the lead SNP.

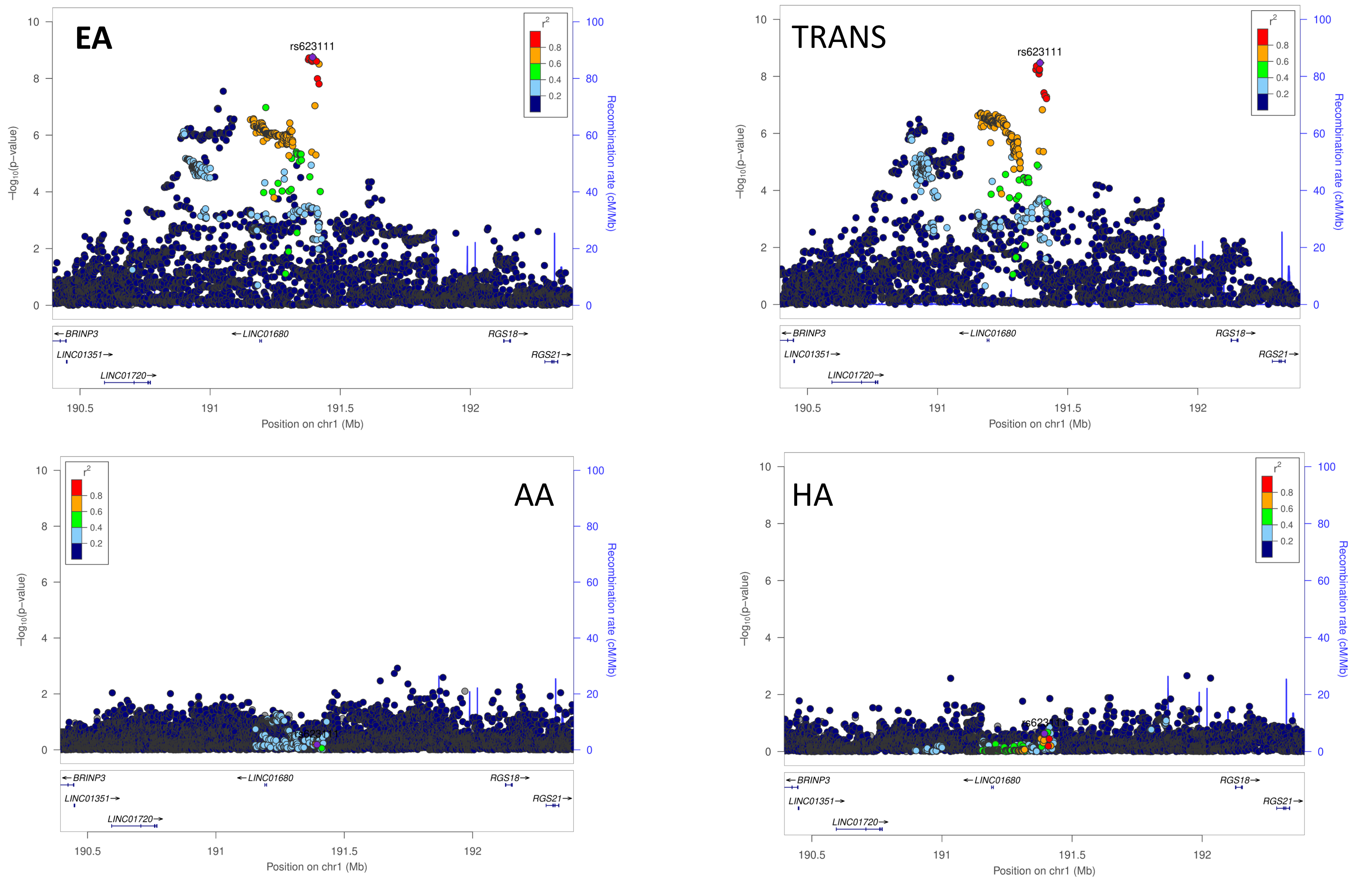


**Supplemental Figure 1c: Comparison of the European ancestry hit rs623111 across European ancestry (EA), Transethnic ancestry (TRANS), African ancestry (AA), and Hispanic ancestry (HA) TBI studies.** Chromosomal position of the regional association plots is indicated on the x-axis, -log10 *p* values for each SNP (filled circles) is indicated on the y-axis, with the lead SNP shown in purple. Annotated genes in the region are drawn in the lower panel. Recombination rate is indicated by a blue line. Additional SNPs in the locus are colored according to linkage disequilibrium (*r*2) with the lead SNP.

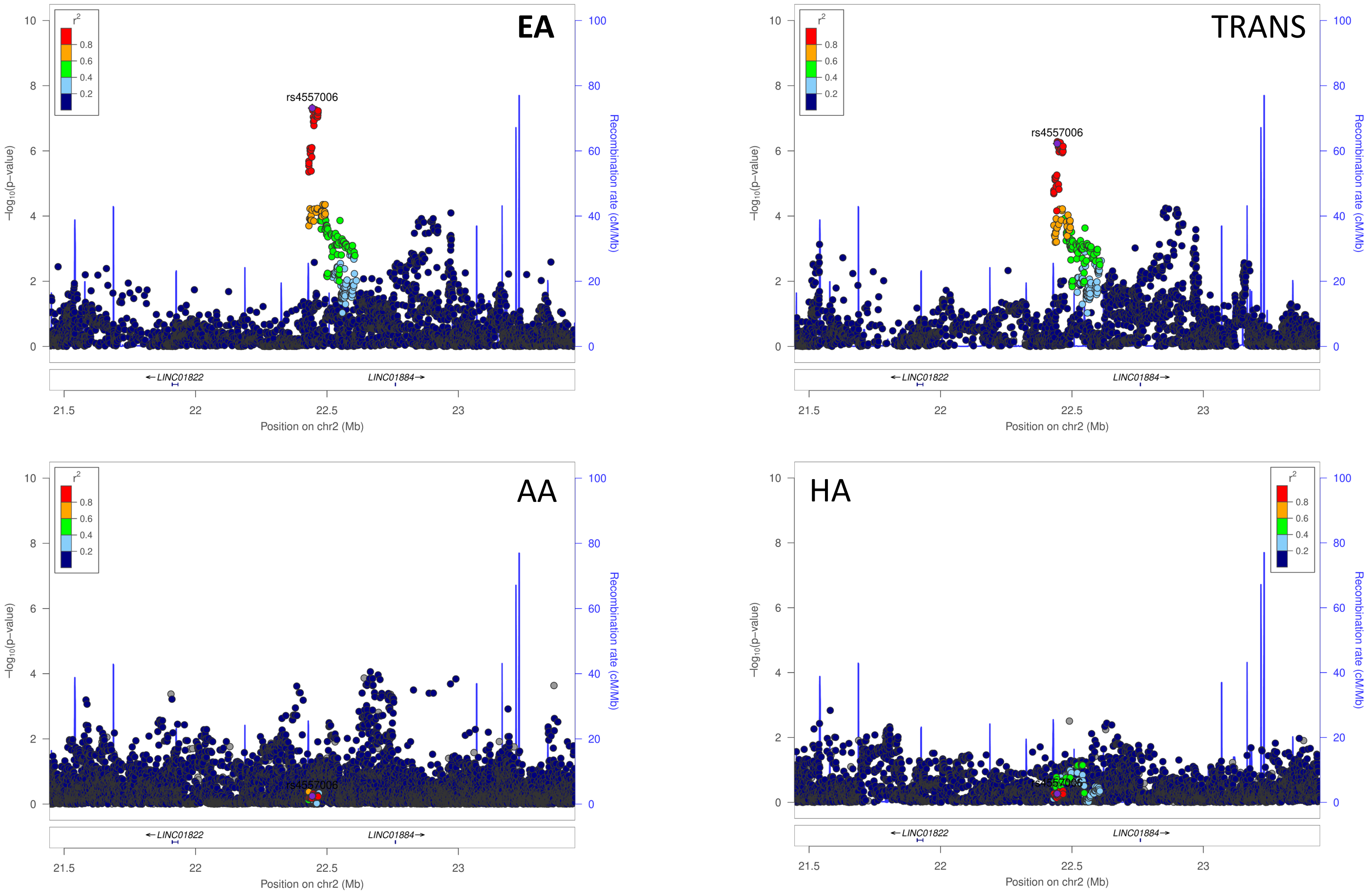


**Supplemental Figure 1d: Comparison of the European ancestry hit rs4557006 across European ancestry (EA), Transethnic ancestry (TRANS), African ancestry (AA), and Hispanic ancestry (HA) TBI studies.** Chromosomal position of the regional association plots is indicated on the x-axis, -log10 *p* values for each SNP (filled circles) is indicated on the y-axis, with the lead SNP shown in purple. Annotated genes in the region are drawn in the lower panel. Recombination rate is indicated by a blue line. Additional SNPs in the locus are colored according to linkage disequilibrium (*r*2) with the lead SNP.

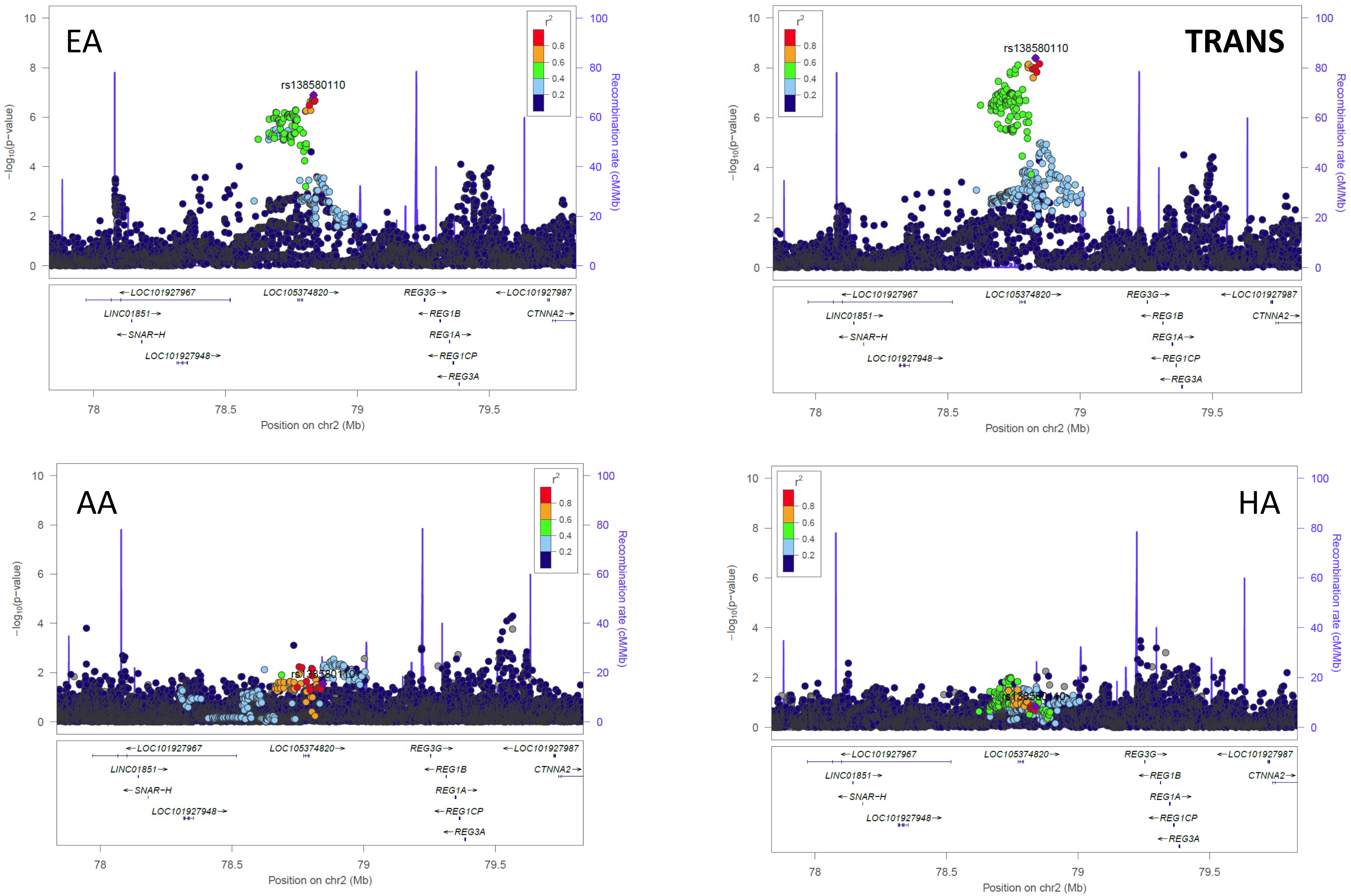


**Supplemental Figure 1e: Comparison of the European ancestry hit rs138580110 across European ancestry (EA), Transethnic ancestry (TRANS), African ancestry (AA), and Hispanic ancestry (HA) TBI studies.** Chromosomal position of the regional association plots is indicated on the x-axis, -log10 *p* values for each SNP (filled circles) is indicated on the y-axis, with the lead SNP shown in purple. Annotated genes in the region are drawn in the lower panel. Recombination rate is indicated by a blue line. Additional SNPs in the locus are colored according to linkage disequilibrium (*r*2) with the lead SNP.

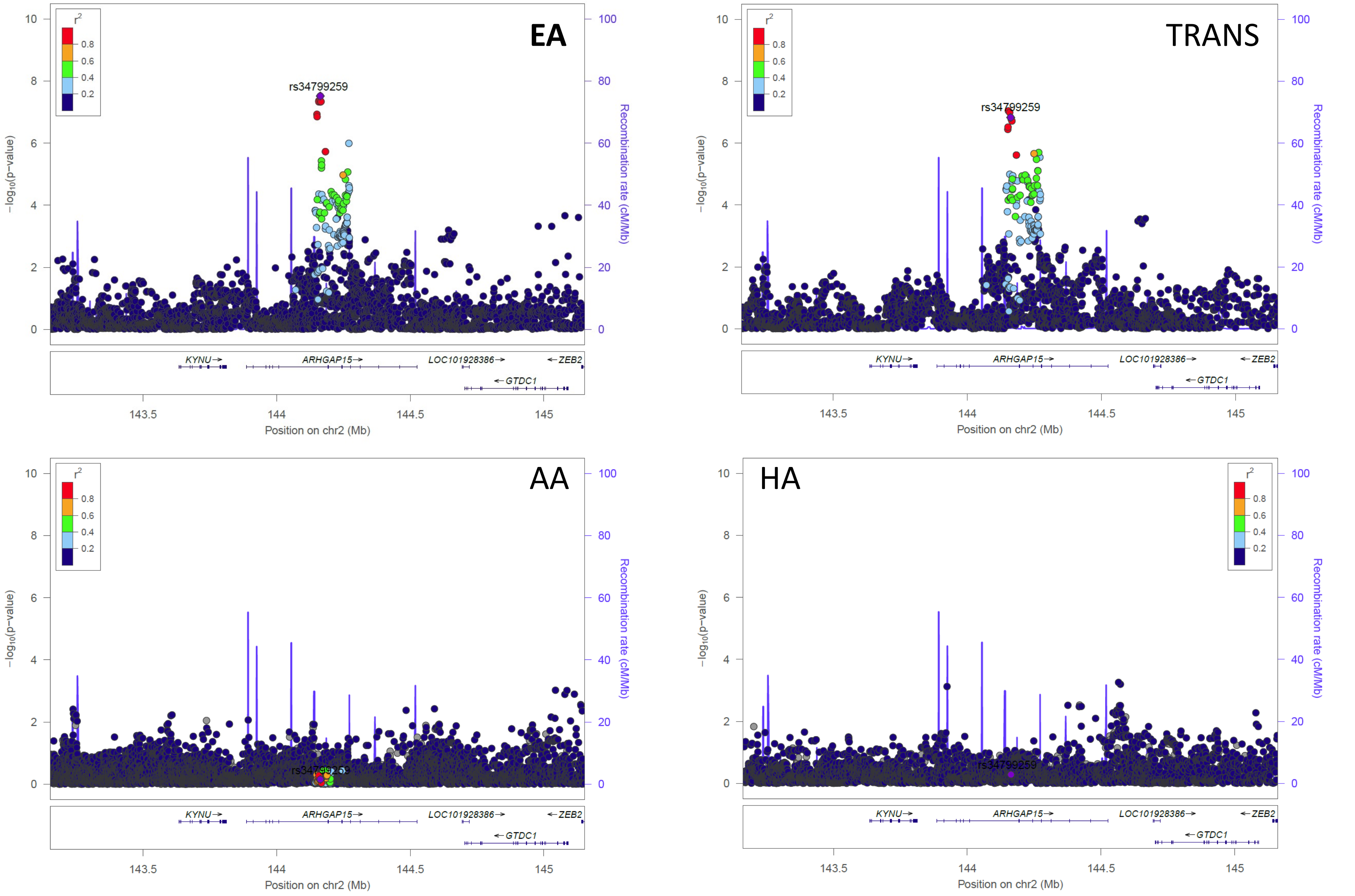


**Supplemental Figure 1f: Comparison of the European ancestry hit rs34799259 across European ancestry (EA), Transethnic ancestry (TRANS), African ancestry (AA), and Hispanic ancestry (HA) TBI studies.** Chromosomal position of the regional association plots is indicated on the x-axis, -log10 *p* values for each SNP (filled circles) is indicated on the y-axis, with the lead SNP shown in purple. Annotated genes in the region are drawn in the lower panel. Recombination rate is indicated by a blue line. Additional SNPs in the locus are colored according to linkage disequilibrium (*r*2) with the lead SNP.

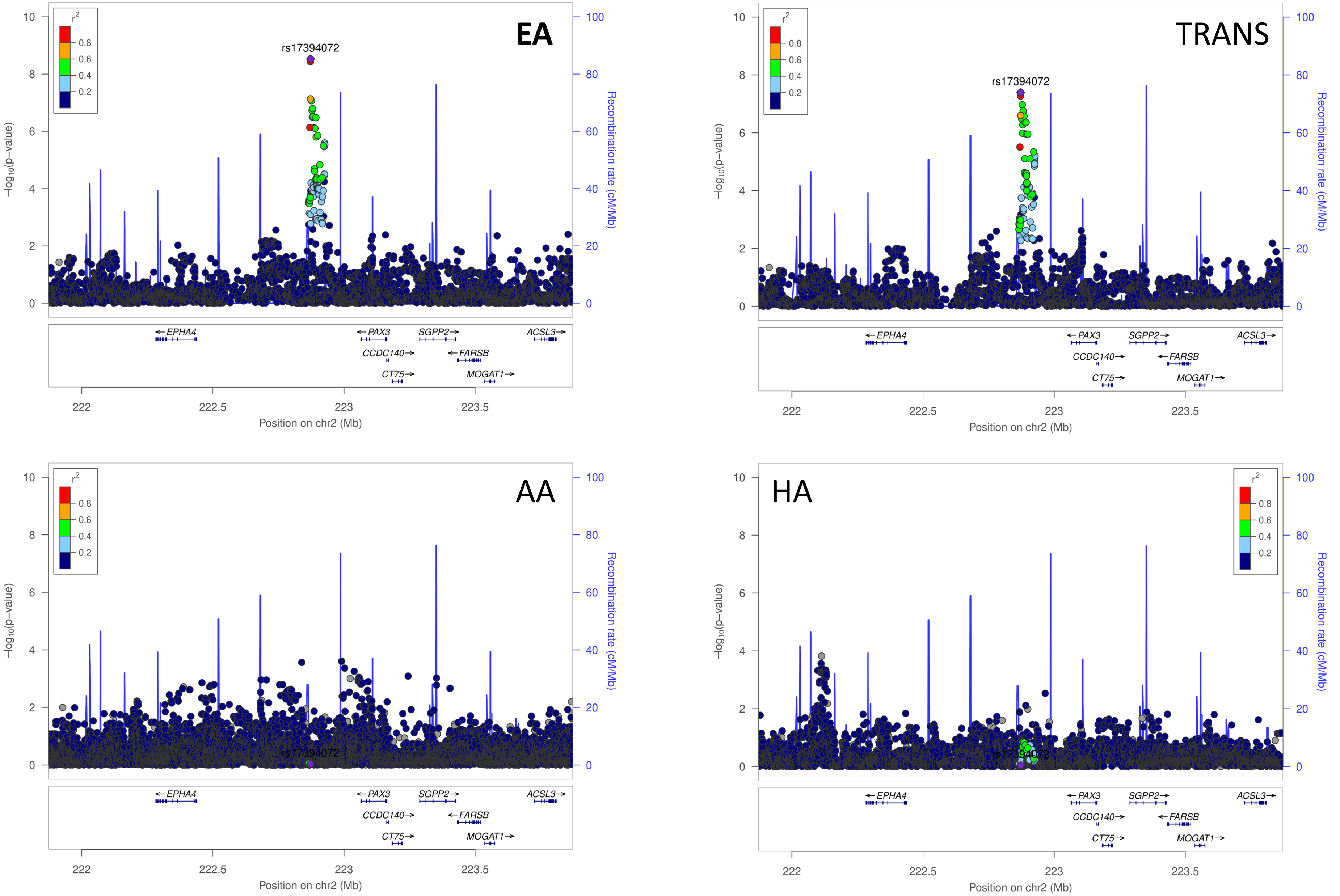


**Supplemental Figure 1g: Comparison of the European ancestry hit rs17394072 across European ancestry (EA), Transethnic ancestry (TRANS), African ancestry (AA), and Hispanic ancestry (HA) TBI studies.** Chromosomal position of the regional association plots is indicated on the x-axis, -log10 *p* values for each SNP (filled circles) is indicated on the y-axis, with the lead SNP shown in purple. Annotated genes in the region are drawn in the lower panel. Recombination rate is indicated by a blue line. Additional SNPs in the locus are colored according to linkage disequilibrium (*r*2) with the lead SNP.

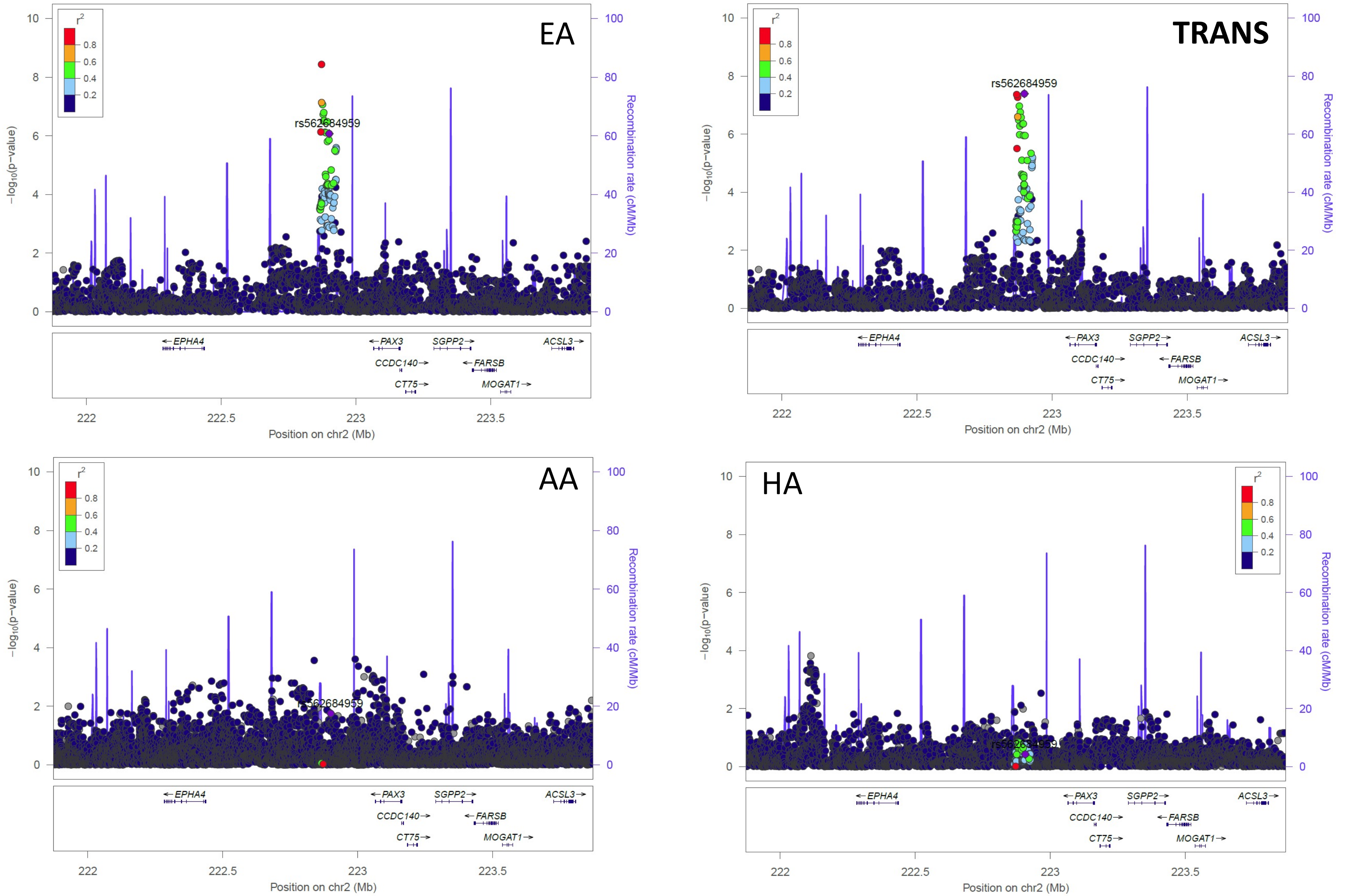


**Supplemental Figure 1h: Comparison of the European ancestry hit rs562684959 across European ancestry (EA), Transethnic ancestry (TRANS), African ancestry (AA), and Hispanic ancestry (HA) TBI studies.** Chromosomal position of the regional association plots is indicated on the x-axis, -log10 *p* values for each SNP (filled circles) is indicated on the y-axis, with the lead SNP shown in purple. Annotated genes in the region are drawn in the lower panel. Recombination rate is indicated by a blue line. Additional SNPs in the locus are colored according to linkage disequilibrium (*r*2) with the lead SNP.

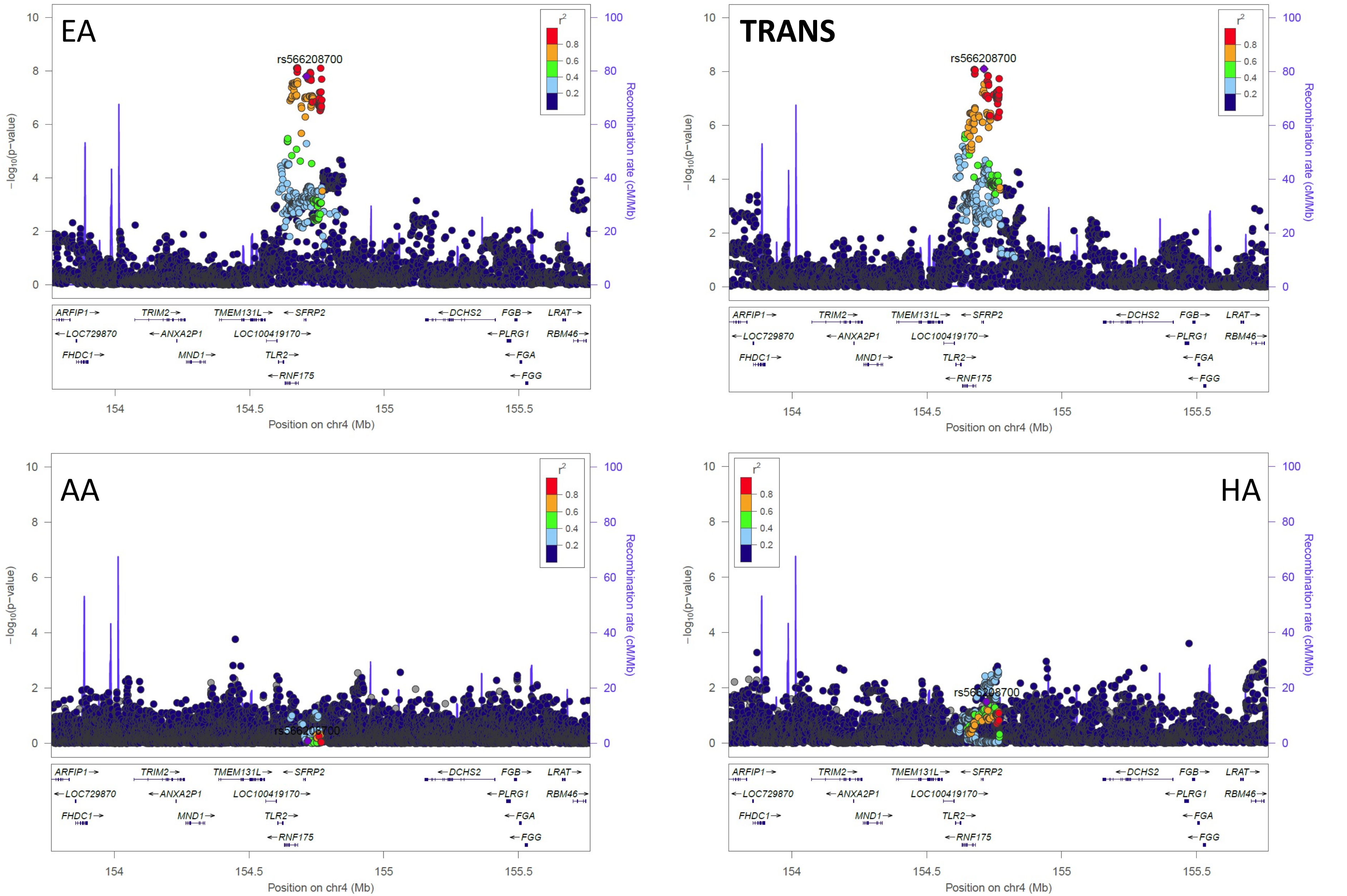


**Supplemental Figure 1i: Comparison of the European ancestry hit rs566208700 across European ancestry (EA), Transethnic ancestry (TRANS), African ancestry (AA), and Hispanic ancestry (HA) TBI studies.**
Chromosomal position of the regional association plots is indicated on the x-axis, -log10 *p* values for each SNP (filled circles) is indicated on the y-axis, with the lead SNP shown in purple. Annotated genes in the region are drawn in the lower panel. Recombination rate is indicated by a blue line. Additional SNPs in the locus are colored according to linkage disequilibrium (*r*2) with the lead SNP.

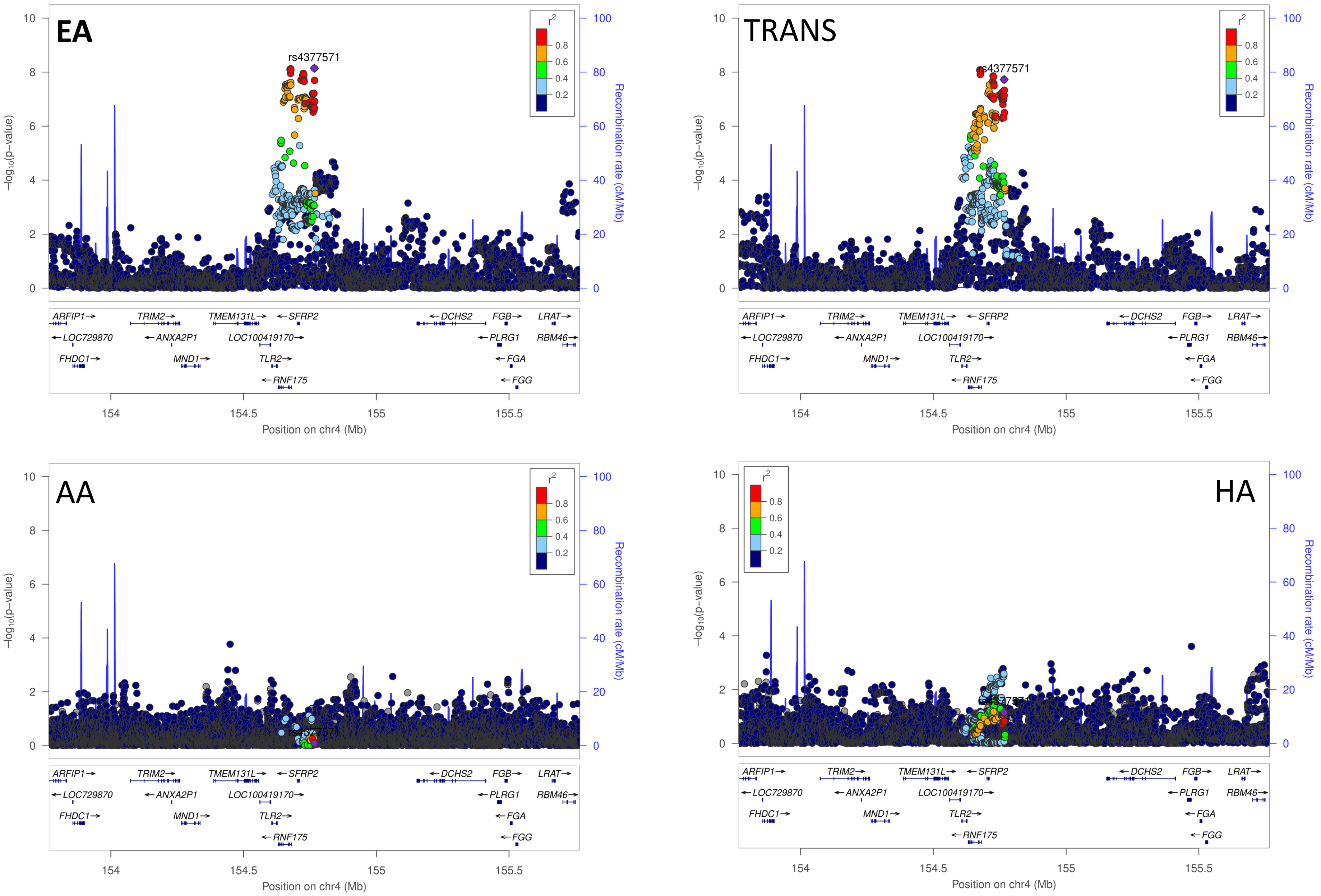


**Supplemental Figure 1j: Comparison of the European ancestry hit rs4377571 across European ancestry (EA), Transethnic ancestry (TRANS), African ancestry (AA), and Hispanic ancestry (HA) TBI studies.** Chromosomal position of the regional association plots is indicated on the x-axis, -log10 *p* values for each SNP (filled circles) is indicated on the y-axis, with the lead SNP shown in purple. Annotated genes in the region are drawn in the lower panel. Recombination rate is indicated by a blue line. Additional SNPs in the locus are colored according to linkage disequilibrium (*r*2) with the lead SNP.

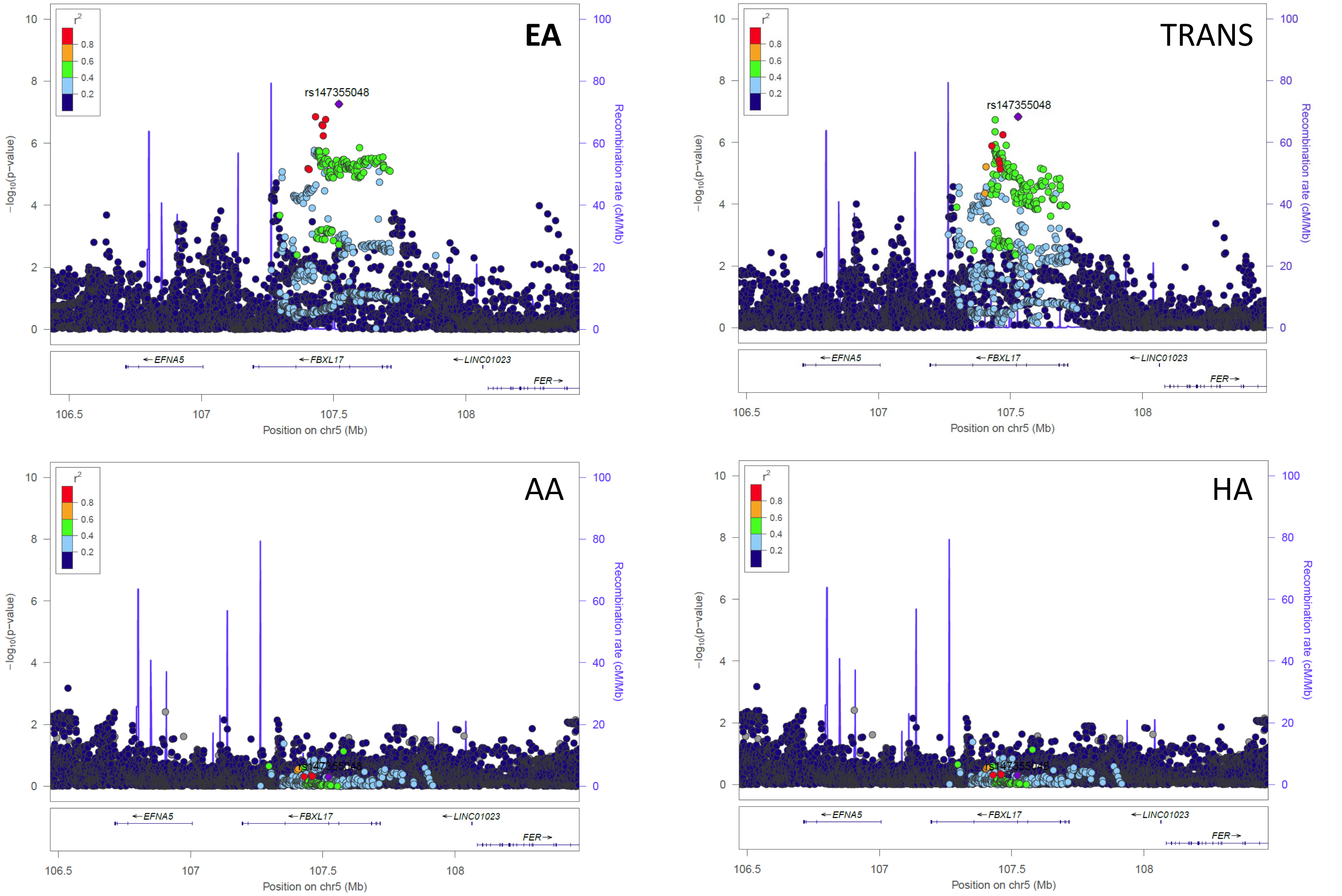


**Supplemental Figure 1k: Comparison of the European ancestry hit rs147355048 across European ancestry (EA), Transethnic ancestry (TRANS), African ancestry (AA), and Hispanic ancestry (HA) TBI studies.**
Chromosomal position of the regional association plots is indicated on the x-axis, -log10 *p* values for each SNP (filled circles) is indicated on the y-axis, with the lead SNP shown in purple. Annotated genes in the region are drawn in the lower panel. Recombination rate is indicated by a blue line. Additional SNPs in the locus are colored according to linkage disequilibrium (*r*2) with the lead SNP.

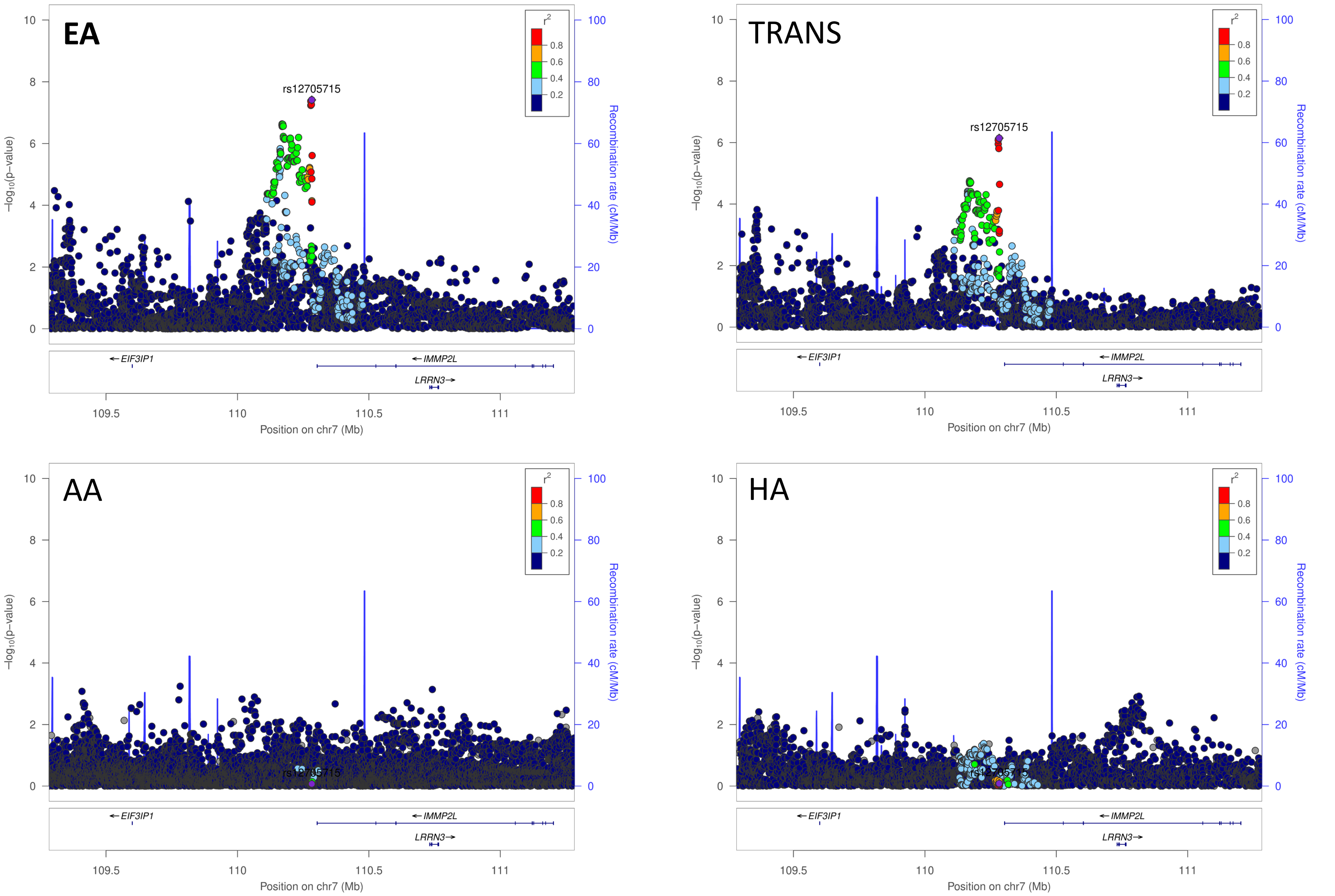


**Supplemental Figure 1l: Comparison of the European ancestry hit rs12705715 across European ancestry (EA), Transethnic ancestry (TRANS), African ancestry (AA), and Hispanic ancestry (HA) TBI studies.** Chromosomal position of the regional association plots is indicated on the x-axis, -log10 *p* values for each SNP (filled circles) is indicated on the y-axis, with the lead SNP shown in purple. Annotated genes in the region are drawn in the lower panel. Recombination rate is indicated by a blue line. Additional SNPs in the locus are colored according to linkage disequilibrium (*r*2) with the lead SNP.

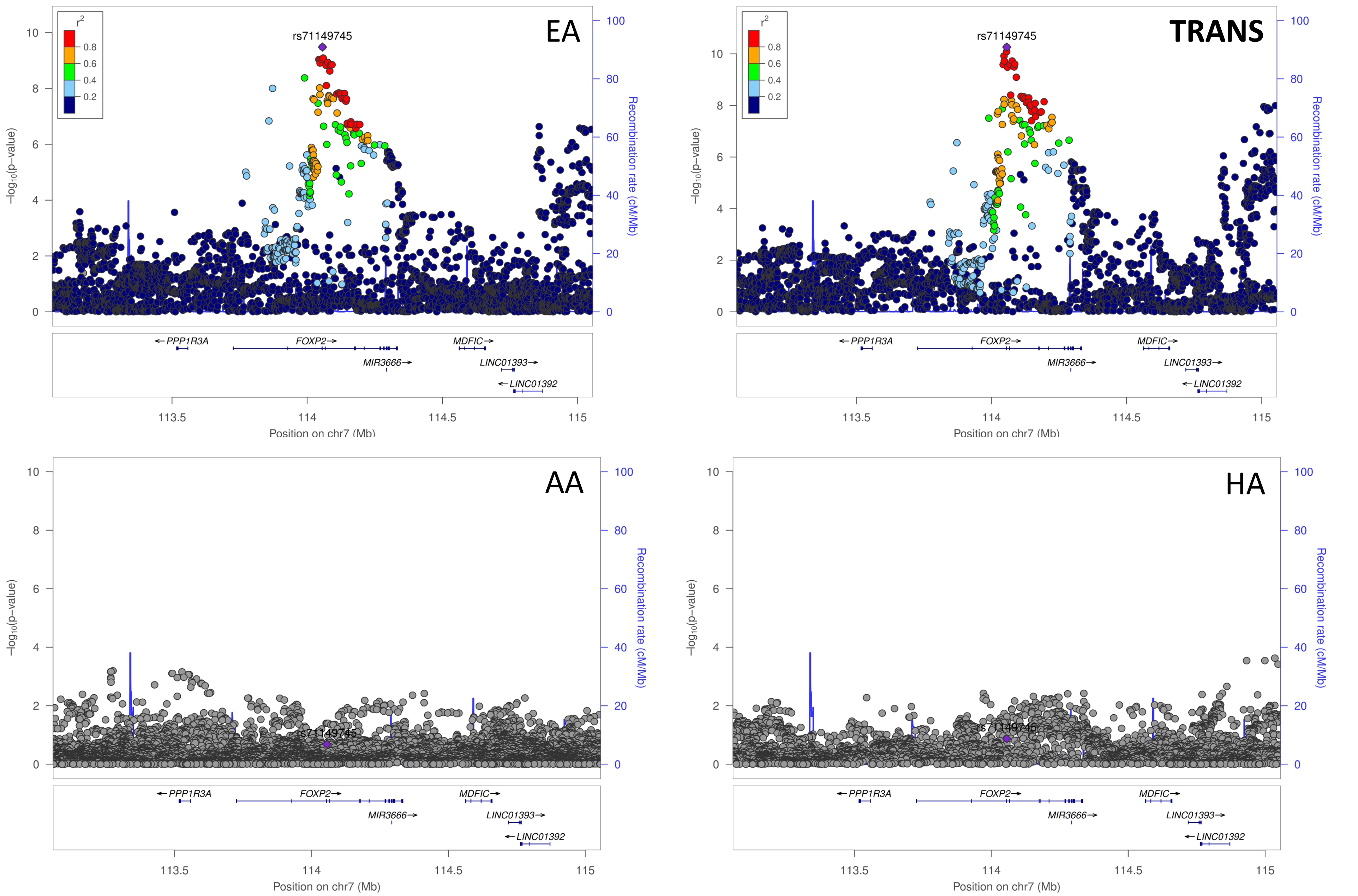


**Supplemental Figure 1m: Comparison of the European ancestry hit rs71149745 across European ancestry (EA), Transethnic ancestry (TRANS), African ancestry (AA), and Hispanic ancestry (HA) TBI studies.** Chromosomal position of the regional association plots is indicated on the x-axis, -log10 *p* values for each SNP (filled circles) is indicated on the y-axis, with the lead SNP shown in purple. Annotated genes in the region are drawn in the lower panel. Recombination rate is indicated by a blue line. Additional SNPs in the locus are colored according to linkage disequilibrium (*r*2) with the lead SNP.

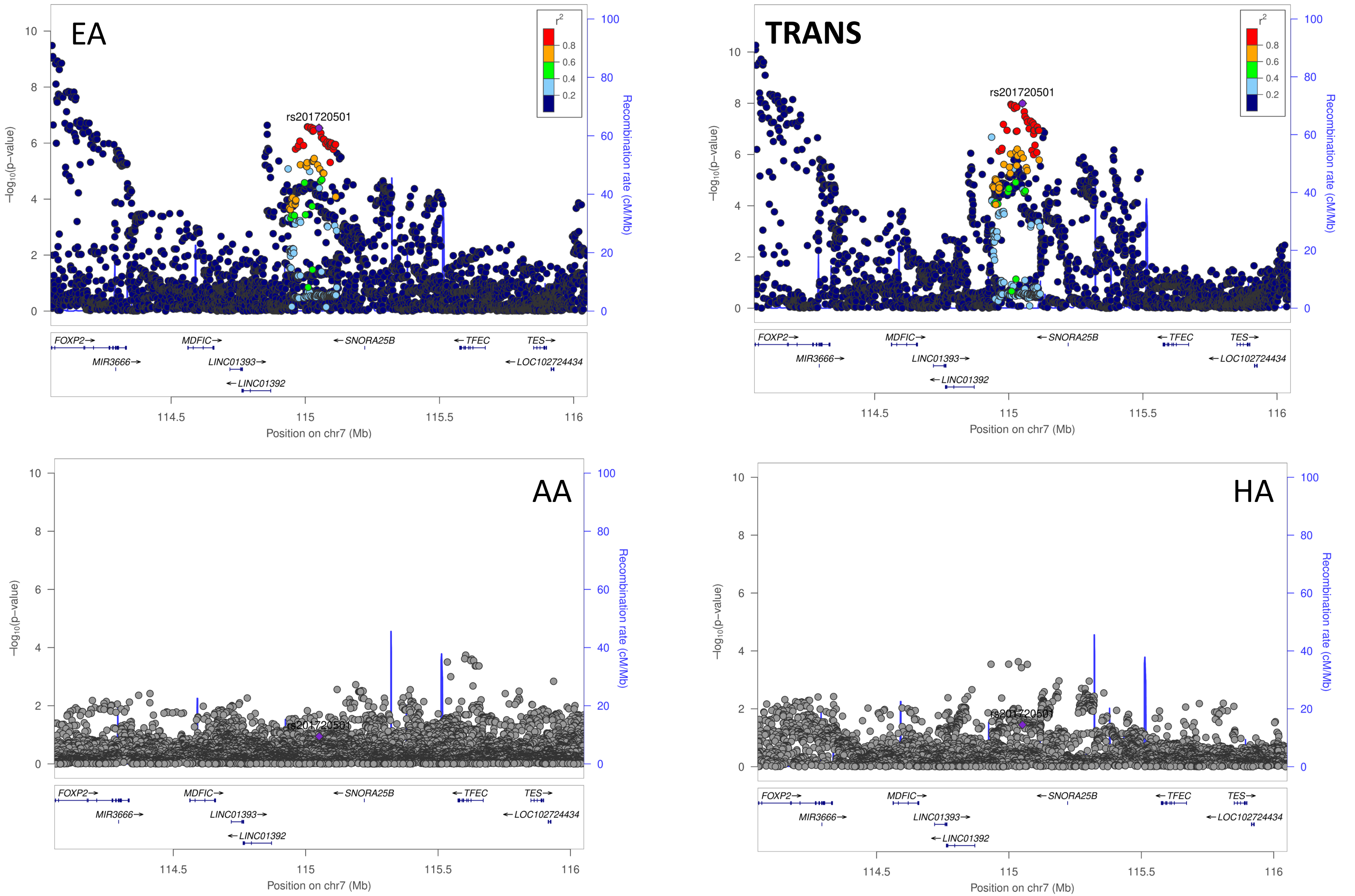


**Supplemental Figure 1n: Comparison of the European ancestry hit rs201720501 across European ancestry (EA), Transethnic ancestry (TRANS), African ancestry (AA), and Hispanic ancestry (HA) TBI studies.** Chromosomal position of the regional association plots is indicated on the x-axis, -log10 *p* values for each SNP (filled circles) is indicated on the y-axis, with the lead SNP shown in purple. Annotated genes in the region are drawn in the lower panel. Recombination rate is indicated by a blue line. Additional SNPs in the locus are colored according to linkage disequilibrium (*r*2) with the lead SNP.

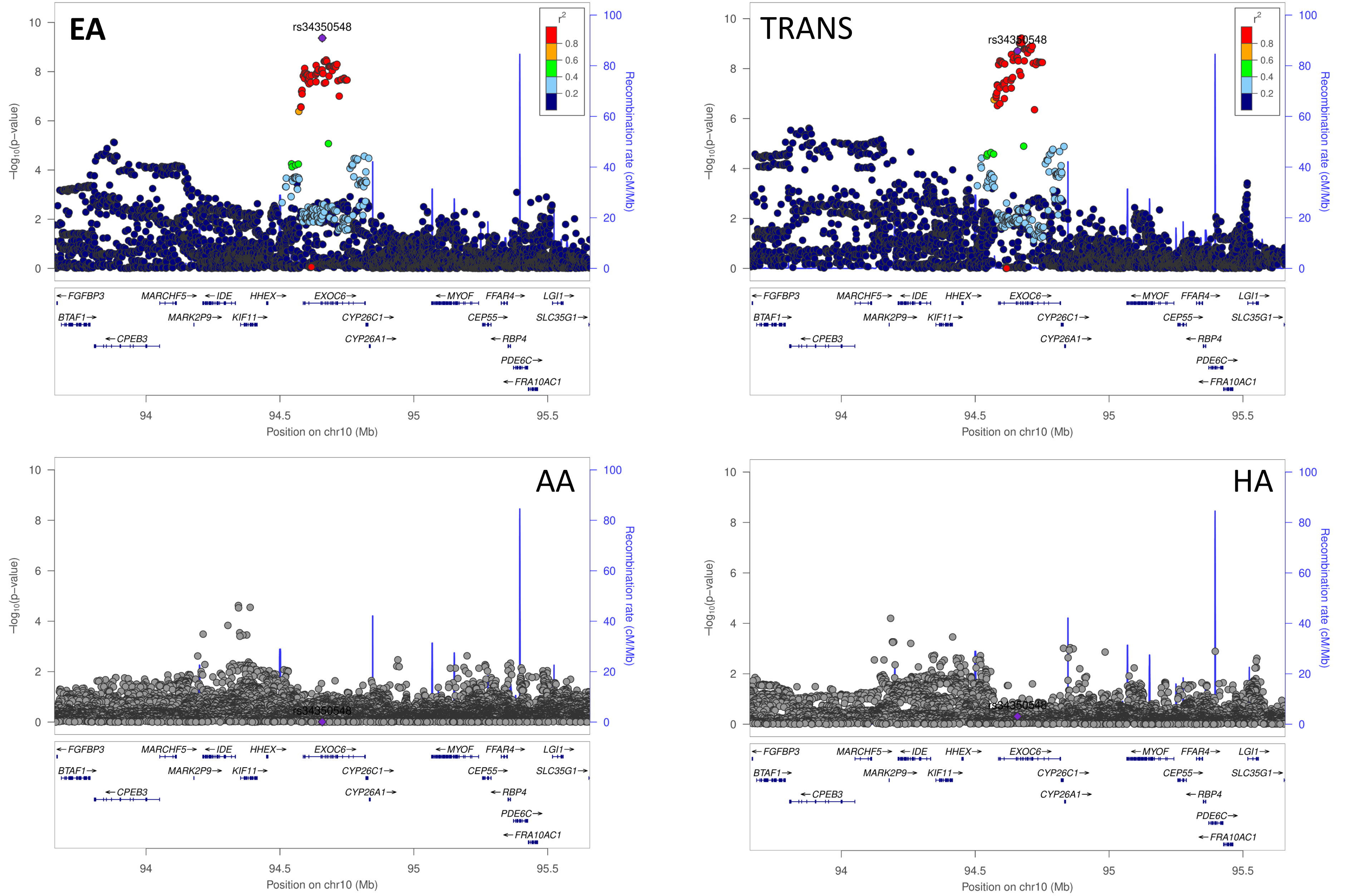


**Supplemental Figure 1o: Comparison of the European ancestry hit rs34350548 across European ancestry (EA), Transethnic ancestry (TRANS), African ancestry (AA), and Hispanic ancestry (HA) TBI studies.** Chromosomal position of the regional association plots is indicated on the x-axis, -log10 *p* values for each SNP (filled circles) is indicated on the y-axis, with the lead SNP shown in purple. Annotated genes in the region are drawn in the lower panel. Recombination rate is indicated by a blue line. Additional SNPs in the locus are colored according to linkage disequilibrium (*r*2) with the lead SNP.

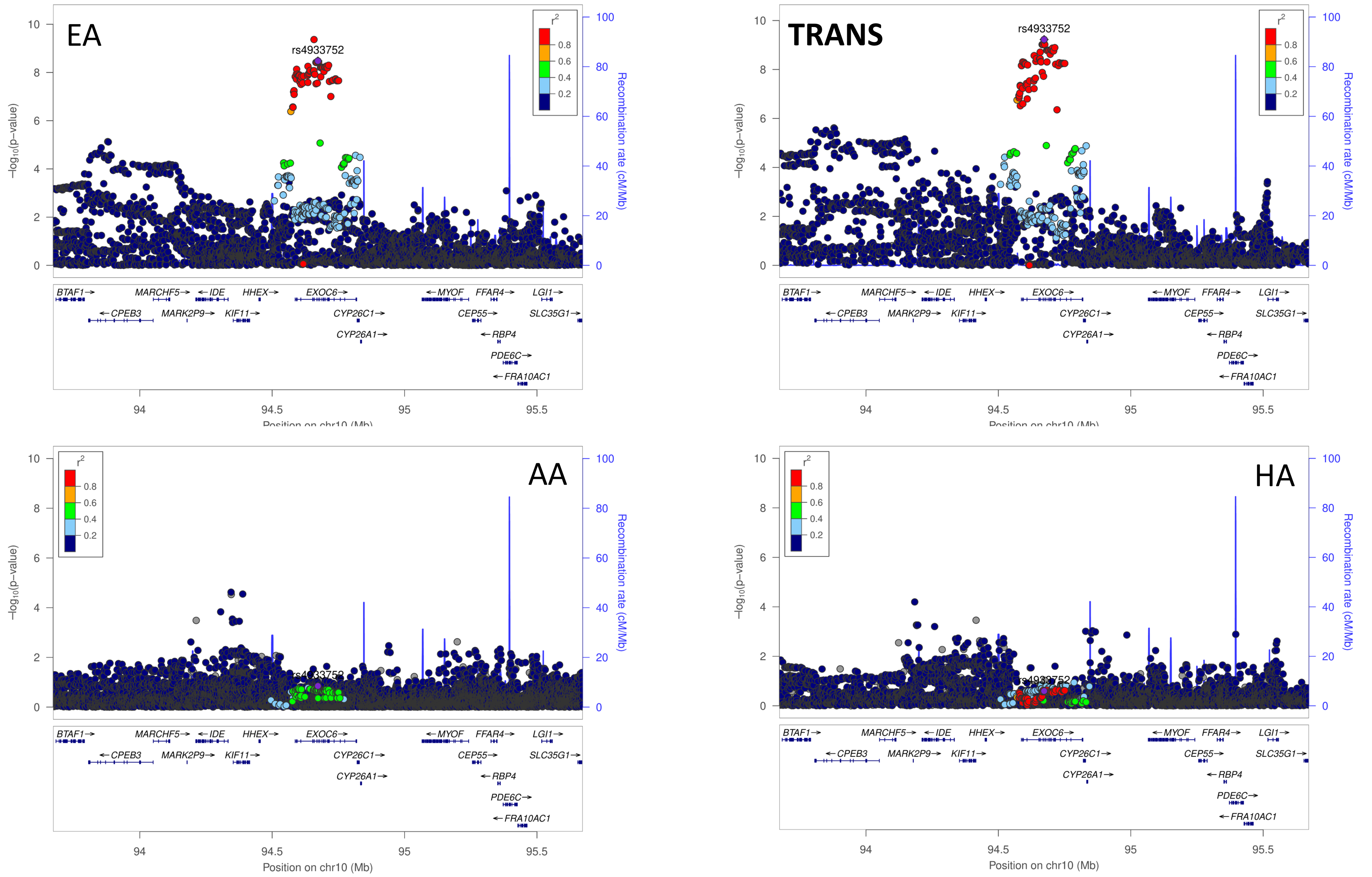


**Supplemental Figure 1p: Comparison of the European ancestry hit rs4933752 across European ancestry (EA), Transethnic ancestry (TRANS), African ancestry (AA), and Hispanic ancestry (HA) TBI studies.** Chromosomal position of the regional association plots is indicated on the x-axis, -log10 *p* values for each SNP (filled circles) is indicated on the y-axis, with the lead SNP shown in purple. Annotated genes in the region are drawn in the lower panel. Recombination rate is indicated by a blue line. Additional SNPs in the locus are colored according to linkage disequilibrium (*r*2) with the lead SNP.

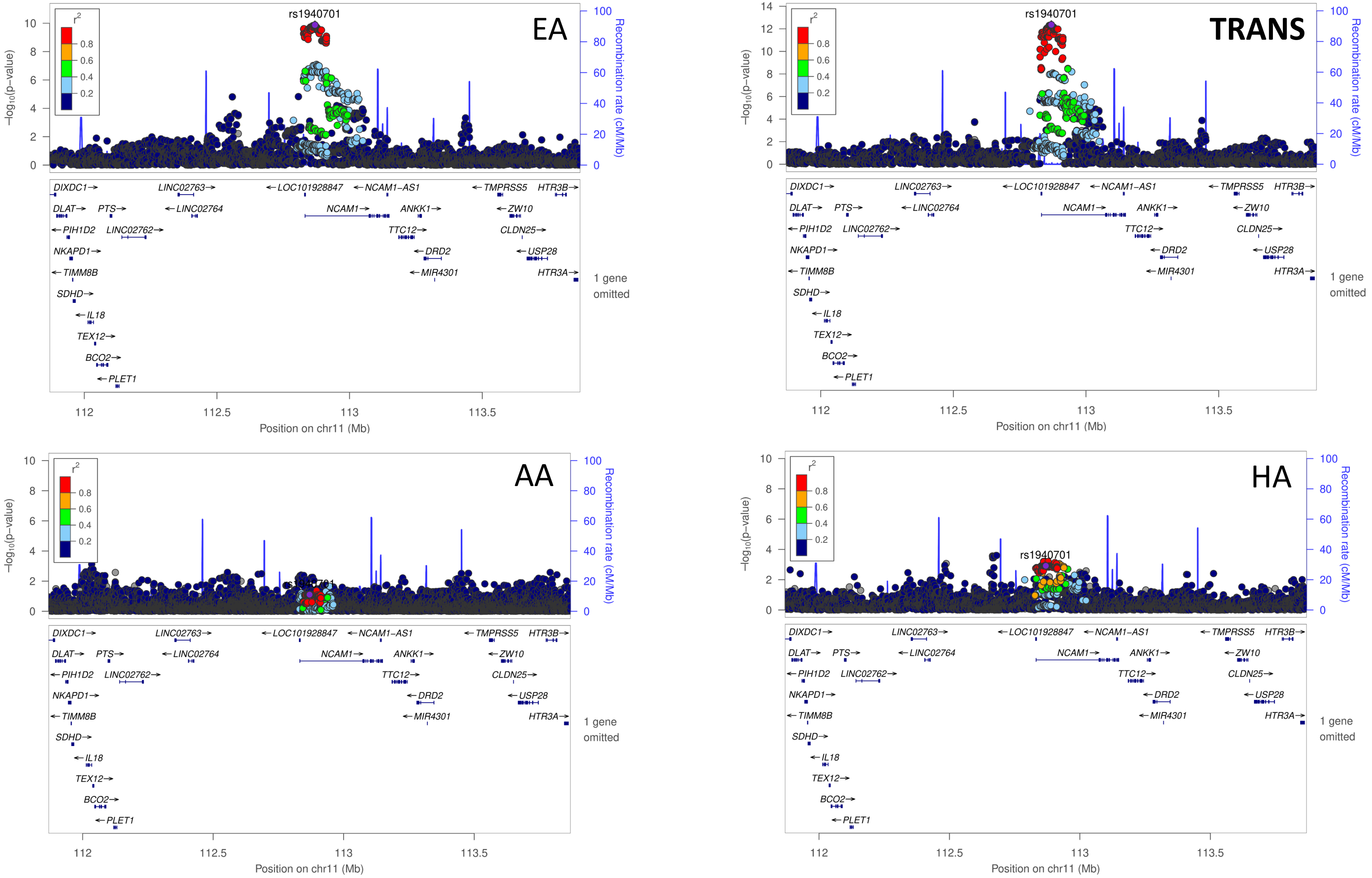


**Supplemental Figure 1q: Comparison of the European ancestry hit rs1940701 across European ancestry (EA), Transethnic ancestry (TRANS), African ancestry (AA), and Hispanic ancestry (HA) TBI studies.**
Chromosomal position of the regional association plots is indicated on the x-axis, -log10 *p* values for each SNP (filled circles) is indicated on the y-axis, with the lead SNP shown in purple. Annotated genes in the region are drawn in the lower panel. Recombination rate is indicated by a blue line. Additional SNPs in the locus are colored according to linkage disequilibrium (*r*2) with the lead SNP.

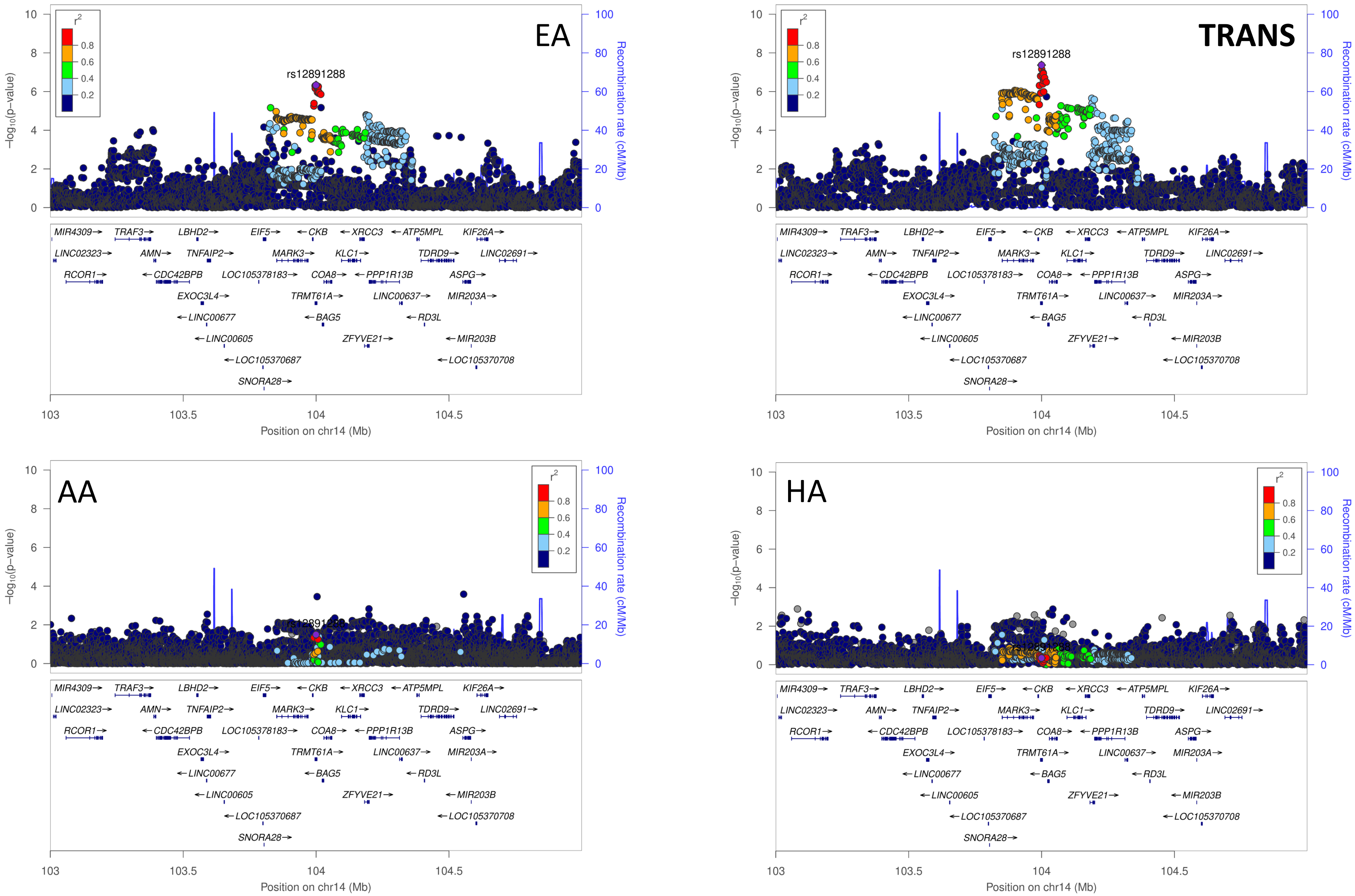


**Supplemental Figure 1r: Comparison of the European ancestry hit rs12891288 across European ancestry (EA), Transethnic ancestry (TRANS), African ancestry (AA), and Hispanic ancestry (HA) TBI studies.** Chromosomal position of the regional association plots is indicated on the x-axis, -log10 *p* values for each SNP (filled circles) is indicated on the y-axis, with the lead SNP shown in purple. Annotated genes in the region are drawn in the lower panel. Recombination rate is indicated by a blue line. Additional SNPs in the locus are colored according to linkage disequilibrium (*r*2) with the lead SNP.

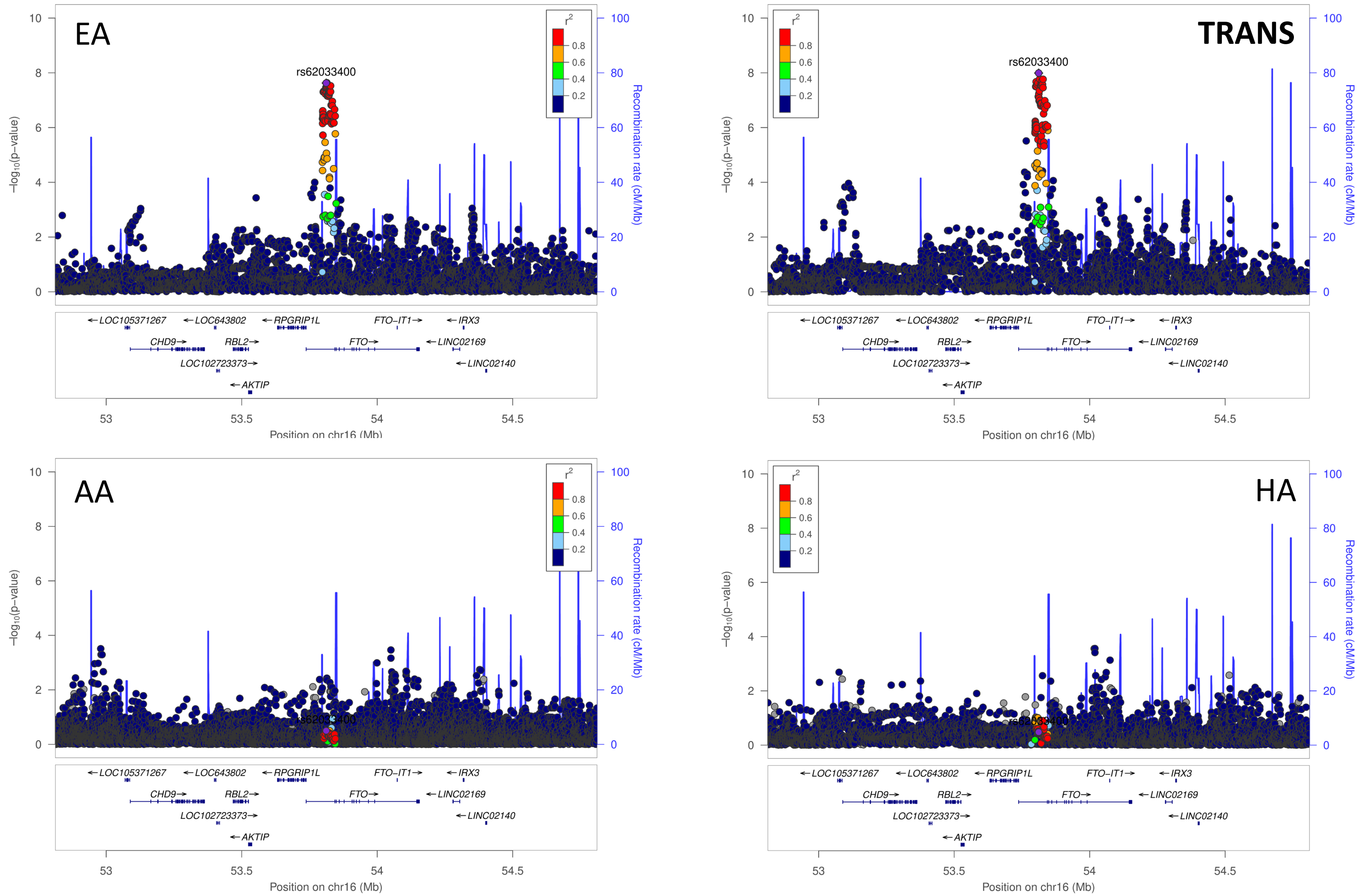


**Supplemental Figure 1s: Comparison of the European ancestry hit rs62033400 across European ancestry (EA), Transethnic ancestry (TRANS), African ancestry (AA), and Hispanic ancestry (HA) TBI studies.** Chromosomal position of the regional association plots is indicated on the x-axis, -log10 *p* values for each SNP (filled circles) is indicated on the y-axis, with the lead SNP shown in purple. Annotated genes in the region are drawn in the lower panel. Recombination rate is indicated by a blue line. Additional SNPs in the locus are colored according to linkage disequilibrium (*r*2) with the lead SNP.

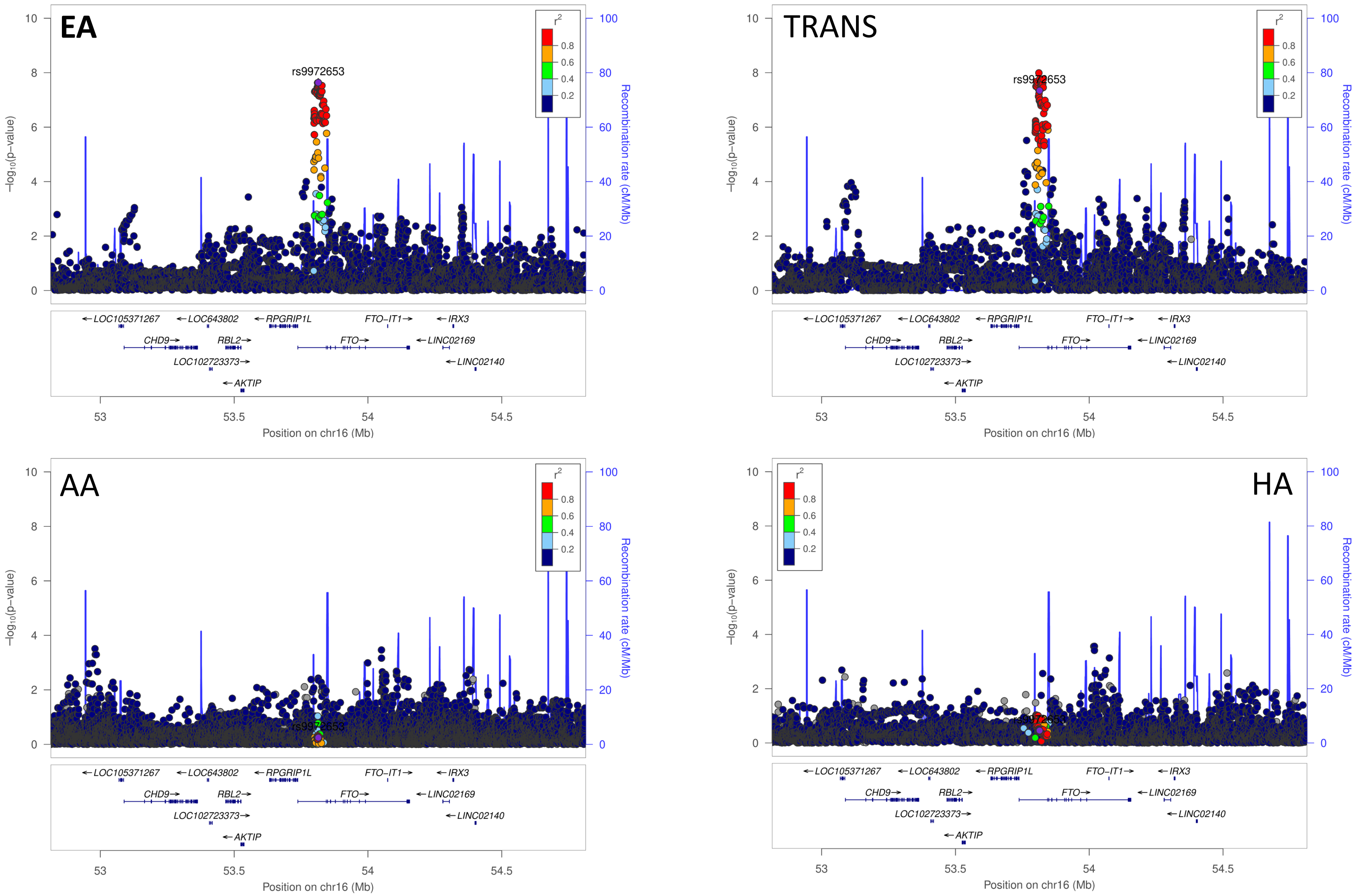


**Supplemental Figure 1t: Comparison of the European ancestry hit rs9972653 across European ancestry (EA), Transethnic ancestry (TRANS), African ancestry (AA), and Hispanic ancestry (HA) TBI studies.** Chromosomal position of the regional association plots is indicated on the x-axis, -log10 *p* values for each SNP (filled circles) is indicated on the y-axis, with the lead SNP shown in purple. Annotated genes in the region are drawn in the lower panel. Recombination rate is indicated by a blue line. Additional SNPs in the locus are colored according to linkage disequilibrium (*r*2) with the lead SNP.

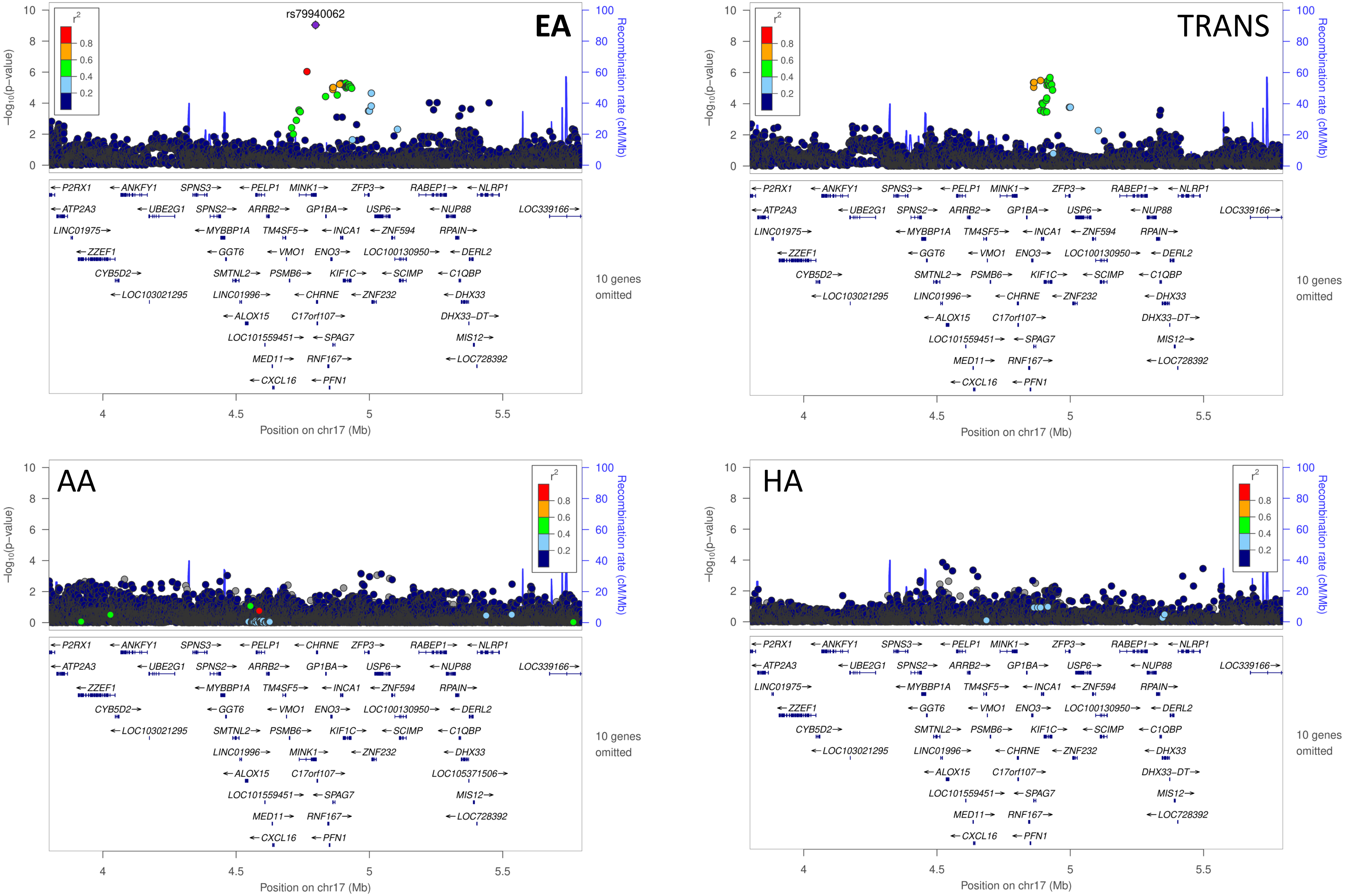


**Supplemental Figure 1u: Comparison of the European ancestry hit rs79940062 across European ancestry (EA), Transethnic ancestry (TRANS), African ancestry (AA), and Hispanic ancestry (HA) TBI studies.** Chromosomal position of the regional association plots is indicated on the x-axis, -log10 *p* values for each SNP (filled circles) is indicated on the y-axis, with the lead SNP shown in purple. Annotated genes in the region are drawn in the lower panel. Recombination rate is indicated by a blue line. Additional SNPs in the locus are colored according to linkage disequilibrium (*r*2) with the lead SNP.

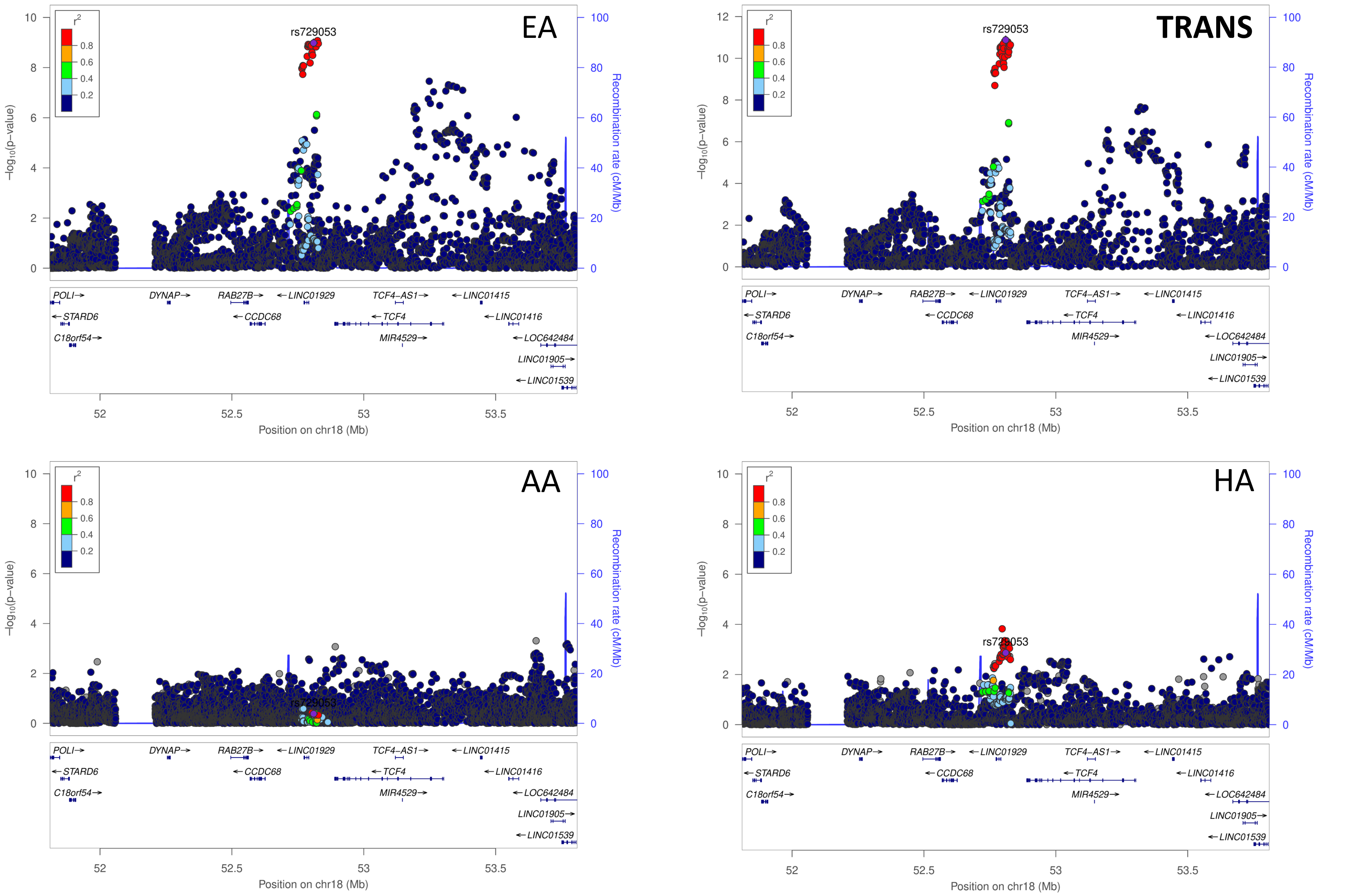


**Supplemental Figure 1v: Comparison of the European ancestry hit rs729053 across European ancestry (EA), Transethnic ancestry (TRANS), African ancestry (AA), and Hispanic ancestry (HA) TBI studies.** Chromosomal position of the regional association plots is indicated on the x-axis, -log10 *p* values for each SNP (filled circles) is indicated on the y-axis, with the lead SNP shown in purple. Annotated genes in the region are drawn in the lower panel. Recombination rate is indicated by a blue line. Additional SNPs in the locus are colored according to linkage disequilibrium (*r*2) with the lead SNP.

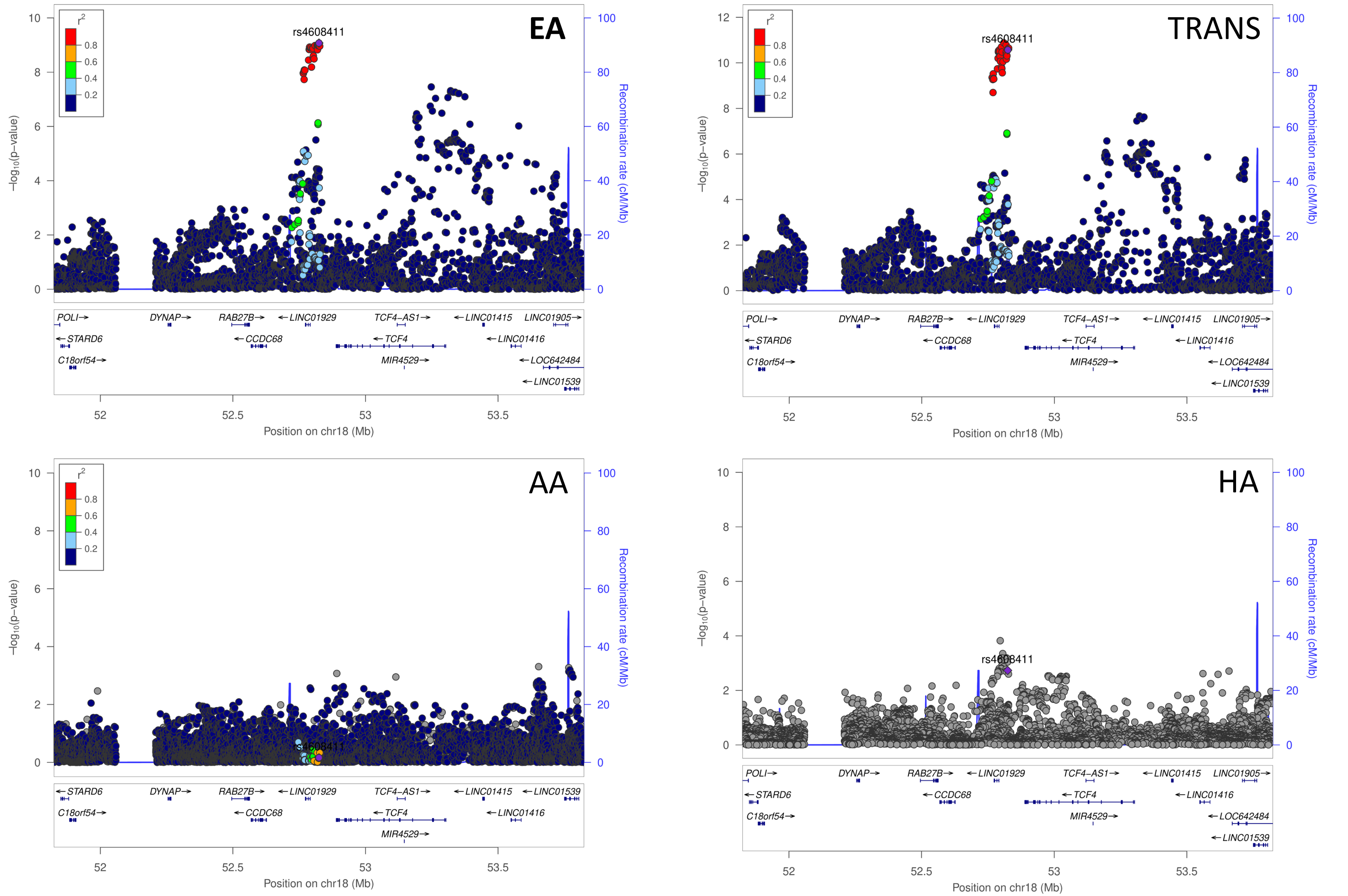


**Supplemental Figure 1w: Comparison of the European ancestry hit rs4608411 across European ancestry (EA), Transethnic ancestry (TRANS), African ancestry (AA), and Hispanic ancestry (HA) TBI studies.** Chromosomal position of the regional association plots is indicated on the x-axis, -log10 *p* values for each SNP (filled circles) is indicated on the y-axis, with the lead SNP shown in purple. Annotated genes in the region are drawn in the lower panel. Recombination rate is indicated by a blue line. Additional SNPs in the locus are colored according to linkage disequilibrium (*r*2) with the lead SNP.

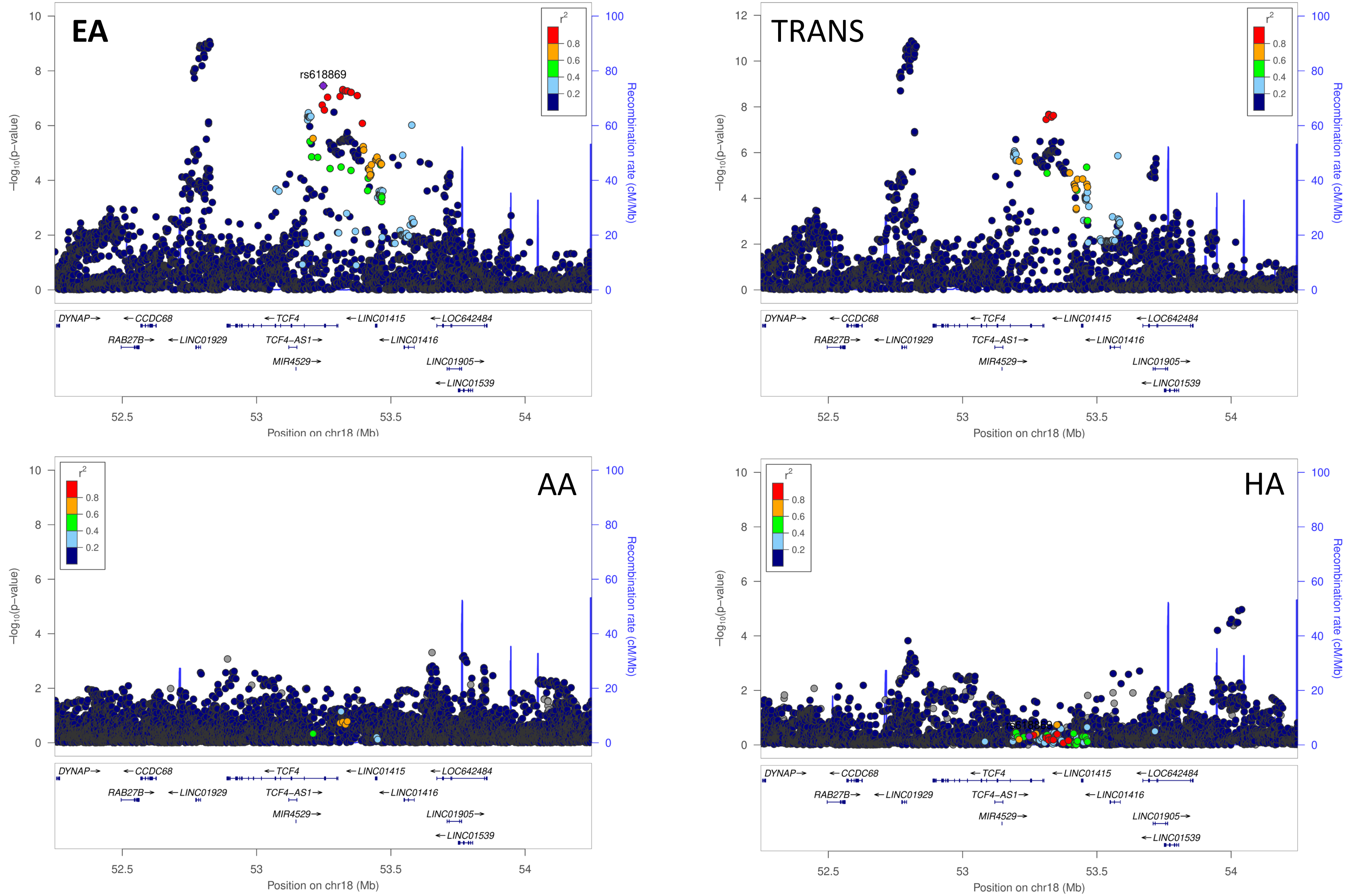


**Supplemental Figure 1x: Comparison of the European ancestry hit rs618869 across European ancestry (EA), Transethnic ancestry (TRANS), African ancestry (AA), and Hispanic ancestry (HA) TBI studies.** Chromosomal position of the regional association plots is indicated on the x-axis, -log10 *p* values for each SNP (filled circles) is indicated on the y-axis, with the lead SNP shown in purple. Annotated genes in the region are drawn in the lower panel. Recombination rate is indicated by a blue line. Additional SNPs in the locus are colored according to linkage disequilibrium (*r*2) with the lead SNP.

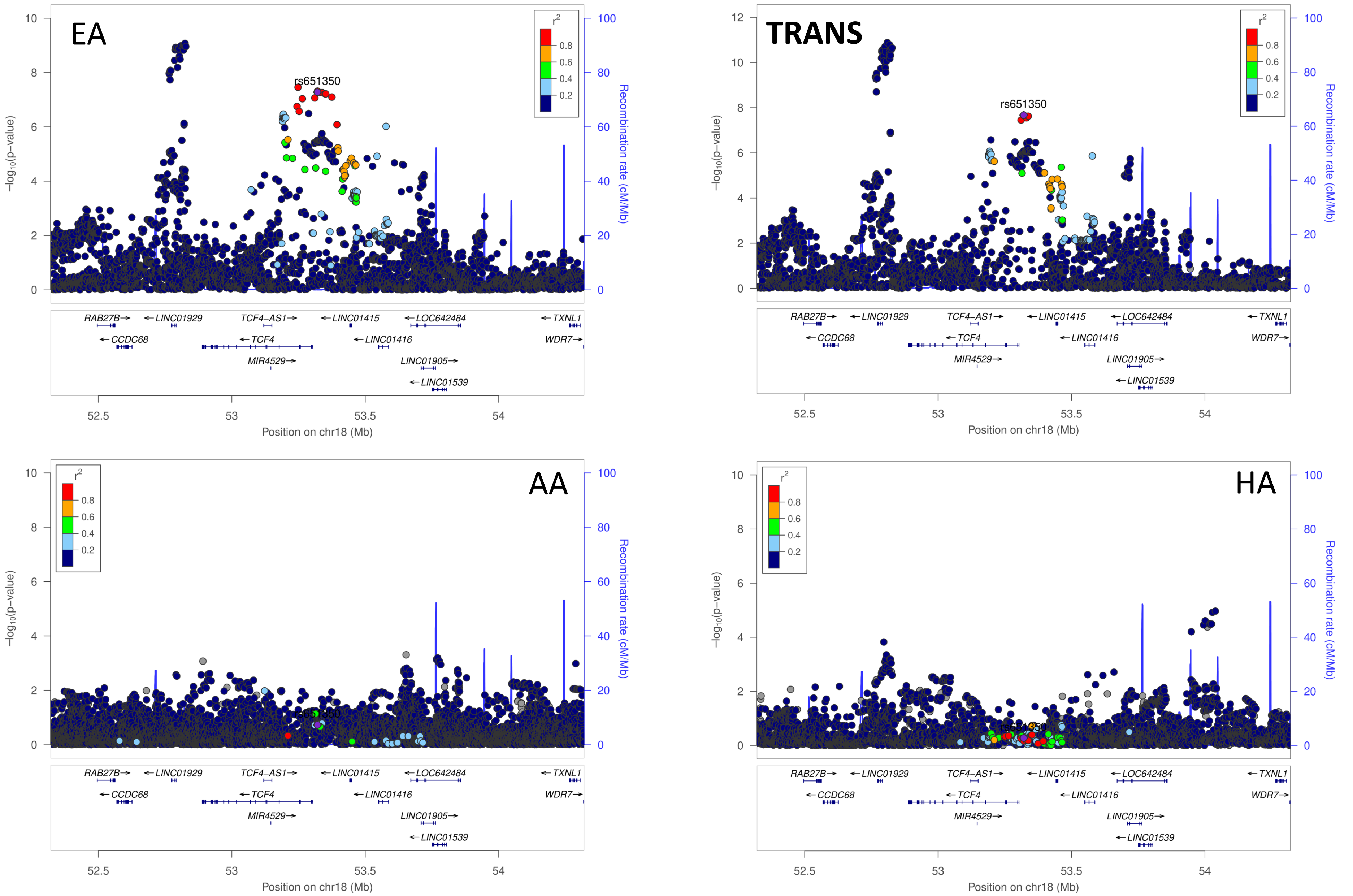


**Supplemental Figure 1y: Comparison of the European ancestry hit rs651350 across European ancestry (EA), Transethnic ancestry (TRANS), African ancestry (AA), and Hispanic ancestry (HA) TBI studies.** Chromosomal position of the regional association plots is indicated on the x-axis, -log10 *p* values for each SNP (filled circles) is indicated on the y-axis, with the lead SNP shown in purple. Annotated genes in the region are drawn in the lower panel. Recombination rate is indicated by a blue line. Additional SNPs in the locus are colored according to linkage disequilibrium (*r*2) with the lead SNP.

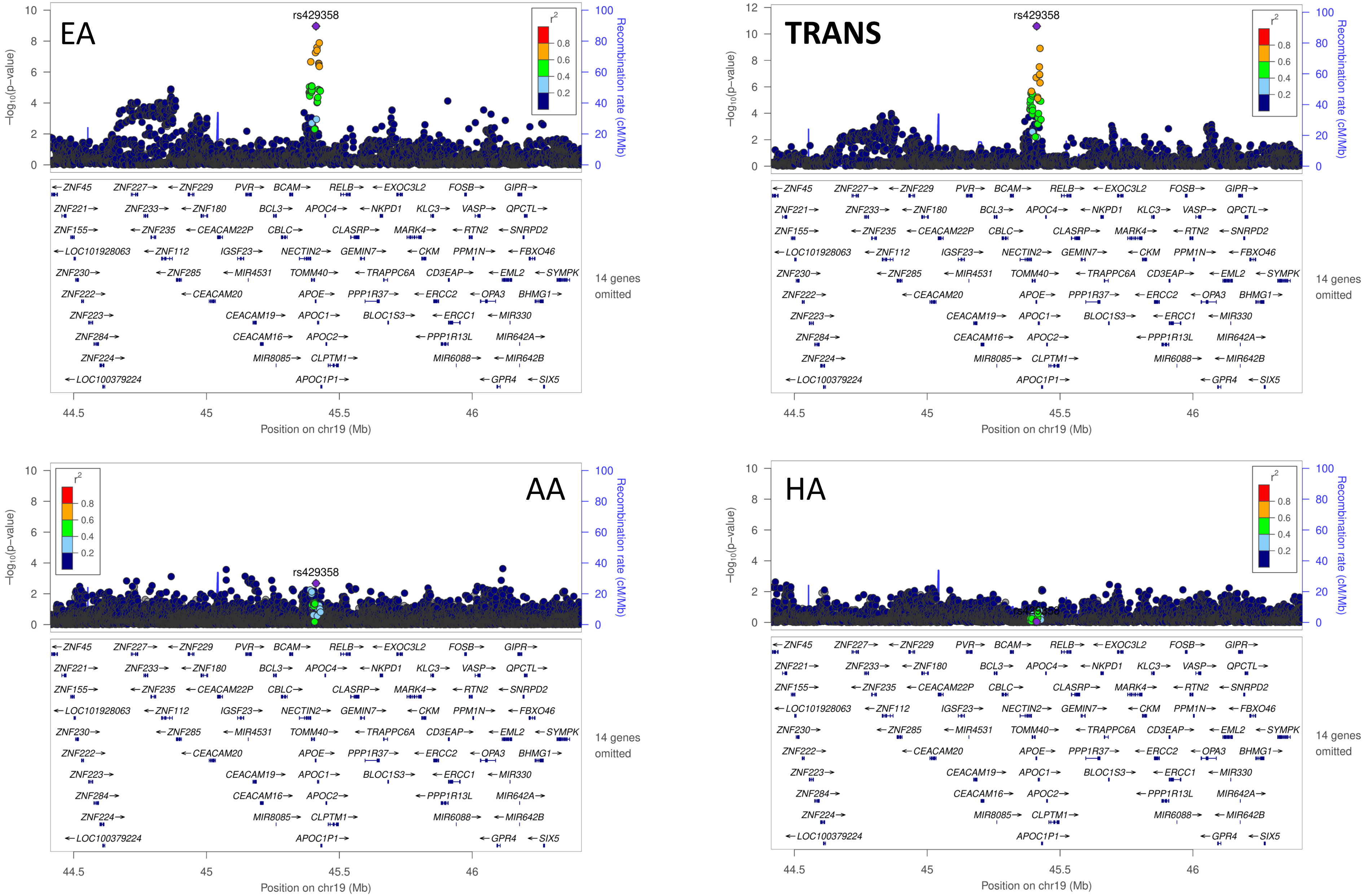


**Supplemental Figure 1z: Comparison of the European ancestry hit rs429358 across European ancestry (EA), Transethnic ancestry (TRANS), African ancestry (AA), and Hispanic ancestry (HA) TBI studies.** Chromosomal position of the regional association plots is indicated on the x-axis, -log10 *p* values for each SNP (filled circles) is indicated on the y-axis, with the lead SNP shown in purple. Annotated genes in the region are drawn in the lower panel. Recombination rate is indicated by a blue line. Additional SNPs in the locus are colored according to linkage disequilibrium (*r*2) with the lead SNP.

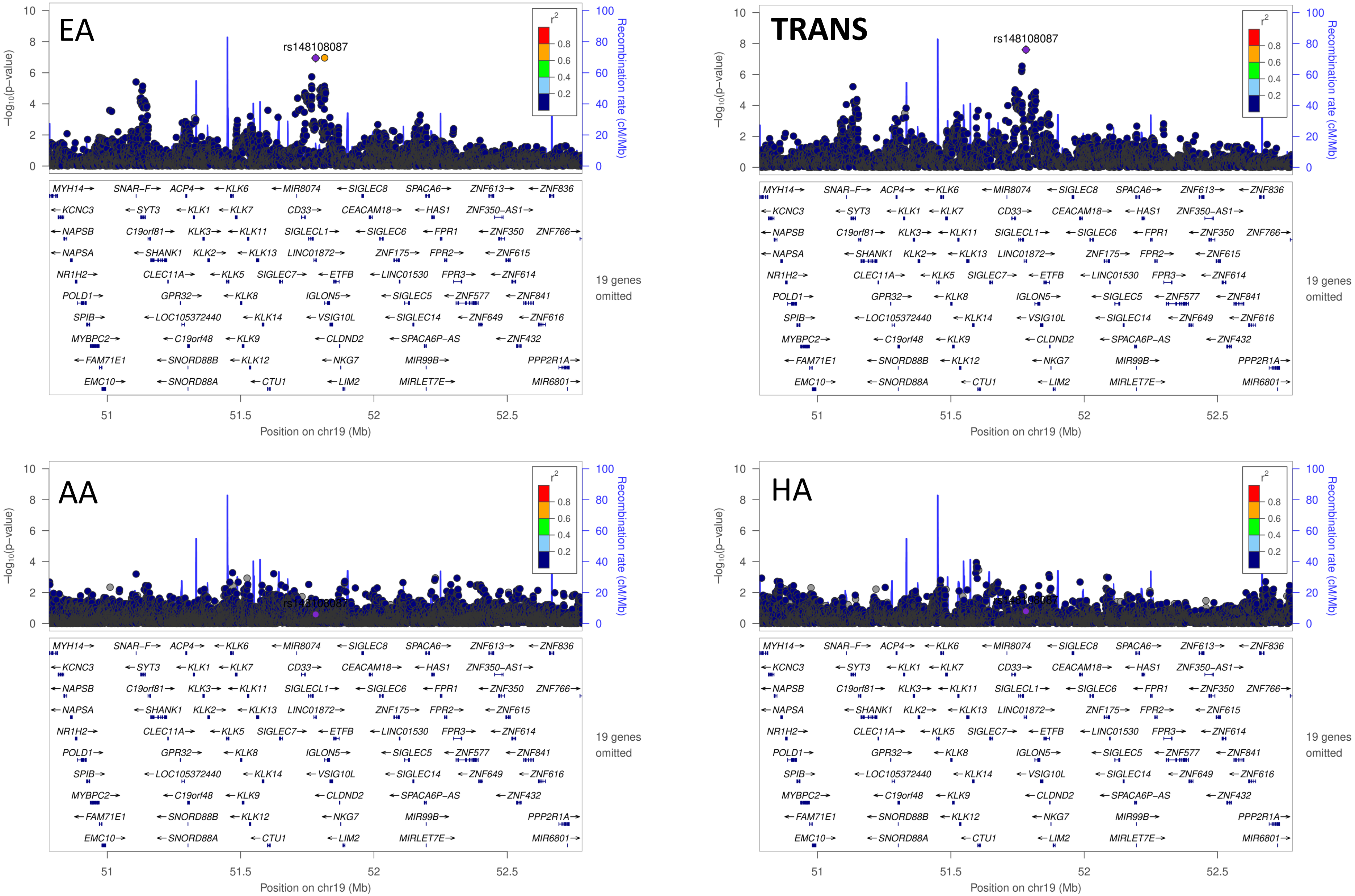


**Supplemental Figure 1aa: Comparison of the European ancestry hit rs148108087 across European ancestry (EA), Transethnic ancestry (TRANS), African ancestry (AA), and Hispanic ancestry (HA) TBI studies.** Chromosomal position of the regional association plots is indicated on the x-axis, -log10 *p* values for each SNP (filled circles) is indicated on the y-axis, with the lead SNP shown in purple. Annotated genes in the region are drawn in the lower panel. Recombination rate is indicated by a blue line. Additional SNPs in the locus are colored according to linkage disequilibrium (*r*2) with the lead SNP.

**Supplemental Figure 2.** Manhattan plot of TBI risk for the **European ancestry** cohort. The x axis displays the chromosome position, and the y axis shows the GWAS *p* value on a $-\log_{10}$ scale. The red dashed line reflects genome-wide significance at $p < 5 \times 10^{-8}$.

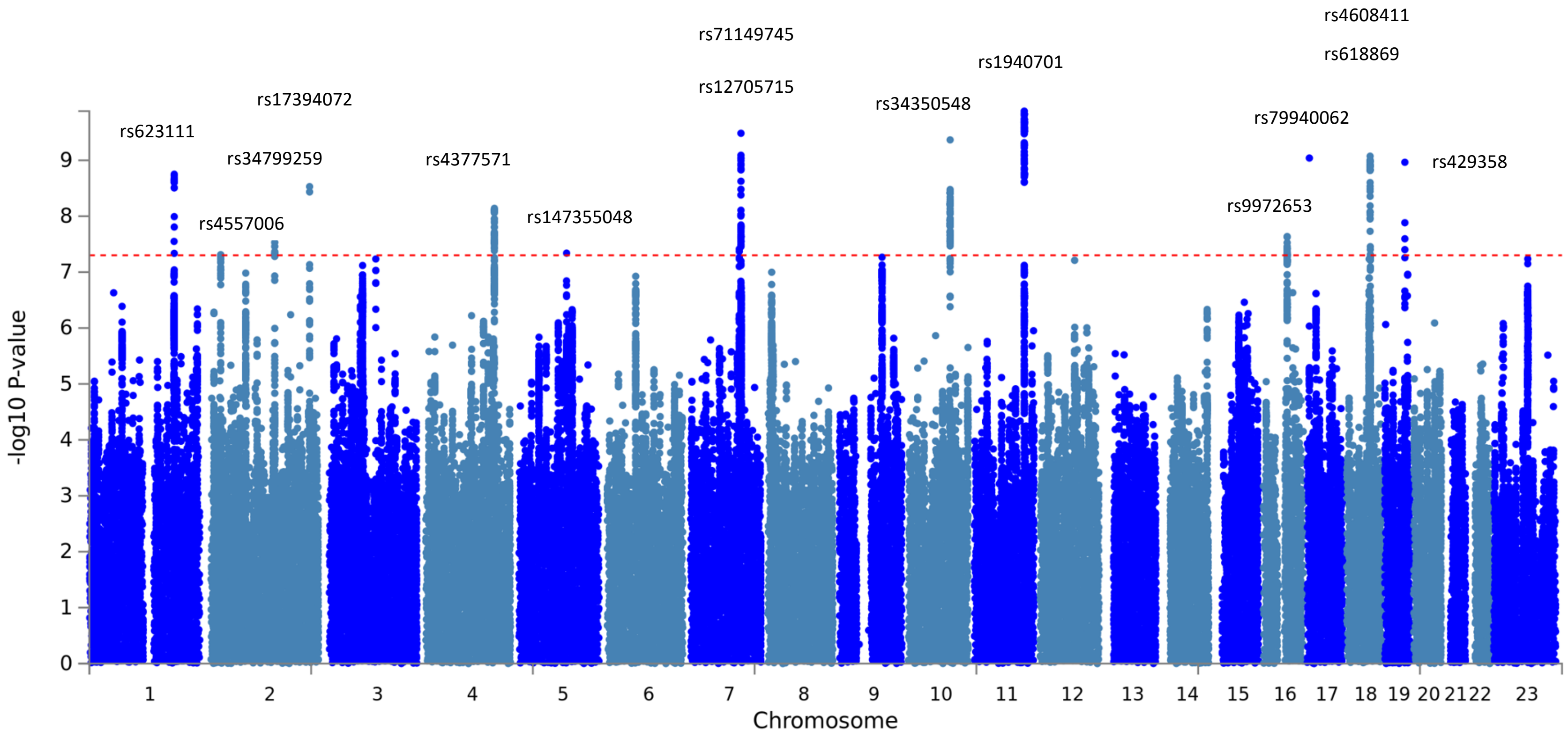

**Supplemental Figure 3.** Manhattan plot of TBI risk for the **African ancestry** cohort. The x axis displays the chromosome position, and the y axis shows the GWAS *p* value on a $-\log_{10}$ scale. The red dashed line reflects genome-wide significance at $p < 5 \times 10^{-8.}$

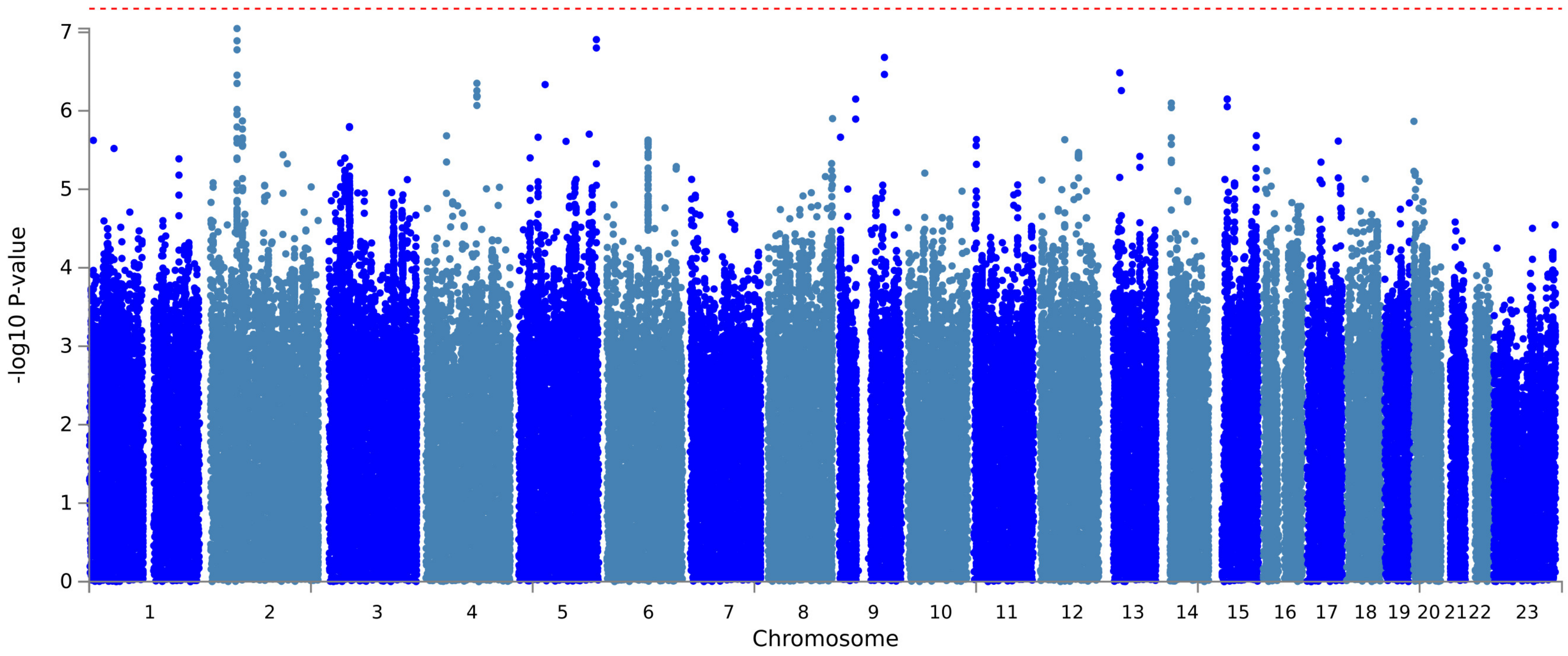

**Supplemental Figure 4.** Manhattan plot of TBI risk for the **Hispanic ancestry** cohort. The x axis displays the chromosome position, and the y axis shows the GWAS *p* value on a $-\log_{10}$ scale. The red dashed line reflects genome-wide significance at $p < 5 \times 10^{-8}$.

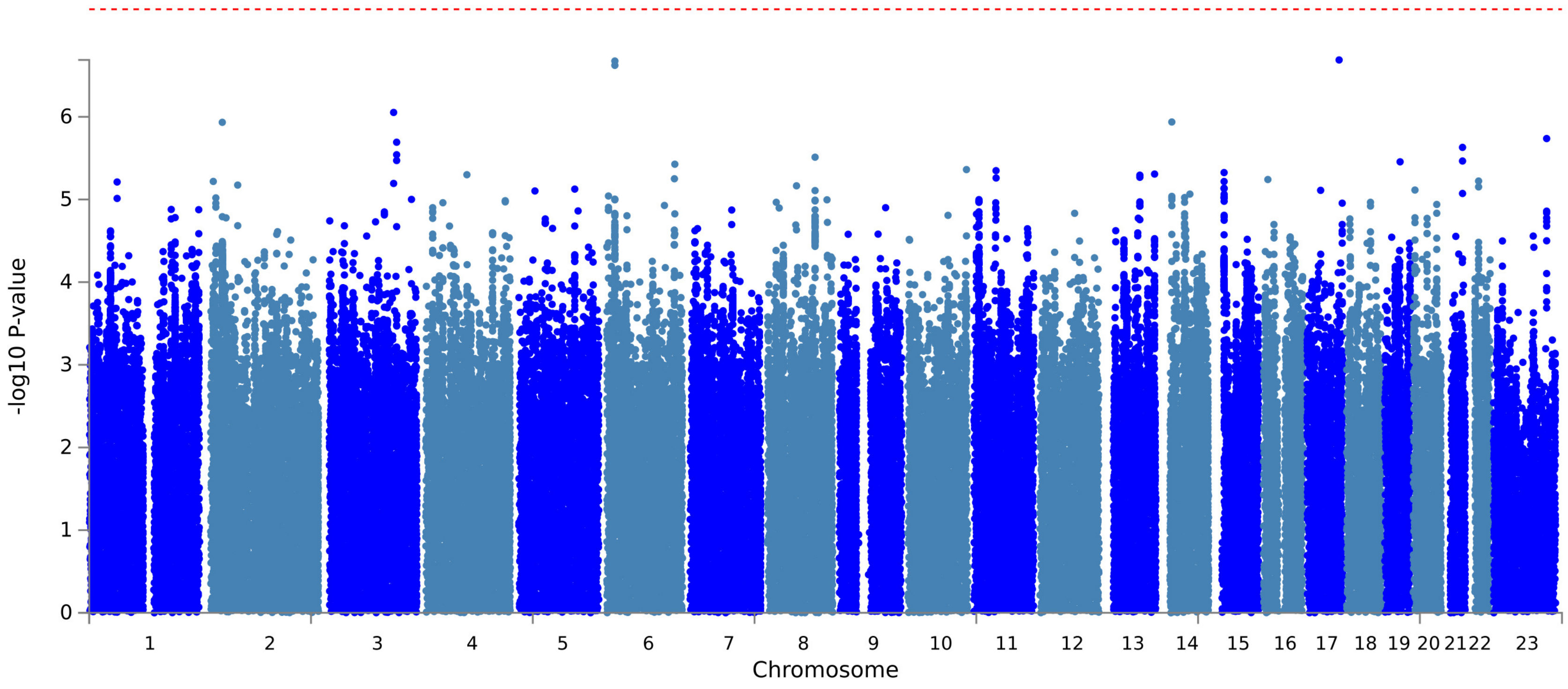

**Supplemental Figure 5a.** Gene-based results for multi-ancestry cohort.

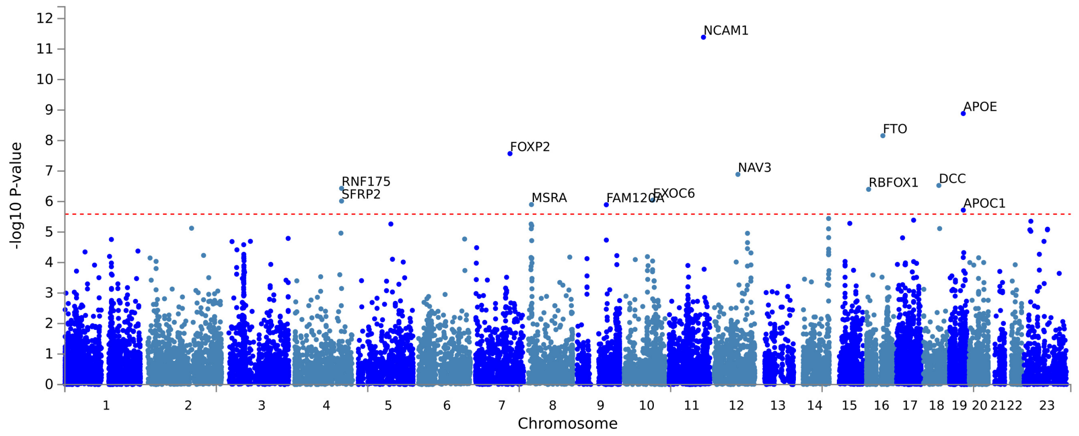


**Supplemental Figure 5b.** Gene-based results for European ancestry cohort.

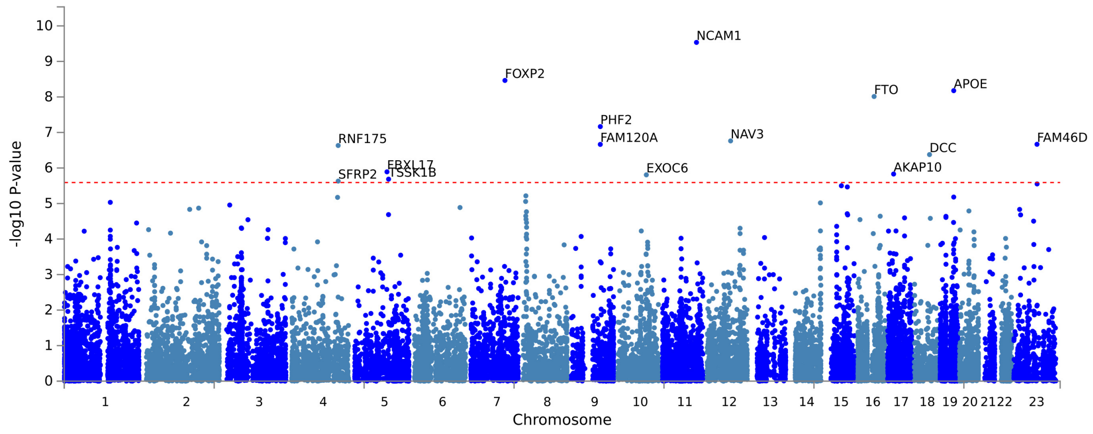

**Supplemental Figure 6a.** Gene tissue expression (tissues aggregated) in the **European ancestry** cohort.

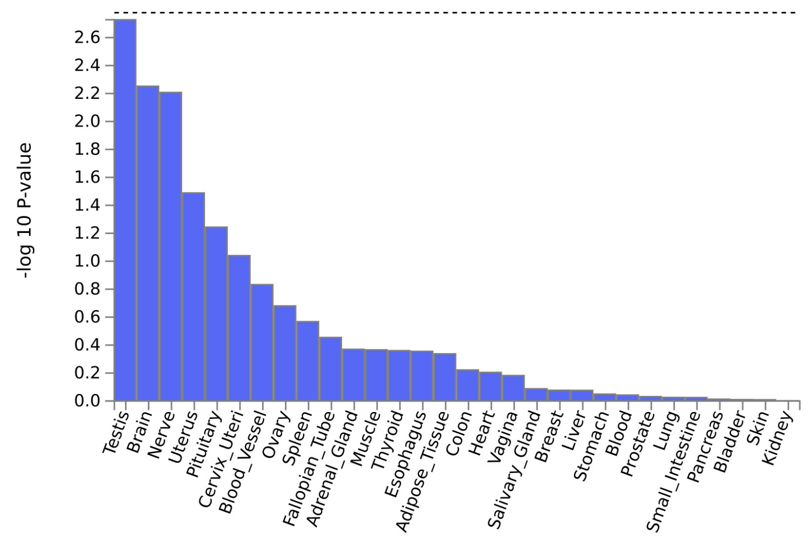


**Supplemental Figure 6b.** Gene tissue expression (stratified by tissue subtype) in the **European ancestry** cohort.

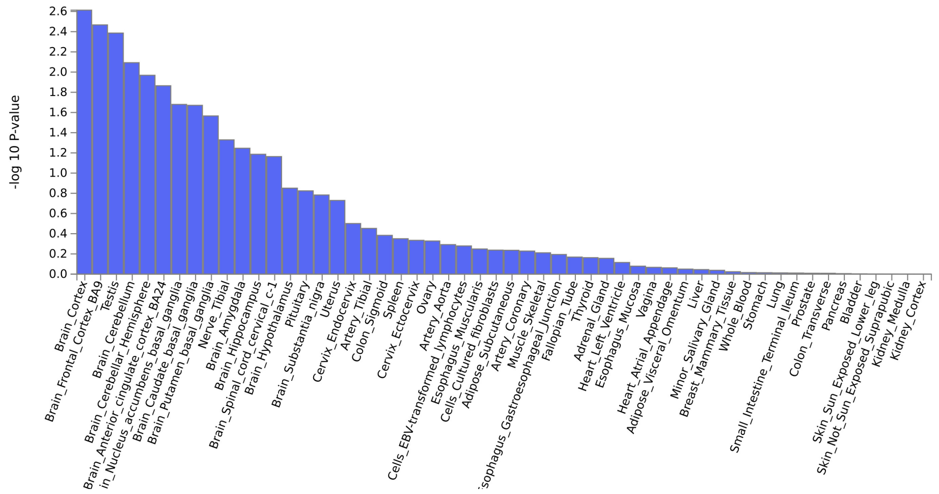

**Supplemental Figure 7.** LDSC correlations between TBI and risk-taking behaviors, psychiatric disorders, and neurocognition.

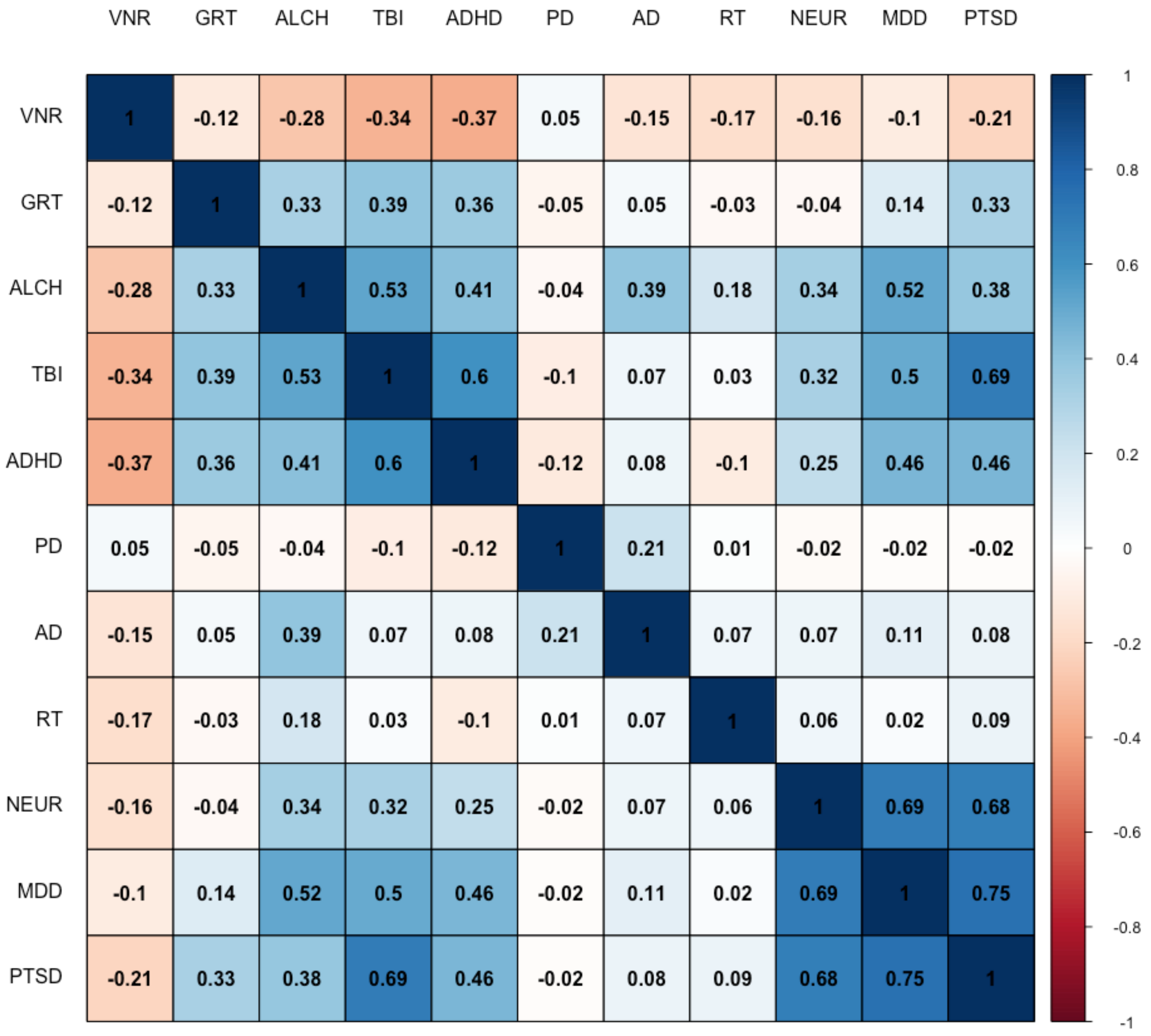

**Supplemental Figure 8.** LDSC correlations between TBI and ENIGMA variables.

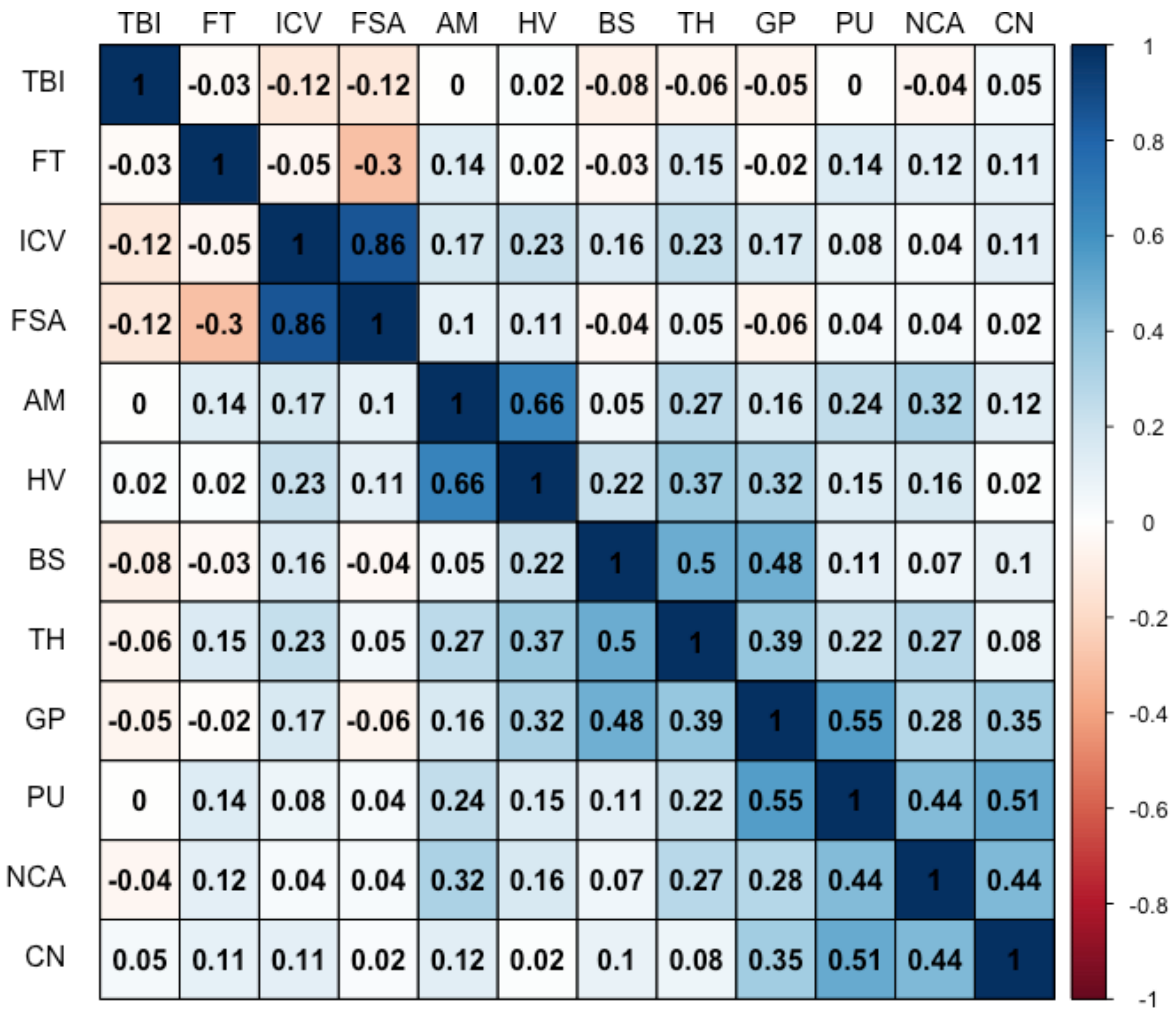

**Supplemental Figure 9.** Bivariate MiXeR analysis of TBI and Alzheimer's disease.

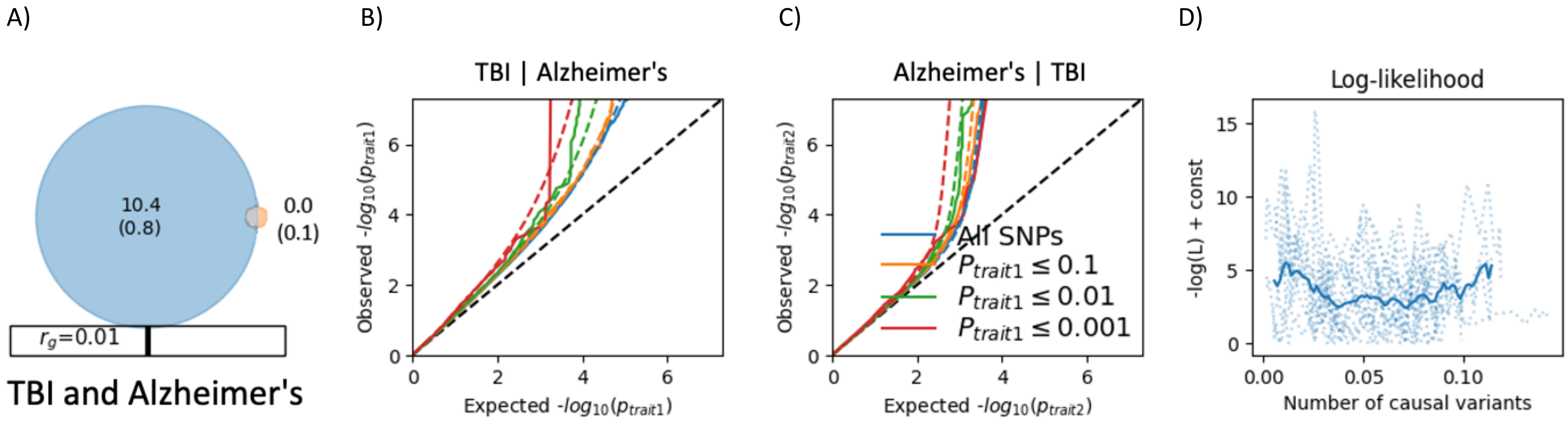


A) The Venn diagram depicts the number of causal variants (in thousands [standard error]) related to TBI (blue circle), Alzheimer's disease (orange circle), and the mutually shared variants (grey overlap). Genetic correlation is depicted as a number and red progress bar.

B) Conditional QQ plot of TBI conditional on Alzheimer's.

C) Conditional QQ plot of Alzheimer's conditional on TBI.

D) Model log likelihoods based on the number of causal variants. The solid blue line reflects the average likelihood across the 20 MiXeR runs. Dotted blue lines reflect likelihoods of individual runs.

**Supplemental Figure 10.** Bivariate MiXeR analysis of TBI and reaction time.

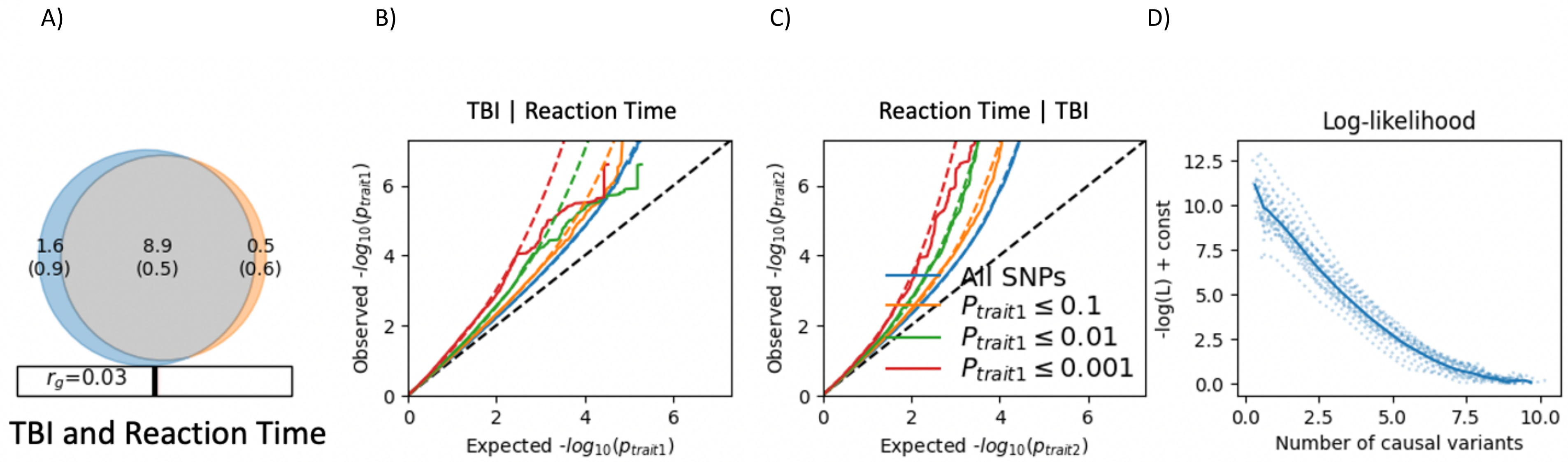


A) The Venn diagram depicts the number of causal variants (in thousands [standard error]) related to TBI (blue circle), reaction time (orange circle), and the mutually shared variants (grey overlap). Genetic correlation is depicted as a number and red progress bar.
B) Conditional QQ plot of TBI conditional on reaction time.
C) Conditional QQ plot of reaction time conditional on TBI.
D) Model log likelihoods based on the number of causal variants. The solid blue line reflects the average likelihood across the 20 MiXeR runs. Dotted blue lines reflect likelihoods of individual runs.

**Supplemental Figure 11.** PheWAS plots for European ancestry cohort.

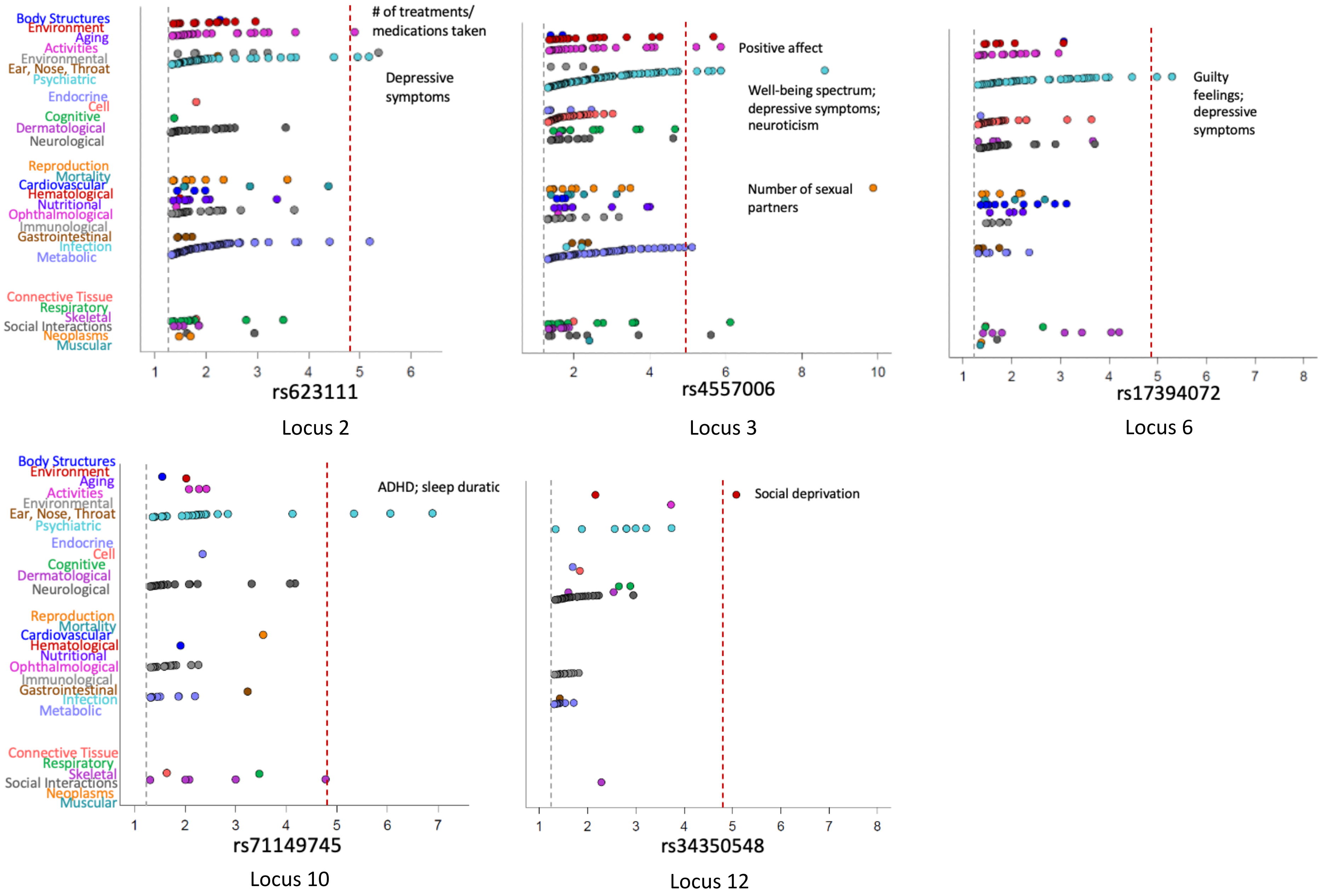

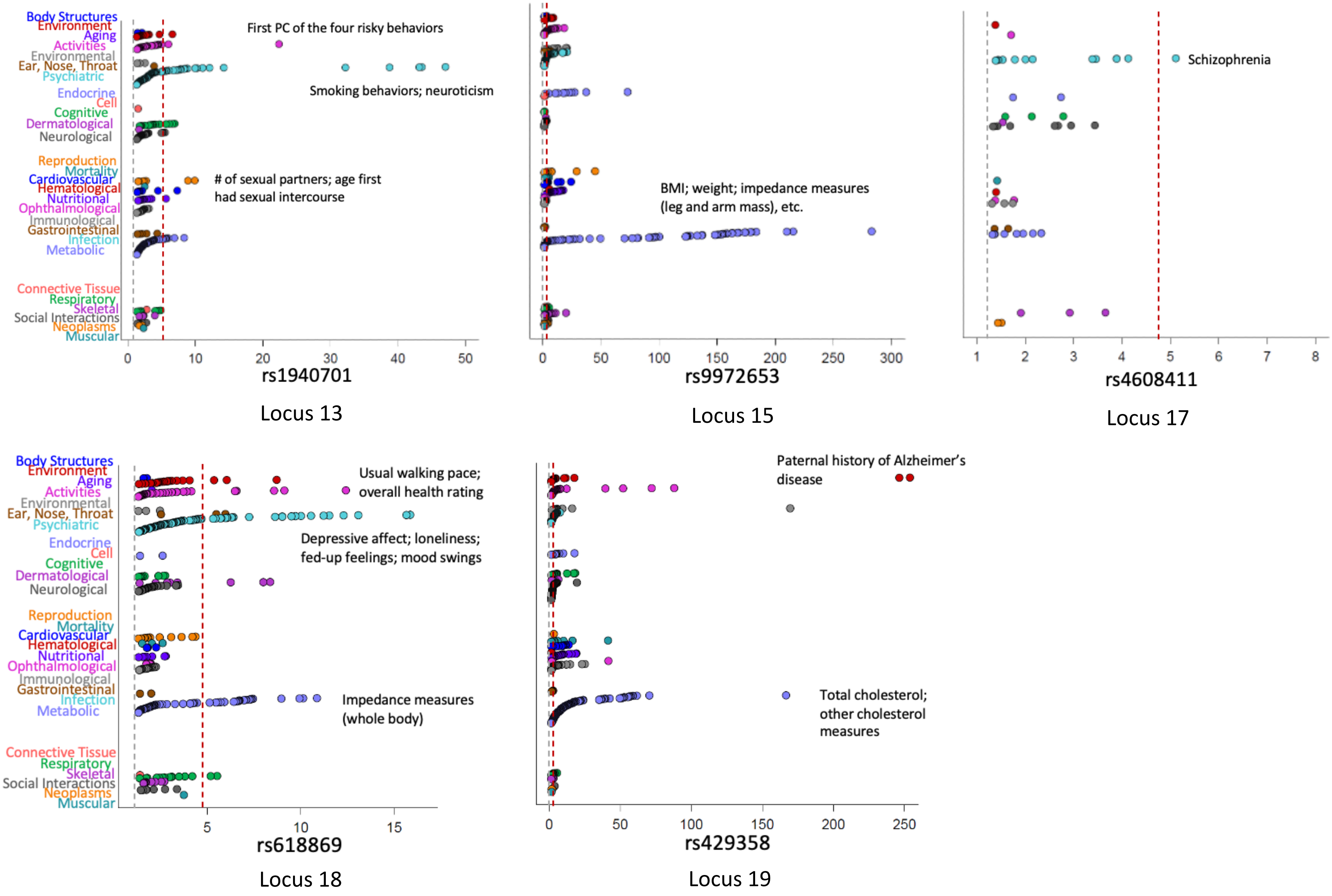
Body Structures
Environment
Aging
Activities
Environmental
Ear, Nose, Throat
Psychiatric
Endocrine
Cell
Cognitive
Dermatological
Neurological
Reproduction
Mortality
Cardiovascular
Hematological
Nutritional
Ophthalmological
Immunological
Gastrointestinal
Infection
Metabolic
Connective Tissue
Respiratory
Skeletal
Social Interactions
Neoplasms
Muscular
First PC of the four risky behaviors
Smoking behaviors; neuroticism
### of sexual partners; age first had sexual intercourse
0 10 20 30 40 50
rs1940701
Locus 13
BMI; weight; impedance measures (leg and arm mass), etc.
0 50 100 150 200 250 300
rs9972653
Locus 15
Schizophrenia
1 2 3 4 5 6 7 8
rs4608411
Locus 17
Usual walking pace; overall health rating
Depressive affect; loneliness; fed-up feelings; mood swings
Impedance measures (whole body)
5 10 15
rs618869
Locus 18
Paternal history of Alzheimer's disease
Total cholesterol; other cholesterol measures
0 50 100 150 200 250
rs429358
Locus 19

**Supplemental Figure 12. PheWAS results for TBI risk loci.**

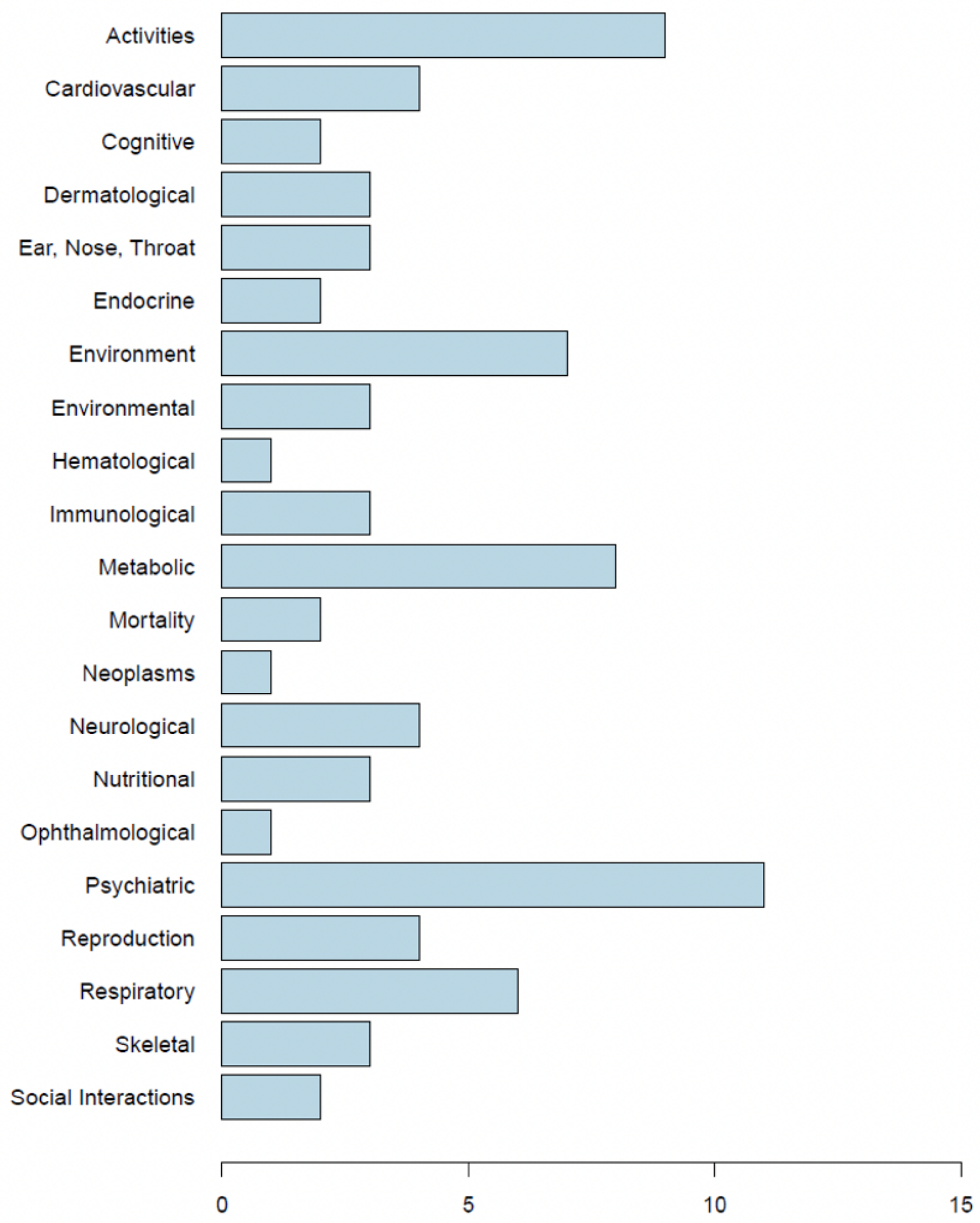


Associations from the PheWAS of 15 TBI loci in the GWAS Atlas. Phenotypes were grouped into broad categories. The X-axis depicts the frequency of loci within each category.
